# Development of a conceptual framework to guide description and evaluation of social interventions for people with serious mental health conditions

**DOI:** 10.1101/2023.01.26.23284827

**Authors:** Rebecca Appleton, Phoebe Barnett, Beverley Chipp, Michael Clark, Peter Goldblatt, Stephen Jeffreys, Karen Machin, Justin J Needle, Prisha Shah, Georgina Thompson, Kylee Trevillion, Martin Webber, Minnie Worden, Sonia Johnson, Brynmor Lloyd-Evans

## Abstract

People with serious mental health conditions face social exclusion and have poorer social outcomes compared to the general population in several areas of life. Social exclusion also negatively impacts mental health. Promising models of support to improve social outcomes for people with serious mental health conditions have been described in the literature and proliferate in practice, but typologies of support are not clearly established and a robust evidence base for effective approaches is lacking in many areas. We conducted a scoping review of relevant literature and consulted with experts in the field to identify models to improve social circumstances across eight life domains, with the aim of developing a conceptual framework to distinguish the main broad approaches to improving the social circumstances of people with serious mental health conditions. We also sought to explore which approaches have been used in models within each life domain, and to detect any preliminary indications of which approaches are most promising, based on evaluations of identified models of support. This work was conducted in collaboration with a group of expert stakeholders, including people with lived experience of accessing mental health services. We developed a conceptual framework which distinguishes sources and types of support, allowing description of complex interventions to improve the social circumstances of people with serious mental health problems, and providing a framework to guide future service development and evaluation.

## 1. Introduction

People with serious mental health problems face social exclusion and have poorer social outcomes than the general population in several areas of life (Boardman, 2022, Payne, 2006). Being socially excluded means that they are not able to participate in key activities usual in their society; for example, they might not have stable housing or employment (Boardman, 2011). There is a growing body of literature which recognises the importance of addressing the social needs of people at risk of social exclusion (Johnson, 2017, Webber and Fendt-Newlin, 2017, Wahlbeck et al., 2017).

Individuals with serious mental health problems have increased risks of social difficulties across most life areas. They have high rates of unemployment, despite often wanting work (Lloyd-Evans et al., 2013, Gühne et al., 2021) and can face barriers to employment such as stigma and discrimination (Centre for Mental Health, 2013). They are also at more risk of poverty and debt (Elliott, 2016, Royal College of Psychiatrists, 2022) and are more likely to be homeless (Local Government Association, 2017) and become victims of crime (Teplin et al., 2005, Desmarais et al., 2014, Khalifeh et al., 2015, Sariaslan et al., 2020). Research also suggests that people with a psychiatric diagnosis are more likely than the general population to be involved in crime perpetration (Sariaslan et al., 2020). Social isolation and loneliness are also commonly reported unmet needs in people with serious mental health problems (Fortuna et al., 2019, Meltzer et al., 2013, Palumbo et al., 2015) who typically have smaller social networks and fewer friends than the general population (Palumbo et al., 2015).

The relationship between social challenges and mental health is bi-directional, as social exclusion can also have a detrimental effect on mental health (Almquist et al., 2017, Allen et al., 2014). Service users have indicated that improving their social circumstances, for example, through the provision of housing or social support, is a key factor in their recovery (Law and Morrison, 2014). It is therefore important that people with serious mental health problems are able to access not only clinical interventions, but also forms of support which focus on meeting their social needs.

For many areas of life, however, there is a lack of evidence-based models to meet the social needs of this population group. Two recent systematic reviews (Barnett et al., 2022, Killaspy et al., 2022) identified a few demonstrably effective programmes (for example, Individual Placement and Support (IPS) and Housing First) and a number of promising interventions which currently lack a robust evidence base beyond small-scale studies establishing feasibility and acceptability. In practice, a variety of untested social interventions proliferate (Johnson, 2017). There is increasing recognition of the need for more research into interventions to improve social outcomes for people with serious mental health conditions, in order to boost the social element of a biopsychosocial treatment paradigm (Priebe et al., 2013).

However, research into social interventions is hampered by the lack of a clear conceptual framework for describing, comparing and evaluating different approaches. Many interventions are complex and may target more than one social outcome, and many share similar components but use different terminology to describe them. To guide future research into models of support to improve social outcomes for people with mental health conditions, we need to be able to describe and compare the wide range of models of support found in the academic literature and in practice, most of which as yet lack robust evaluation.

In this paper we report the development of a conceptual framework to guide the description and evaluation of models of support to improve the social circumstances of people with serious mental health conditions, informed by a literature review and iterative expert consultation. The term ‘serious mental health condition’ is used to refer to any mental health problem of a severity which would typically require intervention from secondary mental health services, regardless of diagnosis. We explored models of support across eight life domains: housing and homelessness, money and basic needs, work and education, social isolation and connectedness, family and caring relationships, victimisation and exploitation, offending, and rights, inclusion and citizenship. The aims of the study were:

1. To develop a conceptual framework to distinguish broad approaches to improving the social circumstances of people with a serious mental health condition.
2. To test whether the components of the framework are necessary and sufficient to describe existing models of support for serious mental health conditions, by using it to describe the components of interventions identified through a scoping review of published literature and an expert consultation.
3. To explore which approaches have been used by models within each life domain, and any preliminary indications of most promising approaches, with reference to existing evaluations of identified social interventions.

## 2. Methods

We conducted a scoping review of existing international literature and an expert consultation to identify models of support to improve social circumstances across eight life domains in people with serious mental health conditions. This informed the iterative development of a conceptual framework to describe and categorise broad approaches of models of support for serious mental health conditions, repeatedly refining the framework in discussion in a stakeholder working group including experts by experience, service providers and academic experts.

### 2.1 Consultation on life domains to be included

A stakeholder working group, comprising academics, practitioners, people with lived experience of mental health problems, and policymakers from the Department of Health and Social Care (DHSC) and Public Health England, was assembled for the review. The group met regularly throughout the project and oversaw all stages of the work. The eight domains used in this review were chosen by our working group as they were identified as important for people’s quality of life and as health determinants. They were also previously used in a systematic review of trials of social interventions by members of our research team (Barnett et al., 2022). These domains were the focus of an initial scoping review to identify social interventions for people with serious mental health conditions (Table 1).

**Table 1:** Life domains included in the scoping review.

| Life domain | Relevant social circumstances |
| --- | --- |
| Housing and homelessness | Housing instability (achieving and sustaining tenancies)<br>Housing quality (individual housing and neighbourhoods) |
| Money and basic needs | Poverty/income<br>Financial barriers to essential resources (including food and fuel poverty and availability, access to transport)<br>Debt<br>Money management and education |
| Work and education | Precarious work (insecure, erratic or low-paid work which offers compensation, hours or security inferior to regular employment)<br>Lack of access to or completion of educational goals<br>Lack of access to voluntary work, or paid work through sheltered employment or social firms (for those who do not want competitive paid employment)<br>Length of illness absence/time to return to work from sick leave due to mental health conditions |
| Social isolation and connectedness | Subjective social isolation/loneliness<br>Objective social isolation/social network<br>Individual social capital (resources that individuals gain as a result of their membership of social networks) |
| Family relationships | Partner/sexual relationships (achieving or sustaining a relationship)<br>Maintaining or developing desired parenting roles or contact with children<br>Family relationships (maintaining desired contact or cohabitation with family members, however defined)<br>Caring responsibilities (maintaining caring role) |
| Victimisation and exploitation | Victim of crime (general)<br>Sexual assault<br>Domestic violence and coercive control<br>Exploitation, harassment and safeguarding concerns |
| Offending | Risk of offending (prevention/diversion from offending)<br>Transition from prison to community<br>Reoffending |
| Rights, inclusion and citizenship | Social exclusion and social participation (including digital exclusion)<br>Difficulties with access to public services including welfare benefits<br>Immigration status (resolution of status, access to support)<br>Lack of privacy or dignity resulting from social circumstances |

### 2.2 Stage 1: Search process

We searched for relevant interventions representing models of support to improve the social circumstances of people with serious mental health conditions in these eight life domains in the following ways:

1. Screening papers from our published systematic review (Barnett et al., 2022) and relevant literature reviews identified during the systematic review search.
2. An updated search for relevant systematic reviews (up to February 2022).
3. Tracking references from relevant papers identified from the above methods.
4. An online consultation with identified domain specialists (academics, service providers and other stakeholders) in June 2022 (23 responses). An online form was circulated which asked them to share information about any published or unpublished interventions which met our inclusion criteria and any organisations or individuals who were involved in developing such interventions. Ethical approval was obtained for this stage of the process from the UCL Research Ethics Committee (REC Ref: 22343.001).
5. Direct email correspondence with additional domain specialists (8 responses).
6. Additional targeted literature searching to identify potential models for inclusion retrieved from the methods above, and to identify additional published evidence regarding included models.

We did not seek further evidence regarding models of support for finding paid employment (Individual Placement and Support) or establishing and maintaining tenancies for people without accommodation (Housing First), as these were found to have a strong evidence base in trials during our previous systematic review (Barnett et al., 2022). However, these models of support for getting paid work and for achieving and sustaining housing tenancies identified during our previous systematic review were included in our conceptual framework.

#### 2.2.1 Eligibility criteria

##### 2.2.1.1 Concept

We defined models of support meeting our inclusion criteria as interventions or approaches which were structured, described in writing, could be replicated, and involved some training or induction, or specified prior expertise of the support providers. Models were also required to be non-pharmacological and designed to improve social circumstances in any of the life domains of interest where this was an explicit, direct focus of the intervention. We also included models designed to improve more than one life domain, for example, through helping people access available services, groups or community resources.

###### Exclusions

In the social isolation and family relationships domains, following discussion with the stakeholder working group we excluded studies of models to improve individual social relationships, or the perceived quality of those relationships, which did not explicitly seek to help people with mental health conditions to develop or maintain desired social roles (for example, as partners, parents, carers or friends). Examples include programmes which focused on improving: i) individual relationship quality, including parent-child attachment and partner relationship quality; ii) family relationship quality, including reducing expressed emotion; and iii) experienced- or self-stigma.

###### Comparators

We included identified models regardless of whether they evaluated effectiveness against a comparator.

###### Outcomes

Where we reported on studies of evaluation of effectiveness, studies had to report at least one outcome specifically relating to the life domains listed in Table 1.

##### 2.2.1.2 Participants

Adults aged 18+ with a serious mental health condition, defined as a serious mental illness (psychosis or bipolar disorder) or any mental health condition of a seriousness and complexity that has involved or would typically require support from specialist mental health services. We included models designed specifically for people with serious mental health conditions, but also models targeted at the general population which were reported by stakeholders as being often used by, or evaluated with, people with serious mental health conditions.

###### Exclusions

We excluded models designed for people with intellectual disability, dementia or other organic mental disorder, neurodevelopmental disorder, acquired cognitive impairment, anti-social personality disorder, adjustment disorder, and substance use disorder (in the absence of any serious mental health condition, as defined above).

Additional information regarding the search process, including search strategies and additional operationalisation of inclusion criteria, is available in Appendix 1.

#### 2.2.2 Data extraction

Information from all included sources was extracted into a standardised extraction form in Microsoft Excel. Sources of information relating to interventions using the same model of support were extracted separately before being combined to ensure all relevant data were included for each model. Data were extracted by one reviewer, with another reviewer checking extraction and combining model information, if the same model was reported across multiple sources. The following information was extracted: intervention setting, target population, intended aims and outcomes, inclusion of lived experience in model development, procedures, frequency and modes of support within the model, and types of treatment provider involved. For evaluation studies, additional information regarding comparator and outcomes specific to the life domain(s) addressed or qualitative textual summaries of results were also extracted.

### 2.3 Stage 2: Development of the conceptual model

For each included model of support, the research team summarised the main aims (Appendix 2) and recorded available information regarding its characteristics and any supporting evidence regarding its effectiveness (see Appendix 3 and 4). A full list of all included sources of information which contributed to model descriptions is available in Appendix 5. This information was presented to our expert stakeholder group to help inform iterative development of an overarching conceptual framework.

#### 2.3.1 Expert stakeholder consultation workshops

In two workshops, a group of expert stakeholders, including people with lived experience of accessing mental health services, practitioners and academics, reviewed a provisional list of included models of support, with brief descriptions. Stakeholders were also invited to suggest additional models which had not been previously identified. Through discussion and with reference to previously published literature describing the models, we then developed a set of broad approaches for inclusion within our conceptual framework and distinguished different sources and types of support. Additional models of support, not previously retrieved through our scoping review, were also suggested by stakeholders at these workshops and considered for inclusion.

#### 2.3.2 Iterations and feedback

Following the two stakeholder workshops described above, the research team then mapped the models identified from the scoping review, expert stakeholder consultation and survey onto our conceptual framework, describing the sources and types of support provided. Revised versions of the framework were developed through internal working group meetings which included a smaller subset of stakeholders and lived experience researchers.

#### 2.3.3 Exploring the utility of the conceptual model

The team then created a summary table describing which sources and which types of support were used by models within each life domain, to test whether the categories within our framework were necessary and sufficient to capture the full range of models in each domain. Examples of types of support were identified and described to illustrate the range of support covered by each category.

The sources and types of support used by models, together with evidence of their effectiveness, were reviewed to identify broad approaches which showed the most promise in achieving improved social outcomes.

The final iteration of the conceptual framework was presented as an illustrative figure and approved at a last stakeholder working group meeting.

## 3. Results

After screening resources identified in our multi-component search strategy, we included 143 publications or written descriptions in our review, providing evidence on 80 different models. A further 12 models were identified from two other sources: responses to the expert consultation only (n=9) and models with high levels of evidence identified in our previous review (n=3), resulting in 92 models across our eight life domains. The full details of the searches and screening process is described in Figure 1. When reviewing the included models of support with stakeholders, it became apparent that there was considerable overlap between models included within the “Rights, inclusion and citizenship” domain and multi-domain models. We therefore merged these domains and reported them as “Inclusion and multi-domain models”.

**Figure 1:**
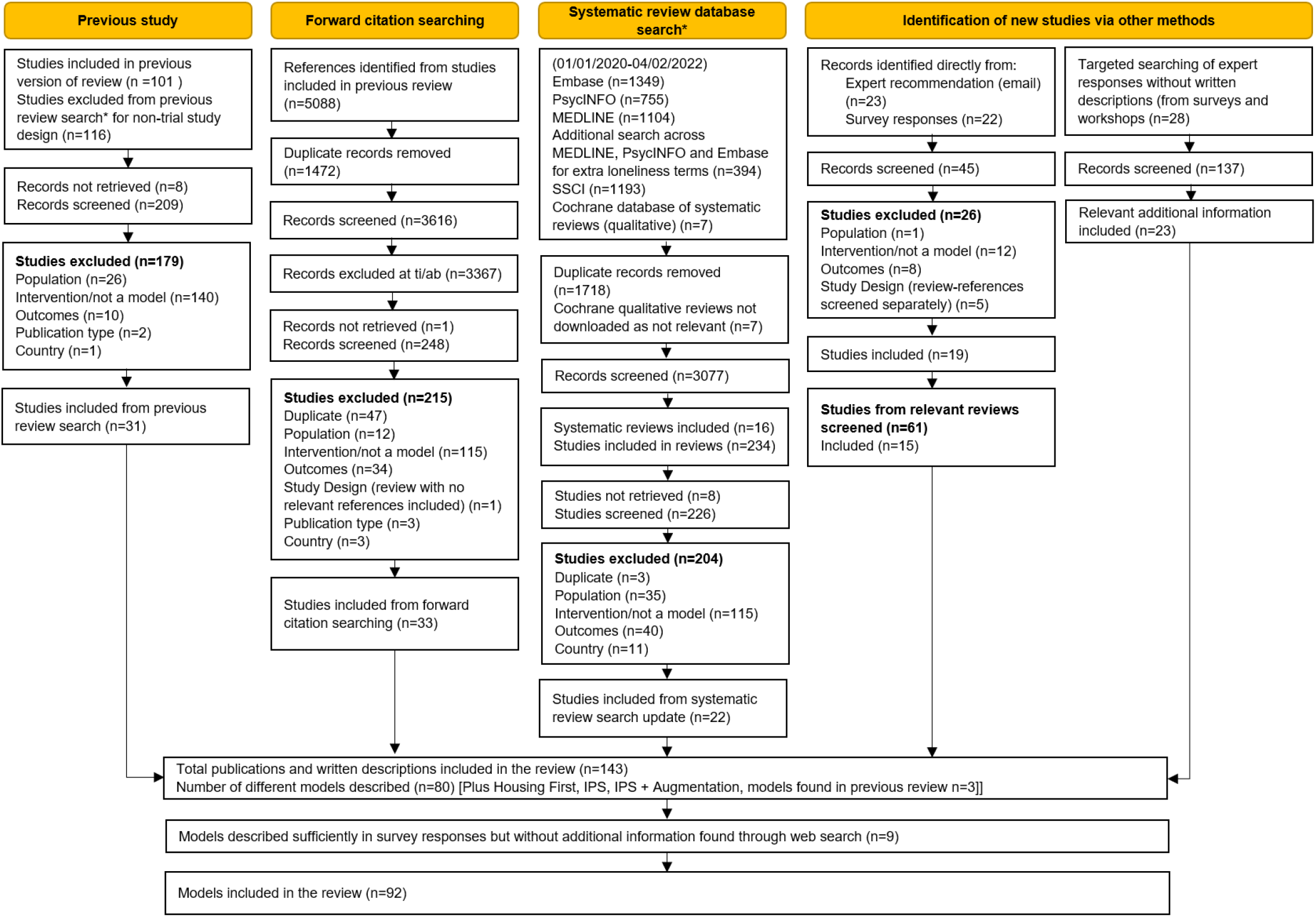
PRISMA diagram showing stages of searching for models and screening for eligibility

The number of models in each life domain were as follows: Housing and homelessness: 12; Money and basic needs: 6; Work and education: 11; Social isolation and connectedness: 27; Family relationships: 4; Victimisation and exploitation: 6; Offending: 6; Rights, inclusion & citizenship [amended to citizenship and multi-domain interventions]: 20

Through our initial review of included models, we distinguished five sources of support and five types of support. These are shown in Figure 2, along with proposed broad mechanisms by which they may improve the social circumstances of people living with serious mental health conditions. We used these overarching categories to describe and distinguish models within each domain.

**Figure 2:**
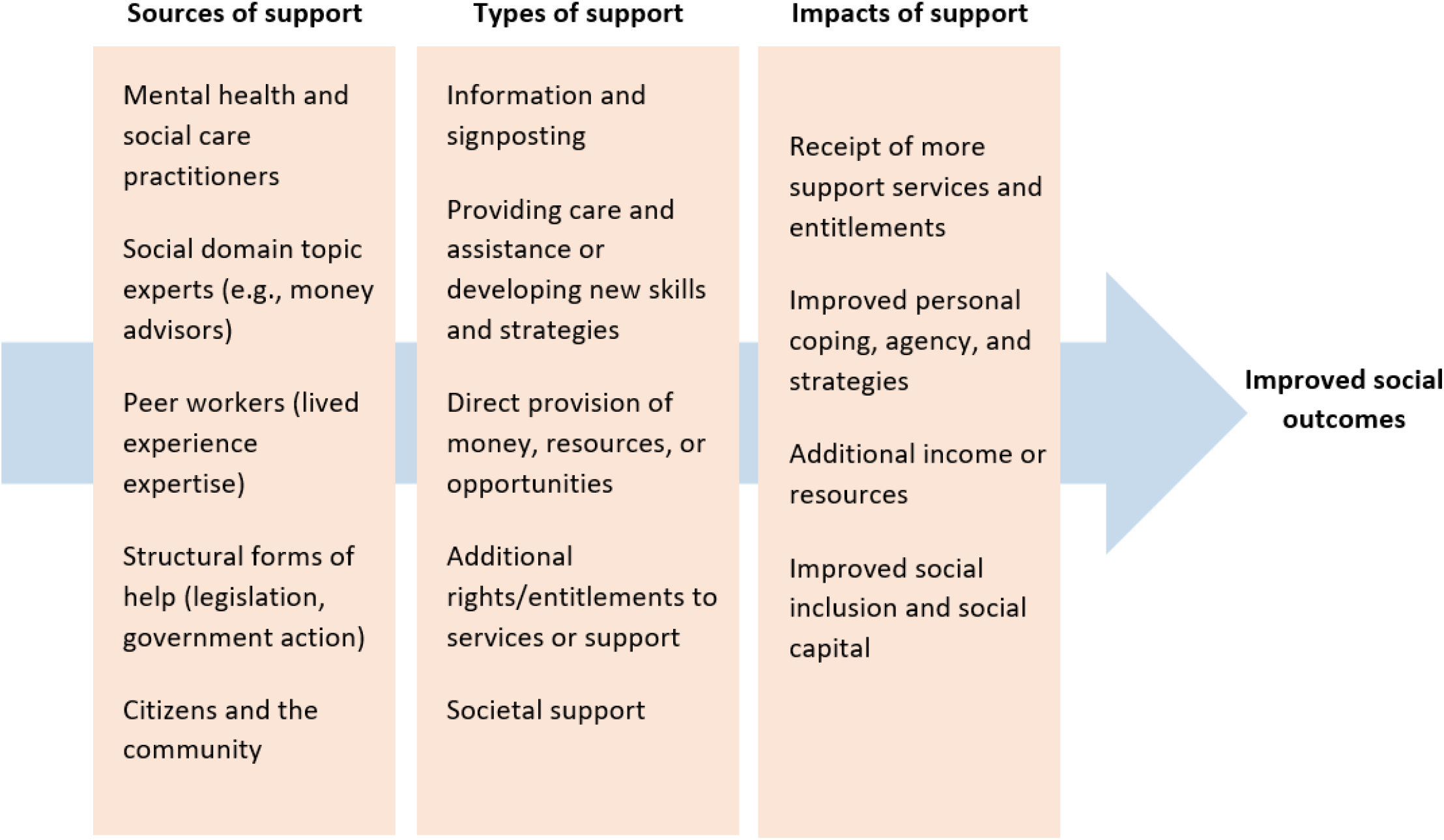
A conceptual model of the sources and types of support included in social interventions and their impacts

### 3.1 Sources of support

We distinguished support provided by:

- Mental health and social care practitioners: Staff working within health service, local authority or voluntary sector organisations with an overall remit to provide mental health or social care support, who may bring expertise or be trained to provide a specific model of support as part of their overall role.
- Domain specialists: Staff with expertise or a dedicated role to provide support with one of our included life domains (for example, an employment support worker or a money advisor). These staff may be embedded within mental health teams or work externally but do not have a broader remit to provide overall mental health or social care support.
- Peer workers: This includes any worker employed in a formal peer support role who uses lived experience and first-hand knowledge of mental health problems to help them to support people with serious mental health conditions (for example, a peer support worker, peer coach or peer mentor).
- Structural support: This category covers support involving or resulting from legislation or government action (for example, court-mandated treatment or support).
- Citizens and the community: This includes volunteers or members of the public providing structured support, with specific training, supervision or coordination from mental health and care provider organisations (for example, people providing a host family placement in their home). It also includes organisations external to the health and care system (for example, an employer using a structured model to provide job retention support to someone with a serious mental health condition).

### 3.2 Types of support

We distinguished five broad types of support, which may be provided or facilitated by more than one of the sources of support distinguished above. The broad types were:

- Information and signposting: Providing information about available guidance, resources or sources of support, or referral on to other helping agencies (for example, social prescribing services which involve providing information about or referral to existing groups and resources in the local community).
- Providing care and assistance, or developing new skills and strategies: Providing active support to help the person to manage a difficult social circumstance (for example, an employment support worker helping someone to complete an application form, develop their CV or practice interview skills).
- Direct provision of money, material resources or opportunities: Providing a tangible resource to meet a need or resolve a social problem (for example, schemes which set up and provide employment opportunities specifically for people with serious mental illness, or rent assistance to support someone to gain a housing tenancy).
- Increased rights or entitlements to services or support: Resulting from legislation or government action (for example, the “Mental Health Breathing Space” debt respite scheme, which offers people increased time and protection from debt enforcements during a mental health crisis).
- Societal support: Practical or emotional support provided outside of formal helping agencies.

Mental health and social care practitioners and specialist domain experts were the most commonly identified sources of support, being involved in models across all eight domains. Peer workers were also involved in a variety of interventions across six domains (housing, work and education, social isolation, victimisation, offending, and citizenship and multi-domain). Structural support from legislation or government action was also used across five domains (housing, money and basic needs, employment and education, victimisation, and offending). Interventions to improve outcomes in relation to work and education had the greatest variety of sources of support.

Both ‘information and signposting’ and ‘providing care and assistance, or developing new skills and strategies’ were found as types of support across all eight domains. ‘Direct provision of resources’ was also commonly reported, as this was used for all domains apart from victimisation. ‘Increased rights or entitlements’ was the least identified type of support, being used in interventions across three domains only (money and basic needs, work and education, and offending).

### 3.3 Example interventions in each domain

In this section, we provide one example of a model from each life domain, showing how it would be categorised using our framework for sources and types of support. The examples mentioned were chosen to allow for a variety of different sources and types of support to be described, rather than selected as examples of best practice. A fuller description of all the models of support we identified is available in Appendices 2 and 3 and information how they map onto our conceptual framework is provided in Appendix 6.

#### 3.3.1 Housing: Housing First

Housing First (Tsemberis, 1999) offers rent assistance to support housing choice in addition to intensive mental health support by **mental health and social care practitioners** and practical assistance. People are offered direct resources in the form of rent assistance to gain a housing tenancy at market rates and are given a degree of choice about type and location of housing; then high-intensity community mental health support, typically following an assertive community treatment model, and including practical support with housing-related needs, to help with mental health recovery and sustain independent housing. This model therefore involves two types of support: **providing care and assistance** and **direct provision of resources**.

#### 3.3.2 Money and debt: Mental health breathing space

Mental Health Breathing Space (Mental Health and Money Advice) is an intervention which aims to provide short-term respite from debt for people with mental health problems. It involves **structural help** in the form of a legal right to a break from creditors enforcing debts or non-payment charges so they can focus on mental health recovery. Support with debt can also be provided through **debt advice topic experts**. A variety of types of support are offered to service users. For example, service users who are identified as eligible for this intervention are **given information and signposted to sources of support**, and are **provided with care and assistance** via debt support and money advice. They also receive **increased rights and entitlements** as they benefit from short-term relief from their debt.

#### 3.3.3 Work and education: Social firms

Social firms (Social Firms UK) are social enterprises which specifically employ people who have a disability or are otherwise disadvantaged in the labour market, including those with severe mental health conditions. Social firms are not-for-profit agencies (**citizens and the community**) and are often funded through their own activities, although they do sometimes receive government funding (representing **structural support**). In this intervention, service users receive **direct provision of resources** and **societal support** from their employer.

#### 3.3.4 Social isolation: Mental health social prescribing

Mental health social prescribing (Dayson et al., 2020) adapts the social prescribing approach usually provided in primary care to offer more sustained and higher-intensity support for people with serious mental health conditions, supporting discharge from specialist community mental health care. People are offered linking support from a specialist social prescribing advisor (**a topic expert**) over a six-month period, plus access to **peer-provided befriending**. This intervention provides support by **giving information and signposting to resources** and **societal support**.

#### 3.3.5 Family roles: Thresholds Mothers’ Project

The Thresholds Mothers’ Project (Hanrahan et al., 2005) aims to provide parents with support to meet basic needs in order to maintain or retain custody of their children. It is a problem-solving approach to psychosocial rehabilitation and intensive case management. Practical problems in daily living are a focus of the model, and mothers are helped to meet their basic needs, stabilise their living arrangements, and begin addressing psychiatric symptoms. Sources of support are **mental health and social care practitioners** and **domain specialists**, as case managers help to secure entitlements, find independent apartments, and function as representative payees when needed. Case managers also assist with enrolling children in regular or special education. Types of support in this intervention can therefore be classified as **providing information and signposting** and the **direct provision of resources**.

#### 3.3.6 Victimisation and exploitation: The Victoria Intervention

The Victoria Intervention (Albers et al., 2021) aims to improve victimisation recovery for people with mental health problems through improving safe social participation. Sources of support includes **mental health and social care practitioners** as well as **domain specialists**, and **peer workers**, who work together to explore how victimisation may impact social participation and develop an action plan to support service users to safely participate in their community. This intervention provides support via **providing information and signposting, providing with care and assistance** and **social support**.

#### 3.3.7 Offending: Mental health courts

Mental health courts (Han, 2020) are an example of a type of **structural support** which aim to reduce reoffending. They operate by linking offenders with mental health conditions who would normally go to prison to long-term community-based mental health treatment. Mental health courts also feature ongoing judicial monitoring to ensure offenders are offered and adhere to community treatment plans. This intervention provides both **information and signposting to sources of support** and **increased rights or entitlements**.

#### 3.3.8 Citizenship & multi-domain: Clubhouses

Clubhouses (Pernice-Duca, 2008) are designed to help individuals transition from secondary mental health settings to community living and to address concerns of social isolation, readjustment to society, and community integration. They provide settings designed to foster social connections and community integration, including employment opportunities, housing support, case management and social programs for individuals living with schizophrenia and other psychiatric conditions. The Clubhouse model considers socialisation with fellow service users as a beneficial facilitator of social connection, provides employment opportunities, housing support and case management by **health and social care practitioners** who are trained in recovery practices. This approach therefore provides support via **providing care and assistance in developing new skills and strategies, direct provision of resources** and **social support**.

### 3.4 Identifying promising approaches

We know from our previous systematic review of trial evidence for social interventions (Barnett et al., 2022) that IPS employment support and Housing First have strong evidence bases. However, we identified few robust evaluations of other models of support through the current scoping review (see Appendix 5). We were therefore unable to draw inferences about the relative effectiveness of different broad approaches, although we note both IPS and Housing First were complex, multi-component programmes involving several of our types of support.

## 4. Discussion

### 4.1 Main findings

We have developed a conceptual framework which distinguishes five sources of support and five types of support, which are necessary and sufficient to describe 92 models of support we identified through a scoping review. Our paper demonstrates a range of broad approaches have been used in most life domains to try to improve social outcomes for people with serious mental health conditions. However, there is currently limited evidence as to which may be more or less effective in each domain and context. This lack of convincing evaluation of most models of support limits how far we can draw any conclusion about most promising approaches, however models of support such as IPS and Housing First are complex interventions involving more than one type of support from our framework, with the most robust current evidence base (Barnett et al., 2022, Killaspy et al., 2022).

As illustrated in Table 2, there is a range of different approaches service planners and providers can utilise to address social needs of people with serious mental health conditions. Different sources of support (community, domain specialists etc) can provide multiple types of support (information, resources etc). Some models of support which were identified primarily in one domain could also result in impacts across others, for example the breathing space debt scheme may also help prevent eviction, even though this is not its focus. Each model does not have to be prescriptive: within each model there are also opportunities to personalise the approach to suit the service and context, while utilising the same sources and broad types of support.

**Table 2:** Sources and types of support used in models identified for each life domain

| Domain | Number of models | *Sources of support* |  |  |  |  | *Types of support* |  |  |  |  |
| --- | --- | --- | --- | --- | --- | --- | --- | --- | --- | --- | --- |
|  |  | Mental health & social care practitioners | Domain specialists | Peer workers | Structural support | Citizens and the community | Information and signposting | Providing care and assistance | Direct resource provision | Increased rights or entitlements | Societal support |
| Housing and homelessness | N=12 | ✓ | ✓ | ✓ | ✓ | ✓ | ✓ | ✓ | ✓ |  | ✓ |
| Money and basic needs | N=6 | ✓ | ✓ |  | ✓ |  | ✓ | ✓ | ✓ | ✓ |  |
| Work and education | N=11 | ✓ | ✓ | ✓ | ✓ | ✓ | ✓ | ✓ | ✓ | ✓ | ✓ |
| Social isolation and connectedness | N=27 | ✓ | ✓ | ✓ |  | ✓ | ✓ | ✓ | ✓ |  | ✓ |
| Family relationships | N=4 | ✓ | ✓ |  |  |  | ✓ | ✓ | ✓ |  |  |
| Victimisation and exploitation | N=6 | ✓ | ✓ | ✓ | ✓ |  | ✓ | ✓ |  |  | ✓ |
| Offending | N=6 | ✓ | ✓ | ✓ | ✓ |  | ✓ | ✓ | ✓ | ✓ |  |
| Citizenship and multi-domain | N=20 | ✓ | ✓ | ✓ |  | ✓ | ✓ | ✓ | ✓ |  | ✓ |

Bespoke, innovative models which lack published description or evaluation may still be valuable in some contexts to meet local needs. However, it is likely that the current lack of evidence regarding the effectiveness of social interventions is also a barrier to receiving parity of funding with physical and psychological interventions. There is therefore a need to understand more about these models and about what forms of help work best, but without constraining them excessively and losing individualisation. Rigorous description of projects and service evaluation is desirable within a practice context, as even this could add to the current evidence base in many areas. We hope that our framework helps define broad approaches to guide further careful steps in research in this area.

### 4.2 Strengths

A particular strength of the conceptual framework development was that the research team included a large, multi-disciplinary group of stakeholders. This group of stakeholders included several people with lived experience of mental health problems and practitioners. The resulting conceptual framework went through several iterations following feedback from our lived experience working group, who were instrumental in ensuring the framework was designed in an accessible manner.

In developing the framework, we gathered data from a variety of sources in addition to searching the academic literature, including academics and topic experts, stakeholders working to deliver interventions for this population group, voluntary sector organisations, and policy makers. Therefore, we can be assured that our framework represents a wide variety of models of support being used in practice, not just those with published evidence.

Finally, we have tested the utility of our conceptual framework through mapping onto it 92 models of support, meaning it is not simply theoretical; we are confident the framework has practical utility to describe and distinguish different broad approaches.

### 4.3 Limitations

There are also some limitations which are important to consider. Firstly, we were unable to undertake an exhaustive scoping review of the literature due to the breadth of the topic, therefore we may have missed some interventions despite the multi-faceted search strategy. The stakeholder consultation and workshops also mainly involved experts based in the UK, meaning we may have missed some international models which are not described in the literature. There were also only a small number of models identified in some domains e.g. family relationships, meaning we were unable to test the framework on a broad range of models across all domains.

We only included models of support which have been developed for or evaluated with, or reported as frequently used by people with serious mental health conditions. We may therefore have excluded models for the general population or other clinical groups such as those with less serious mental health conditions which are also useful for people with more severe problems.

Our conceptual framework included theorised impacts of support proposed by our stakeholder group. However, we recognise these are not comprehensive and do not reflect all mechanisms or confirmed mechanisms of action for specific models. In addition, the limited extent of published evaluations meant we were not able to confidently infer the more or less promising approaches for each life domain, indicating a need for further research exploring the effectiveness of models of support.

Finally, whilst our framework distinguishes different broad approaches, the specific content of individual models within each source and type of support might vary considerably and leave room for flexibility and personalisation within the programme.

## 5. Conclusions

We developed a framework which distinguishes five sources and five types of support to meet the social needs of people with serious mental health conditions and used this framework to describe 92 models of support. This framework can be used in future primary research or systematic reviews to describe models of support and support evaluation of different broad approaches.

## Data Availability

All data produced in the present work are contained in the manuscript

## Lived Experience Commentary by Karen Machin and Beverley Chipp

This conceptual framework for social interventions may not only assist researchers to describe, compare and evaluate different approaches, but might also be useful in practice within health and social care, including for people accessing services. This framework has the potential to create clarity about the various approaches on offer, as well as highlight local gaps, and facilitate a more joined-up network of services between providers.

The paper was developed within a UK-funded project and, whilst people have universal needs, model applications may need adjustments for different cultural and socio-political systems. The framework could be developed through shared learning from an international audience including those with mental health conditions whose priorities should not be paternalistically assumed.

Personal happiness and sense of purpose also need to be addressed alongside the practicalities. In popular conception what matters for a good life has also shifted over the course of this project (Barrington-Leigh, 2022). For example, before Covid-19 we were slower to consider digital exclusion, which seems obvious to include now, but may be a lesser priority for people in nations where the gateway to services and information is less dependent upon internet access. Personal priorities may be influenced by international variations in income, comparative working conditions, housing situations and existence of a welfare state.

This framework is based upon published evidence, as well as the views and experiences of a range of people including those with lived and learned experience. Many approaches have not yet been thoroughly evaluated, but that does not mean they do not work. We also do not know which approaches work best for which people. However, as people with lived experience, we would argue for choice: that people needing help have agency and a range of options. What services are on offer, how to access them and, importantly, what we might expect from them, are often unclear. This framework, defining the range of potential offers, may support a shared language for voicing choices and improving lives.

Appendix 1 - Additional details on the search process

Appendix 2 - Models and descriptions

Appendix 3 - Model characteristics

Appendix 4 - Evaluation outcomes

Appendix 5 - References of Included sources

## Acknowledgements

We would like to thank TK, a member of our lived experience working group, for their help with this work.

