## Appendices for "Development of a conceptual framework to guide description and evaluation of social interventions for people with serious mental health conditions"

### Appendix 1: Additional details on the search process

#### Systematic review search strategy

Embase 2020-February 4^th^ 2022: 1349 records identified

PsycINFO 2020-February 4^th^ 2022: 755 records identified

MEDLINE 2020-February 4^th^ 2022: 1104 records identified

Additional search across MEDLINE, psycINFO and Embase for extra loneliness terms 2020-February 4^th^ 2022: 394 records identified

SSCI 2020- February 4^th^ 2022: 1193 records identified

Cochrane database of systematic reviews also searched for non-RCT (qualitative) reviews published in the last 2 years: 0/7 identified as relevant.

4795 records identified

1718 duplicates removed

3077 records screened

##### Example strategy: EMBASE

Embase <1974 to 2022 February 04>

1	systematic review/ or meta analysis/ or network meta-analysis/	443252

2	((systematic or structured or evidence or trials or studies) and (review or overview or look or examination or update* or summary)).ti.	243055

3	(0266-4623 or 1469-493X or 1366-5278 or 1530-440X or 2046-4053).is.	28564

4	(systematic review? or evidence report* or technology assessment?).jw.	35835

5	(meta-analys* or meta analys* or metaanalys* or meta synth* or meta-synth* or metasynth*).ti,ab,kw,hw.	370237

6	((systematic or meta) adj2 (analys* or review)).ti,kw. or ((systematic* or quantitativ* or methodologic*) adj5 (review* or overview*)).ti,ab,kw,sh. or (quantitativ* adj5 synthes*).ti,ab,kw,hw.	476890

7	exp "clinical trial (topic)"/ and review.ti,kw,pt.	149250

8	(integrative research review* or research integration).ti,ab,kw. or scoping review?.ti,kw. or (evidence adj3 review*).ti,ab,kw. or (narrative adj (review* or synthesis)).ti,ab,kw.	99533

9	review.pt. and (medline or medlars or embase or pubmed or scisearch or psychinfo or psycinfo or psychlit or psyclit or cinahl or electronic database* or bibliographic database* or computeri#ed database* or online database* or pooling or pooled or mantel haenszel or peto or dersimonian or der simonian or fixed effect or ((hand adj2 search*) or (manual* adj2 search*))).ti,ab,kw,hw.	186571

10	review.pt. and ((evidence based adj (medicine or practice)) or (outcome? adj (assessment or research)) or treatment outcome).hw.	231087

11	or/1-10	984646

12	mental disease/ or mental health/ or community mental health/ or psychopathy/ or *psychological well-being/	398040

13	(mental* adj2 (disorder* or health* or ill*)).ti,kw.	101768

14	anorexia nervosa/ or binge eating disorder/ or bulimia/ or eating disorder/ or automutilation/ or suicidal behavior/ or self poisoning/ or suicidal ideation/ or suicide/ or suicide attempt/ or mania/ or hypomania/ or manic psychosis/ or bipolar disorder/ or bipolar depression/ or bipolar i disorder/ or bipolar ii disorder/ or bipolar mania/ or cyclothymia/ or manic depressive psychosis/ or rapid cycling bipolar disorder/ or *depression/ or agitated depression/ or atypical depression/ or chronic depression/ or depressive psychosis/ or dysphoria/ or dysthymia/ or endogenous depression/ or involutional depression/ or late life depression/ or major depression/ or melancholia/ or minor depression/ or mourning syndrome/ or organic depression/ or perinatal depression/ or antenatal depression/ or postnatal depression/ or post-stroke depression/ or postoperative depression/ or premenstrual dysphoric disorder/ or reactive depression/ or recurrent brief depression/ or seasonal affective disorder/ or *subsyndromal depression/ or treatment resistant depression/ or puerperal psychosis/ or neurosis/ or affective neurosis/ or anxiety neurosis/ or dysthymia/ or hysteria/ or neurasthenia/ or psychasthenia/ or adjustment disorder/ or anxiety disorder/ or acute stress disorder/ or *distress syndrome/ or generalized anxiety disorder/ or panic/ or posttraumatic stress disorder/ or separation anxiety/ or mutism/ or selective mutism/ or obsessive compulsive disorder/ or compulsion/ or obsession/ or phobia/ or agoraphobia/ or claustrophobia/ or neophobia/ or social phobia/ or somatoform disorder/ or body dysmorphic disorder/ or conversion disorder/ or delusional pregnancy/ or hypochondriasis/ or masked depression/ or psychogenic pain/ or somatic delusion/ or malingering/ or somatization/ or mood disorder/ or affective neurosis/ or affective psychosis/ or blunted affect/ or major affective disorder/ or minor affective disorder/ or munchausen syndrome by proxy/ or munchausen syndrome/ or psychosexual disorder/ or kleptomania/ or trichotillomania/ or emotional disorder/ or dissociative disorder/ or depersonalization/ or dissociative amnesia/ or dissociative fugue/ or multiple personality/ or personality disorder/ or antisocial personality disorder/ or avoidant personality disorder/ or borderline state/ or catatonia/ or character disorder/ or compulsive personality disorder/ or dependent personality disorder/ or diogenes syndrome/ or histrionic personality disorder/ or narcissism/ or paranoid personality disorder/ or paranoia/ or passive aggressive personality disorder/ or schizoidism/ or schizotypal personality disorder/ or psychosis/ or acute psychosis/ or affective psychosis/ or brief psychotic disorder/ or delusion/ or depressive psychosis/ or endogenous psychosis/ or hallucination/ or manic psychosis/ or paranoid psychosis/ or schizophrenia/ or catatonic schizophrenia/ or hebephrenia/ or latent schizophrenia/ or negative syndrome/ or paranoid schizophrenia/ or positive syndrome/ or residual schizophrenia/ or schizophrenic reaction/ or simple schizophrenia/ or treatment-resistant schizophrenia/ or schizophrenia spectrum disorder/ or delusional disorder/ or schizophreniform disorder/ or schizoaffective psychosis/	946969

15	(acute stress or adjustment disorder* or ADNOS or affective disorder* or agoraphobi* or anorexia nervosa or anxiety or astheni* or attachment disorder* or BPD or binge eat* or binging or bipolar or body dysmorphi* or bulimi* or catatoni* or combat disorder* or compulsi* or conversion disorder* or cyclothymi* or delusion* or depersonali#ation or depressed or depression or depressive or dissociative disorder* or dyssomni* or dyspareunia* or dysphori* or dysthymi* or dystoni* or eating disorder* or EDNOS or emotional trauma or fear or health anxiety or hoarding or hyperactivity or hypochondri* or hysteri* or medically unexplained or malingering or mania or manic or MDD or mental* or mood? or munchausen or MUPS or mutism or neurastheni* or neurotic or neuros* or obsess* or panic or paranoi* or parasuicid* or perceptual disorder* or personality disorder* or phobi* or PND or ((post-trauma* or posttrauma*) adj stress*) or psychiatr* or psychogenic or psychopathol* or psychosomatic or psychotic or psychos* or PTSD or schizo* or (self adj (injur* or harm or mutilat*)) or (sexual dysfunction* adj3 psycho*) or social anxiety or somati* or somatoform or suicid* or trichotillomani* or stalking).ti,kw.	1136912

16	or/12-15	1622342

17	((child* or adolescen* or teen* or school or schools or youth?) not (childbirth or adult* or family or families or mother? or woman* or women* or female? or father? or men or mens or male? or relations or former)).ti.	1082398

18	(dementia not carer?).ti.	71522

19	prevalence.ti.	202577

20	16 not (17 or 18 or 19)	1467617

21	11 and 20	78882

22	(social prescribing or ((chang* or develop* or enhanc* or initiative? or intervention? or program* or mitigat* or address* or improv* or target*) adj3 (community or living or social) adj3 (condition? or circumstance?)) or ((communit* or social) adj (connect* or engagement? or link* or referral? or intervention? or wellbeing))).ti,ab,kw.	18944

23	"sense of belonging".ti,ab,kw.	1498

24	21 and (22 or 23)	282

25	exp *environmental planning/ or *city planning/ or *neighborhood/ or *social environment/ or psychosocial environment/	26254

26	*housing/ or *assisted living facility/ or *community living/ or *emergency shelter/ or homelessness/ or exp homeless person/	24941

27	*deinstitutionalization/ or halfway house/	2365

28	housing.ti. or (((chang* or develop* or enhanc* or initiative? or intervention? or program* or mitigat* or address* or improv* or target*) adj3 (housing or neighbo?rhood?)) or homeless* or ((housing adj (first or stability or instability)) or permanent housing) or ((housing adj (strateg* or polic* or project* or program* or quality)) or new* buil* or social housing*) or ((autonomous or assisted or sheltered or support*) adj3 (housing or accommodation or dwelling?)) or (((clubhouse or club house) adj model?) or ((autonomous or independent or assisted) adj living)) or ((independ* or assist* or support* or secur* or sustain* or maint*) adj3 (tenanc* or tenure?)) or ((halfway or satellite) adj (dwelling? or home? or house?)) or (neighbo?rhood? adj (characteristic* or intervention* or program*)) or ((environment* or housing or neighbo?rhood?) and infrastructure)).ti,ab,kw.	46979

29	21 and (25 or 26 or 27 or 28)	669

30	money/ or welfare/ or social welfare/ or *socioeconomics/	62549

31	household income/ or personal income/ or family income/ or *financial management/ or "salary and fringe benefit"/ or *pension/ or *salary/	76807

32	human needs/ or basic needs/ or personal needs/ or social needs/	3802

33	(((access* or improv* or manag* or supplement*) adj2 (cash or money or financ* or income? or savings)) or ((financial adj (autonomy or security or insecurity)) or loans or borrowing or budgeting or microcredit or microfinance or social fund*) or high poverty or ((address* or escap* or improv* or support* or target*) adj2 (depriv* or poor or poverty)) or (((food or fuel) adj poverty) or food bank?) or ((alleviat* or ease or manag* or prevent* or reduc* or stop*) adj2 (poverty or ((economic or financial) adj hardship?))) or ((alleviat* or eas* or manag* or prevent* or reduc* or relief or stop*) adj1 debt?) or (((basic or minimum) adj3 (wage? or income?)) or zero hours) or paid work or (family adj (income? or tax credit?)) or welfare benefit?).ti,ab,kw.	41600

34	(money or poverty or debt).ti.	12717

35	"out of poverty".ab.	161

36	"dealing with money".ab.	11

37	21 and (30 or 31 or 32 or 33 or 34 or 35 or 36)	506

38	*unemployment/ or *employment status/ or supported employment/ or sheltered workshop/ or *vocational rehabilitation/	10969

39	*absenteeism/ or presenteeism/ or *job security/ or *return to work/	9358

40	(return* adj2 work).ti. or (return* adj2 education).ti,ab,kw.	3575

41	(employment or employee? or unemploy* or (vocational and rehabilit*) or absenteeism or presenteeism or worker* or work disabilit* or (labo?r adj (force or market))).ti.	106021

42	21 and (38 or 39 or 40 or 41)	606

43	37 or 42	1082

44	*loneliness/ or *social isolation/ or social alienation/	10225

45	community involvement/ or *social support/ or *social network/ or psychosocial environment/ or *psychosocial rehabilitation/	35348

46	(loneliness or social inclusion or social participation or ((social* or communit* or peer) adj2 (network* or support*))).ti.	25894

47	(((subjective or objective) adj social isolation) or ((chang* or develop* or enhanc* or initiative? or intervention? or program* or mitigat* or address* or improv* or target*) adj2 (loneliness or ((social or community) adj (connect* or inclusion or isolation or network? or participation or relations*)))) or ((alleviat* or ease or manag* or mitigat* or prevent* or overcom* or reduc* or stop*) adj2 (isolation or exclusion)) or ((address* or enhanc* or improv* or increas* or promot* or target*) adj2 (inclusion or inclusivity)) or navigator? or ((address* or enhanc* or improv* or increas* or promot* or target*) adj3 (social* or communit*) adj (network? or support)) or social mobilit* or anti-stigma* or ((intervention? or alleviat* or prevent* or reduc* or stop*) adj2 stigma*)).ti,ab,kw.	27287

48	21 and (44 or 45 or 46 or 47)	949

49	international relations/ or family functioning/ or family conflict/ or *divorce/ or exp divorced person/ or separated parent/ or single parent/ or child custody/ or custodial care/	18807

50	(((family or families or intergenerat* or inter-generat*) adj (relation* or conflict?)) or ((sexual or intimate or partner? or marital) adj (relation* or conflict?)) or ((develop* or enhanc* or initiative? or intervention? or program* or address* or improv* or promot* or target*) adj2 relationship?) or ((carer? or partner or relationship? or marital) adj support*) or (child* adj2 (access or contact or custody or maintenance)) or ((care proceeding? or family court? or child removal or fostercare or foster care) and (parent* or mother? or father?)) or parent* outcome?).ti,ab,kw.	54998

51	parent* mental health.ti,ab,kw.	1072

52	(21 and (49 or 50)) or ((11 and 51) not (17 or 18 or 19))	777

53	(victimization or victimisation or revictimi#ation or crime victim? or revictimi or ((victim* or crime?) adj5 survivor*) or ((domestic or partner? or spouse?) adj3 (abus* or violen*)) or ((domestic or marital or partner? or spous*) adj3 (rape or sex* assault*)) or (intimate partner adj2 abus*) or coercive control or ((female? or women?) adj (refuge? or shelter?)) or exploitation or safe guarding or safeguarding or recidivism or ((crime? or criminal* or offend* or offence? or recidiv*) adj3 (initiative? or intervention? or program* or mitigat* or address* or rehabilitat*)) or ((crime? or criminal* or offend* or offence? or recidiv*) adj3 (diver* or prevent*)) or ((inmate? or prisoner? or convict? or felon?) adj3 (rehabilitat* or releas*)) or (community adj2 (reentry or re-entry or rehabilitat* or re-habilitat*))).ti,ab,kw.	70525

54	*crime victim/ or *offender/ or *recidivism/ or *prisoner/ or *prison/ or *community reintegration/	21874

55	exp *domestic violence/ or *human trafficking/ or *sex trafficking/	42084

56	21 and (53 or 54 or 55)	960

57	human rights/ or citizenship/ or civil rights/ or freedom/ or personal autonomy/ or reproductive rights/ or social justice/ or women's rights/	68907

58	digital divide/	378

59	rights.ti.	13448

60	(((citizen? or civil* or human or legal or social or voting) adj rights) or social justice or equal protection or social protection or ((public or social) adj polic*) or equity-focus* or ((social or community or neighbo?rhood?) adj3 (equit* or inequit* or inequalit* or dispar*)) or internet access or (digital adj (inclusion or exclusion or divide or disparit* or equit* or inequit* or inequalit*))).ti,ab,kw.	47383

61	21 and (57 or 58 or 59 or 60)	494

62	24 or 29 or 37 or 42 or 48 or 52 or 56 or 61	4594

63	(2020* or 2021* or 2022*).yr,dp,dc.	4353629

64	62 and 63	1349

#### Additional operationalisation of inclusion criteria

##### Model criteria

The review inclusion criteria for interventions – i.e. that models of support must be structured, described in writing, could be replicated, and involve some training or induction, or specified prior expertise of the support-providers – were operationalised to allow consistent screening. Models of support were included if they meet all of these criteria:

a) The model was named.

b) The model was explicitly described as seeking to address a social outcome in one of the eight life domains included in our review.

c) Written information was available from academic papers or other sources identified through the literature search or survey describing: i) who provides the support; ii) where support is provided; iii) the duration of support (e.g. whether a set or maximum number of sessions, or open-ended); iii) a description of the nature of the support sufficient to categorise it as psychological/therapeutic, practical or other).

d) Providers of the model of support required some training or specified prior expertise. Where this was unclear from retrieved papers or reports, the researchers sought further information, including through contacting provider organisations or developers if need be.

##### Population criteria

The review inclusion criteria for serious mental health conditions - a serious mental illness (Psychosis or bipolar 1); or any mental health condition of a seriousness and complexity that has involved or would typically require support from secondary mental health services – was operationalised with regard to the severity, chronicity and disability of the condition, to allow consistent screening. We included studies involving people with common mental disorders, anxiety, depression or unspecified mental health populations:

i) if they refer to severe, major, enduring, complex or treatment resistant conditions, or related terms

OR

ii) If participants were using any specialist mental health service other than in a primary care health service setting.

Studies of people with common mental disorders, anxiety, depression or unspecified mental populations were excluded if they did not meet either if these criteria. All otherwise eligible studies of people with psychosis or bipolar 1 were included. Studies of people with other included mental health populations, e.g. eating disorders or personality disorders, were included unless the studies were explicitly limited to people receiving support from primary care services, not secondary care or other specialist mental health services.

### Appendix 2: Models and descriptions

| Source | Domain(s) targeted | Name of Model | Description |
| --- | --- | --- | --- |
| Search | Housing | Board and care homes [floating outreach] | Provide housing for people with severe and long-term psychiatric disabilities |
| Expert | Housing | Critical time intervention | a 9-month case management intervention delivered in 3 phases- intensive support and assessment of existing resources, testing and adjusting systems of support identified and transfer of care where responsibility is handed over to the community resources that provide long-term support. |
| Search | Housing | Group homes [residential care] | Provide housing for people with severe and long-term psychiatric disabilities |
| Expert/previous review | Housing | Housing First | Programme involving placement in accommodation taking into account the persons choice, with rent assistance, plus access to practical and mental health support- used with homeless people with a serious mental health condition |
| Survey | Housing | Intensive supported accommodation [supported housing]* | Designed to support people with coexisting conditions who are rough sleeping or homeless. Street support provided and individuals encouraged to access supported accommodation that is less restrictive. The team also engages with current and potential professional and personal supporter networks to build their confidence, skills and understanding in supporting each person, designing bespoke strategies that tackle many years of disengagement, disenfranchisement and of being seldom heard and seldom effectively supported in services. Accepting that recovery is not a linear journey, people may be supported in a housing setting or at street level for times when they relapse. |
| Survey | Housing | Nottingham Supported accommodation [residential care]* | Nottinghamshire supported accommodation: intended to achieve a phased step-down to least supported housing option possible for people with a serious mental health condition. |
| Search | Multi-domain | Los Angeles' Homeless Opportunity Providing Employment (LA's HOPE) | This programme aimed to provide support to homeless mentally ill participants by providing supportive services and housing assistance, followed by support to gain employment through a variety of avenues |
| Search | Housing | Peer-enhanced case management programme | Peer assistants assist participants by providing support negotiating their new surroundings, locating needed services in their neighbourhoods, acting as mentors, and encouraging socialisation with other participants from approximately 1 month prior to their transition to assisted living |
| Survey | Housing | Rethink Registered Care Home, Salisbury (Herbert house) [residential care]* | Delivers a range of supported housing services through accommodation services that offer a safe environment in which people can recover and build confidence, helping them feel better equipped to live independently in the community. |
| Survey | Housing | Rethink Supported Housing-Staffordshire [supported housing]* | Delivers a range of supported housing services through accommodation services that offer a safe environment in which people can recover and build confidence, helping them feel better equipped to live independently in the community. |
| Search | Housing | Sheltered housing [supported housing] | A housing programme to support individuals with a serious mental health condition within the community in their rehabilitation, and to prevent unnecessary inpatient admissions |
| Search | Housing | Supported accommodation [supported housing] | Provide housing for people with severe and long-term psychiatric disabilities. Medium intensity support. Group homes where residents have a tenancy and typically have facilities to cook and clean for themselves. Maybe 24-hour support or less with staff on site |
| Survey | Money | Hertfordshire money advisors in mental health teams* | takes referrals directly from community-based mental health clinical staff. Money advisors, who are located in mental health settings including CMHTs and Crisis Assessment and Treatment Teams, work with patients to increase their incomes and reduce mental health crises by improving mental health and wellbeing. The service also aims to reduce demand on mental health and specialist staff so they can concentrate on providing clinical care. |
| Expert | Money | Mental health and money toolkit | a self-help toolkit and the and guidance for mental healthcare professionals, developed with DHSC funding, in helping people with mental health problems manage finances and money stresses |
| Expert | Money | Mental health crisis Breathing Space | The Mental Health Crisis ‘Breathing Space’ scheme gives people in problem debt and receiving treatment for a mental health crisis a ‘breathing space’ from the people they owe money to, so they can focus on their mental health recovery. They can also access debt support and money advice. |
| Search | Money | Payeeship/representative payees | Money management program where the money manager is not the clinician to avoid effects on therapeutic alliance. When patients are "incapable" of managing their funds in their "best interests" the Social Security Administration stipulates that a representative payee is required to receive and manage their disability payments. |
| Survey | Money | Sheffield money advisors in primary and secondary mental health teams* | This programme provides support for people in primary and community mental health services. Patients are assigned an adviser with mental health, debt and financial capability expertise, who then works with a client’s whole support structure to sustain a more financially secure future. Most of the work relates to debt and welfare benefits. |
| Survey | Money | South London IAPT money advice pilot* | Clients receiving high intensity services within IAPT are routinely asked about money worries. Where worries are identified, clients are directly referred to citizens advice for a follow up appointment with a money advisor. Support is then delivered concurrently and or separately with mental health therapy depending on client preference. |
| Workshop | Employment/Education | Access to Work programme | help to get or stay in work if you have a physical or mental health condition or disability, provided by the UK government. |
| Search | Employment/Education | BRIDGE supported education/IPS with education focus (slightly different focus compared to IPS with education focus but wonder if they can be combined?) | A supported education service for adults with psychiatric disabilities who require assistance to pursue post-secondary education, employment or both. Consists of sessions exploring qualification options, skills for study and stress management as well as mentoring. |
| Search | Employment/Education | Collaborative mental health care | To extend the availability of specialty mental health resources in primary care settings, enhance communication and promote continuity and follow-up care |
| Expert/previous review | Employment/Education | Individual Placement and Support (IPS) | Support from an employment specialist to find open market employment, then ongoing support for service user, including engaging with employers, to retain and manage the job |
| Search | Employment/Education | Individual Placement and Support with education focus [Vocational support with early intervention (VIBE)- Major 2010)] | The approach following principles to meet EITHER employment or education goals, providing supported education instead of supported employment for those patients whom this may be a more appropriate goal (e.g. first-episode samples) |
| Expert/previous review | Employment/Education | IPS + Augmentation | IPS + a range of psychological interventions (e.g. CBT, cognitive remediation therapy, social skills training) designed to remove barriers to managing and retaining paid employment |
| Search | Employment/Education | Mind@Work | The Mind at Work programme is made up of 9 modules to improve job tenure. It is designed to be delivered to people who are already employed, either on site or online. |
| Expert | Employment/Education | Peer-delivered self-management intervention | The CORE study involves a specialised workstream in which a peer-delivered self-management intervention to bridge the gap between crisis and continuing care is provided. Peer support workers are provided with training and employed within the NHS to support the individual to identify facilitators for their recovery goals. Peer support workers are also provided with supervision from their NHS crisis team. |
| Search, Web | Employment/Education | Social firms | Types of social enterprise with a defining criterion of supporting and empowering disadvantaged people by drawing at least 25% of their workforce from groups facing barriers to mainstream employment |
| Survey | Employment/Education | The MENTOR programme | A programme that aims to intervene early and help people experiencing mental health conditions to stay in the workplace and get support to reduce the risk of them having to go off on longer term sick leave. The support offered is not therapy, and is instead support to improve workplace functioning, wellbeing and productivity, thereby improving people’s mental health |
| Expert | Employment/Education | User Employment Programme | The User Employment Programme aims to increase employment in mental health services for people who have mental health problems themselves (supported employment plus a Charter for the Employment of People who have Experienced Mental Health Problems that is designed to decrease employment discrimination) |
| Survey | Social isolation/inclusion | Arts based groups | A choir and a creative writing group for people referred from community mental health teams |
| Expert | Social isolation/inclusion | Community Navigator | 6-months, 10-sessions and group support from a social connections coach to help develop social relationships |
| Search | Social isolation/inclusion | Compeer | Compeer matches adult volunteers from the community in intentional friendships (called ‘matches’) with people in treatment for serious mental health conditions |
| Expert | Social isolation/inclusion | Connecting People Intervention | Connecting people is a model to enhance the care of service users in which the care provider and service user work together to help enhance the service users’ social networks |
| Search | Social isolation/inclusion | Consumer providers | Consumer providers offer a form of mutual support with a recovery orientated focus |
| Search | Social isolation/inclusion | Creative workshops | Creative workshops for people with a serious mental health condition and their carers, which are held in a museum setting to facilitate non-clinical improvements such as well-being or social connectivity |
| Search | Social isolation/inclusion | Education groups | 20 weeks of education groups covering topics such as relating to other people, problem solving and assertiveness, and more general illness related topics such as causes of schizophrenia, managing symptoms and signs of relapse, with the hypothesis that being provided with information about their condition improves social functioning. |
| Expert | Social isolation/inclusion | Expand your world | A series of workshops developed with a participatory approach to help service users identify social, vocational and health activities which are useful and enjoyable to them |
| Workshop | Social isolation/inclusion | Focus, Act, Connect Every-day (FACE) | a group of people with a history of mental health difficulties, trauma, substance misuse, homelessness and incarceration who support each other to engage with the community with various activities |
| Search | Social isolation/inclusion | Groups 4 Health | Groups 4 Health is a 5-module programme which targets the development and maintenance of social group relationships to treat psychological distress arising from social isolation. |
| Search | Social isolation/inclusion | Guided peer support | Minimally guided peer support to offer structure, continuity and sense of security without actively interfering with the group process. |
| Search | Social isolation/inclusion | Hearing Voices Network support groups | Hearing voices network support groups allow people who hear voices to come together and access peer support |
| Search | Social isolation/inclusion | Occupation and Social Skills Training | This programme offers sessions aimed at improving occupational and social skills with personalised goals. Themes included preparation for community living, practicing basic conversation skills, identifying desired activity goals and finding strategies to achieve them. Problem solving strategies to deal with daily living challenges are also taught. |
| Search | Social isolation/inclusion | Participatory Video | Participatory video (PV) is a group intervention where marginalised people get together to produce educational documentary-type videos about their lives and associated issues. |
| Search | Social isolation/inclusion | Peer Specialists | Peer specialists use their experiences as the basis of the service intervention to help service users with their recovery. The current model uses a mixture of peer specialists delivering structured recovery treatment sessions and unstructured 'relationship-based' sessions. |
| Search | Social isolation/inclusion | Project Connect | Supports people with mental illness to create and sustain connections in their communities based on their interests and talents and in way that they define most helpful to them. Community connections are facilitated by community partners who bridge the divide between the mental health community and the general community. |
| Search | Social isolation/inclusion | Recovery narrative photovoice | Photovoice is a community-based participatory action approach where participants take photos and write narratives to describe the images. |
| Search | Social isolation/inclusion | Role Development programme | Role development programme is an intervention where staff work collaboratively with participants to identify and develop the participant's social roles |
| Search | Social isolation/inclusion | Scatter -site & Congregate housing | Independent scatter-site housing included only housing categorised as “supported housing", whilst congregate housing is different residences all with 24 hour staff support on site |
| Search | Social isolation/inclusion | Social cognition and interaction training + peer mentoring | A manualized group-based intervention which targets impairments in social cognition alongside social mentoring services which support in vivo efforts to take practical steps toward achieving personally meaningful goals. |
| Search | Social isolation/inclusion | Social network intervention | Suggestions for areas of interest for social activity participation for patients which is outside of the community mental health centre resources, followed by 3-6 months of support to help integrate the patient in the activity by either a member or staff or natural facilitators such as volunteers or family members. |
| Search | Social isolation/inclusion | Social Prescribing | link workers identify social and psychological needs of patient and help them to access non-clinical or community activities/groups that can help with self-care and long-term psychological and social difficulties. |
| Search | Social isolation/inclusion | Social recreation programme | This social recreation component of a community mental health programme offers activities to meet individual and social/environmental goals, such as skills training and social network building |
| Expert | Social isolation/inclusion | Structured social coaching intervention | A social coaching model in which a coach works with a service user to identify opportunities for socialisation and support the service user to take part in social activities |
| Expert | Social isolation/inclusion | Supported socialisation | This supported socialisation model involves pairing a participant with a volunteer befriender (who may or may not have lived experience of mental illness) and a stipend for them to take part in social activities |
| Search | Social isolation/inclusion | TREE (Toward recovery, empowerment and experiential expertise) recovery programme | supports persons with severe mental illness to exchange experiences and offer mutual support, develop and share knowledge, and promote user-led change within mental health care organisations. |
| Search | Social isolation/inclusion | Volunteer befriending programmes | Befriending models involve people without mental illness volunteering to spend time with someone who does. They generally involve supervision by a third party. These have been tested on a one-to-one basis and in a group format (one person befriends up to three people with mental health conditions) |
| Expert | Family | Family support services (Parents, advocacy, coordination, education; PACE) | Supportive and unconditional relationships developed with families and services such as case management parenting skills training, child development education and individual therapy provided. 24-hour on call support and emergency assistance, monthly social support group for mothers, tenant-landlord mediation, crisis planning, financial assistance and transitional planning after hospitalisation are also provided, with most services provided within the home |
| Expert | Family | Integrated family treatment | A family specialist clinician provides home based services to parents and children while they participate in mental health services- e.g. engagement into treatment, linkage to environmental supports, educations about child development, parenting skills training, modelling and coaching. |
| Expert | Family | Mothers and children’s project | Program designed to supplement other mental health care which involves a visit by the program director, followed by weekly group meetings that focus on developmental education and role modelling. |
| Expert | Family | Thresholds mothers project | A problem-solving approach to psychosocial rehabilitation and intensive case management- practical problems in daily living are a focus of the model, and mothers are helped to meet their basic needs, stabilize their living arrangements, and begin addressing psychiatric symptoms. Case managers help to secure entitlements, find independent apartments, and function as representative payees when needed. Care managers also assist with enrolling children in regular or special education. |
| Workshop | Victimisation | BRAVE | Domestic violence training for clinicians in community mental health teams, training for domestic violence advocacy professionals, and direct referral pathways between CMH teams and domestic violence advocacy services for service users who have experienced domestic violence. This model was based on the LARA intervention and adapted for a Dutch context. |
| Workshop | Victimisation | Health Pathfinder project | Health Pathfinder is a multilevel system change intervention to transform the health response to domestic violence and abuse, including training professionals, co-locating domestic violence and abuse services in clinical settings, implementing new interventions, supporting domestic violence and abuse coordinators, undertaking needs assessments, enhancing data collection strategies, and reviewing clinical policies relating to domestic violence and abuse. |
| Workshop | Victimisation | LARA (Linking Abuse and Recovery through Advocacy) | Domestic violence training for clinicians in community mental health teams, training for domestic violence advocacy professionals as well as referral to domestic violence advocacy for service users. |
| Workshop | Victimisation | LINKS project | a pilot of two specialist domestic abuse independent domestic violence advocate-educators working with community mental health teams to provide domestic violence training to staff, give ongoing support to professionals, receive referrals, and provide a proactive and culturally sensitive support service for female and male inpatient psychiatric patients over 16 who were experiencing domestic abuse. |
| Expert/previous review | Victimisation | Self-wise, Other-wise, Streetwise (SOS) training | A six-week group training focused on enhancing emotion regulation skills, conflict resolution skills and street skills. |
| Search | Victimisation | The Victoria intervention | Explores the victimisation experience with participants and then develops action plan to promote safe participation in the community which reduces the chance of revictimization |
| Search | Offending | Forensic peer support service | Forensic peer support services are where a peer supporter who has experience of the MH service use assist forensic service users (can be in a variety of roles) to help model recovery and help connect with their community and any re-entry difficulties e.g. accessing services when out of prison |
| Workshop | Offending | Liaison and diversion schemes | Liaison and diversion schemes are a model used to divert people with certain needs or vulnerabilities away from the criminal justice system and refer them referred to appropriate interventions or treatment in the healthcare system. (unlike mental health courts, no ongoing supervisory role over offenders) |
| Search | Offending | Mental Health Court | Mental health courts link offenders with mental illness who would normally go to prison to long-term community-based mental health treatment. They have ongoing judicial monitoring to ensure offenders adhere to community treatment plans |
| Search | Offending | Serious and persistent mental illness (SPMI) Release Planning service | a pre-release transition service that attempts to connect offenders with needed services in the community following release e.g. housing, vocational, chemical dependency, psychiatric, disability, medical, medication, and transport needs |
| Search | Offending | Thinking for a Change (T4C) | A structured programme of 25 sessions delivered over 3 months that covers social skills training, cognitive restructuring and problem-solving modules |
| Search | Offending | RESET | The RESET intervention supports prisoners with mental health needs for 12 weeks after release to coordinate their transition into the community and obtaining secure housing |
| Search | Multi-domain | Clubhouses | Clubhouses have been implemented to transition individuals from hospital to community living and to address concerns of social isolation, readjustment to society, and community integration. They provide settings designed to foster social connections and community integration including employment opportunities, housing support, case management and social programs for individuals living with schizophrenia and other psychiatric conditions and are often credited for the creation of social connections that participants would not have otherwise formed |
| Survey | Multi-domain | Community-Enhanced Social Prescribing | Community-enhanced social prescribing (CESP) is a new model of social prescribing combining community engagement, organisational change and individual-level practice which aims to improve both community and individual wellbeing by recognising that individuals enrich the health of communities by developing opportunities for more active engagement with them. |
| Search | Multi-domain | EMILIA | Training related to mental health and social inclusion, personal development and planning, employment and recovery, as well as opportunities for unpaid and paid activities mainly within the demonstration sites themselves. |
| Expert | Multi-domain | Horyzons Project | Digital platform merging: peer-to-peer social networking; theory-driven and evidence-informed therapeutic interventions targeting social functioning, vocational recovery and relapse prevention; expert clinician and vocational support; and peer support and moderation |
| Survey, Web | Multi-domain | Hospital discharge service | The hospital discharge and crisis support service involves our support staff working alongside the crisis team or on the mental health wards, providing housing, benefit and claims, life skills, and community integration support for those identified as needing it, from the point of contact through to discharge within the community. |
| Search | Multi-domain | Intensive Psychiatric Rehabilitation (IPR) based on Choose-get-keep (CGK) | Helps people set their own individual rehabilitation goals, in terms of housing and employment and then supports them through regular meetings to achieve these goals |
| Search | Multi-domain | Intentional recovery communities | Recovery communities aim to improve the psycho-social needs of people who experience vulnerabilities and have a serious mental health condition. They can be residential or day centres and are underpinned by a recovery philosophy, through peer support from other service users and professional staff. |
| Workshop | Multi-domain | Look Ahead's mental health pathways | Look Ahead is a provider of housing and support services, working in integrated care systems in the UK. It integrates mental health care, social care and housing support, enabling individuals to receive tailored treatment, interventions and care closer to home and within the least institutionalised environment. |
| Expert | Multi-domain | Mental health day centres | Local centres funded by social services which offer patients a "drop in" facility, cheap food, and therapeutic groups and member's meetings |
| Survey | Multi-domain | Mental health inclusion service* | Wiltshire mental health inclusion service: one-to-one support with an inclusion coach to identify hobbies and interests as well as barriers to these |
| Survey, Web | Multi-domain | Mental Health Navigation | A model of support for people with mental illness who present at primary care with unmet non-clinical needs. This model builds on the well-established social prescribing model, offering a dedicated capacity to assist and empower people to access the right support, to manage a range of needs and social distresses that can affect an individual’s mental health and in doing so alleviates pressure on healthcare systems. |
| Search | Multi-domain | Peer-delivered centres for independent living | Embraces advocacy, peer support, consumer control and the removal of external barriers. Unmet needs where the patient would like more support assessed by a trained peer specialist (e.g. housing, activities, employment, education), guidance and advocacy provided until need is met. |
| Survey | Multi-domain | Psychologically Informed Environments | PIEs combine a therapeutic setting with housing, and try to meet the fundamental needs of hostel residents by providing psychological safety and rebuilding damaged attachment relationships through the provision of a professional home and family |
| Search, workshop | Multi-domain | Recovery Colleges | Recovery Colleges take an educational rather than a clinical or rehabilitation approach to improving mental health- they provide educational platforms to help self-directed recovery and provide learning opportunities for people with mental health problems. They also include programmes designed to contribute towards recovery and wellbeing, put people back in control of their lives, increase confidence and skills and provide support for accessing further opportunities. Training is offered by both professionals and individuals with lived experience, and courses promote choice and control, peer support and promote participation in the local community. As far as possible the distinction between service users and professionals is avoided and there is an emphasis on co-production, co-delivery and co-participation in the learning. |
| Survey | Multi-domain | Rethink mental illness community services: Brent mental health service* | Brent mental health service: provides support to adults with mental health needs aged 18+, within the Brent borough alongside statutory services. The service has 4 main aspects of delivery which are Befriending service utilising volunteers; Peer Navigators Support; Substance Misuse Support delivered via Change, Grow, Live (CGL) staff; Mental Health and Recovery Workshops. |
| Workshop | Multi-domain | Shared Lives | Providing up to two years of support for adults transitioning from care, matching people with a Shared Lives carer who supports them to move in and share family and community life. |
| Survey, Web | Multi-domain | Social cafes | Social cafes are free groups for people to go and meet others, socialise, and reduce isolation. They are usually held in community settings. Opportunities are also sometimes provided to service users to challenge their problems and gradually prepare for a job by taking part in daily work in the café, and access to computers to reduce digital exclusion is also provided |
| Workshop | Multi-domain | Social Welfare Legal services in health settings | provide specialist legal advice and assistance, within a healthcare setting; for example, issues relating to housing and homelessness, welfare benefits, debt, employment and family issues, among others, which can have an impact on physical and mental health and wellbeing. |
| Survey, Web | Multi-domain | STAR | Service users are able to access training and employment, and improve their physical and mental health. Support is also provided by the charity Shelter. |
| Search, Web | Multi-domain | The Citizens/Citizenship Project | The Citizenship Project is a 5-month programme involving formal and informal contacts with Peer Mentors and staff, group classes on social participation and community integration, projects to foster gained valued social roles and peer support. The model theorizes that greater community integration will reduce offending and drug use. |
| ** : Information for models gained only from survey responses. No additional information found through targeted searching* | | | |

### Appendix 3: Model characteristics

| Name of Model | Country model is described in (number of publications) | Aims/purpose of the model | Procedures/activities/processes in the model | Model details |
| --- | --- | --- | --- | --- |
| *Housing models* | | | | |
| Board and care homes [floating outreach] | Canada | To offer housing to people with severe and long-term psychiatric disabilities, with the exception of individuals who are violent or who have a recent history of drug or alcohol abuse | The Board and care homes (BCH) provide accommodation, meals, and, in some cases, recreation. The BCH do not have explicit rehabilitation goals or methods. Rather, they aim to provide comfort and support. If residents wish, they can avail themselves of support from external mental health workers, who could help them to improve their life skills and access social-recreational or employment opportunities in the community. | **Setting:** Non-profit housing organisations  **Modes and format:** F2F  **Frequency and intensity:** NR  **Types of treatment provider involved:** Housing coordinators and directors |
| Critical time intervention | USA | To gradually pass responsibility to community sources for providing ongoing support for people with severe mental illness, leading to a durable reduction in the risk of future homelessness. | While living in the transitional residence, all participants received basic discharge planning services and access to psychiatric treatment. After discharge, participants in both conditions received a range of “usual” community-based services depending on the individual’s needs, preferences and living situation. These services usually included various types of case management and clinical treatment. In addition to the services noted above, participants randomly assigned to the experimental condition received nine months of CTI following discharge from the transitional residence. In brief, it is a nine-month case management intervention delivered in three phases, each of which lasts approximately three months. Phase one--transition to the community--focuses on providing intensive support and assessing the resources that exist for the transition of care to community providers. Ideally, the CTI worker will have already begun to engage the client in a working relationship before he or she moves into the community. This is important because the worker will build on this relationship to effectively support the client following discharge from the institution. The CTI worker generally makes detailed arrangements in only the handful of areas seen as most critical for community survival of that individual. Phase two—try out-- is devoted to testing and adjusting the systems of support that were developed during phase one. By now, community providers will have assumed primary responsibility for delivering support and services, and the CTI worker can focus on assessing the degree to which this support system is functioning as planned. In this phase, the worker will intervene only when modification in the system is needed or when a crisis occurs. Phase three—transfer of care-- focuses on completing the transfer of responsibility to community resources that will provide long-term support. One way in which CTI differs from services typically available during transitional periods is that the transfer of care process is not abrupt; instead, it represents the culmination of work occurring over the full nine months. | **Setting:** Community  **Modes and format: F2F, individual**  **Frequency and intensity:** Nine-month intervention delivered in 3 phases where each one lasts approximately 3 months.  **Types of treatment provider involved:** Delivered by 3 workers trained by several of the model developers. 2 were bachelors-level employees of the New York State Office of Mental Health reassigned to this project from their regular duties and the third worker was a more experienced worker who had delivered CTI in an earlier trial. Weekly supervision was carried out by clinically trained staff experienced in the model. |
| Group homes [residential care] | Canada | To provide stable housing for people with severe and long-term psychiatric disabilities, reduce symptoms, and promote community integration, independence, and well-being. | Group homes provide high levels of support to people early in their recovery process before they "graduate" to lower support from supported accommodation. | **Setting:** Non-profit housing organisations  **Modes and format: F2F**  **Frequency and intensity: NR**  **Types of treatment provider involved:** Housing coordinators and directors |
| Housing First | USA (1), Canada (2), France (1) | To create a recovery-oriented culture for homeless people with severe mental illness that puts participant/tenant choice at the centre of all its considerations with respect to the provision of housing and support services. | Participants contribute a percentage (usually 30%) of their income toward rent, and subsidies cover the difference. Housing units consisted mostly of private-market scattered-site units. Clients are assisted to choose among available units and furnish and move into them. Study participants have to agree to observe the terms of their lease but there are not any prerequisites for psychiatric treatment or sobriety. They are then offered through multidisciplinary ACT services 24-hour support. Participants can choose what support to make use of (e.g. they can refuse clinical services altogether). | **Setting:** Community  **Modes and format: F2F individual**  **Frequency and intensity:** At least one weekly visit by support staff at a time suitable to the service user.  **Types of treatment provider involved:** Multidisciplinary mental health teams |
| Intensive supported accommodation [supported housing] † | UK | To support people with coexisting mental health and substance use problems conditions who are rough sleeping or homeless. | A specialist dual diagnosis team provides street support to individuals, encouraging them to access supported accommodation that is less restrictive. The team also engages with current and potential professional and personal supporter networks to build their confidence, skills and understanding in supporting each person, designing bespoke strategies that tackle many years of disengagement, disenfranchisement and of being seldom heard and seldom effectively supported in services. Accepting that recovery is not a linear journey, people may be supported in a housing setting or at street level for times when they relapse. | **Setting:** Community  **Modes and format:** NR  **Frequency and intensity:** NR  **Types of treatment provider involved:** NR |
| Los Angeles' Homeless Opportunity Providing Employment (LA's HOPE) | USA | To support mentally ill homeless to access housing and employment | Candidates were referred on to the Department of Mental Health to determine eligibility, then once deemed eligible, were enrolled onto one of three mental health programmes that were specially state funded, which provided supportive services and housing support to those with serious mental illness who were homeless/at immediate risk of homelessness- these participants received substantially more housing support that other participants in these programmes. Participants were then helped to apply for a shelter care plus certificate and to locate suitable housing, and provided with temporary housing in the meantime. Finally, participants were supported to gain employment, through either their own case managers connections and support, through the services' employment specialist (who could provide training, uniform, and employment preparedness training etc.), or two nearby workforce development centres, with coordination between these teams. | **Setting:** community mental health centres (outpatient)  **Modes and format:** Individual  **Frequency and intensity:** NR  **Types of treatment provider involved:** case workers, community mental health team staff, employment specialists |
| Nottingham Supported accommodation [residential care] † | UK | NR | In Nottingham(shire) supported accommodation includes the first step down from inpatient psychiatric stays, people stay at the accommodation under licence and access intensive personalised support over 6-24 months from a 24/7 on-site team. The model is trauma and psychologically informed and is delivered by experienced highly-skilled mental health recovery workers with as many of these as possible being people with lived experience. They apply strengths-based assessment and support approaches to reinforce and build skills and confidence to live as independently as possible in the community. This includes becoming registered with primary healthcare providers and improving physical health, being registered to vote, accessing employment and education, (re)building relationships, optimising income, building budgeting skills, understanding, and sustaining a tenancy. The outcome is to support move-on to supported living or general housing options with personalised transition plans and informed ongoing support plans designed to further and sustain recovery and self-determination outcomes. | **Setting:** Community  **Modes and format:** NR  **Frequency and intensity:** NR  **Types of treatment provider involved:** NR |
| Peer-enhanced case management programme | USA | To help facilitate homeless mentally ill veterans’ transition to independent community living | The goal of the intervention was for PAs to assist participants by providing support negotiating their new surroundings, locating needed services in their neighbourhoods, acting as mentors, and encouraging socialization with other participants in the program as well as through self-help groups. PAs met with participants starting approximately 1 month prior to the homeless veteran’s anticipated transition to community living and for approximately 12 months thereafter. Peers met with participants individually, in groups, and by telephone. Individual and group contacts usually took place in community settings such as coffee shops or parks. Each PA was expected to work with a maximum of seven individuals but due to slower than anticipated recruitment, peers worked with up to four individuals at one time. PAs were paid $7 per hour for up to 20 h per week. Peer advisors met weekly as a group for 12 weeks for training in relevant issues, including role of the PA, communication skills, relationship building and empathic listening. Training was based on the Handbook for Communication and Problem-Solving Skills Training (Bedell & Lenox, 1996). A psychologist with several years’ experience implementing peer-assisted case management programs led the group. After the initial training, PAs received weekly individual supervision with a licensed social worker. | **Setting:** Community drop-in centre for veterans  **Modes and format:** F2F individual and group, phone individual  **Frequency and intensity:** Peer advisors met weekly as a group for 12 weeks for training, then met each veteran for 13 months (each PA worked up to 20h/wk)  **Types of treatment provider involved:** Peer support workers, psychologist, social workers |
| Rethink Registered Care Home, Salisbury (Herbert house) [residential care] † | UK | To provide supported housing services to offer a safe environment in which people can recover and build confidence, helping them feel better equipped to live independently in the community. | Herbert House is a registered care home (CQC) providing accommodation and personal care to 13 people with support needs related to their mental health. The service can support up to 15 people, with the other 2 beds allocated as “crisis beds” for short-term (1-2 week) stays. | **Setting:** Community  **Modes and format:** NR  **Frequency and intensity:** NR  **Types of treatment provider involved:** NR |
| Rethink Supported Housing-Staffordshire [supported housing] † | UK | To provide supported housing services to offer a safe environment in which people can recover and build confidence, helping them feel better equipped to live independently in the community. | Staffordshire Supported Housing provides high quality accommodation and low-level housing management support for people affected by mental illness aged 18 years and above, who are at risk of not accessing or sustaining a home because of their mental health, vulnerability and/or a history of housing related problems. The service has 57 accommodation units, located across four local authority areas and requires minimum stay of 6 months. There is also an out-of-hours housing related on-call service. The data broadly indicates various positive outcomes for service users. | **Setting:** Community  **Modes and format:** NR  **Frequency and intensity:** NR  **Types of treatment provider involved:** NR |
| Sheltered housing [supported housing] | Norway | To support the residents (individuals with a serious mental health condition) in a rehabilitation process to prevent unnecessary admissions to mental health institutions. | The sheltered houses are organized as units consisting of one building complex with 7 to 30 one person fully equipped apartments with all amenities such as their own bathroom, kitchen and living room. They also have access to a shared accommodation room where the staff also attended. The residents are offered a 3-year tenancy agreement which has to be renewed at the end of the term. The sheltered housing looks like any other private homes in the area and is placed in different residential neighbourhoods. There are facilities like shops and walking areas nearby, and the distance to city centre is 10–20 min by bus. Live-in staff attended each unit 24 h a day, 7 days a week (24/7). Most residents have daily or weekly meetings with a mental health nurse or a service provider to discuss topics such as how to cope with the psychiatric disease, somatic health, household tasks, and financial issues. All services offered are voluntary, meaning that users can decide whether they want to accept help or not. | **Setting:** community (sheltered housing accommodation)  **Modes and format:** individual  **Frequency and intensity:** 3-year tenancy which can be renewed  **Types of treatment provider involved:** live in carers/MH staff, community mental health staff e.g. psychiatrists/OTs for meetings to discuss mental health |
| Supported accommodation [supported housing] | Canada | To provide stable housing for people with severe and long-term psychiatric disabilities, reduce symptoms, and promote community integration, independence, and well-being. | Supported accommodation has similar goals to group homes but offer a lower level of support. They act on a continuum with group homes such that people can "graduate" from group homes to supported accommodation. | **Setting:** Non-profit housing organisations  **Modes and format:** F2F  **Frequency and intensity:** NR  **Types of treatment provider involved:** Housing coordinators and directors |
| *Money and debt models* | | | | |
| Hertfordshire money advisors in mental health teams † | UK | To increase patient incomes and reduce mental health crises | Hertfordshire mental health project: The Mental Health Project takes referrals directly from community-based mental health clinical staff. Money advisors, who are located in mental health settings including CMHTs and Crisis Assessment and Treatment Teams, work with patients to increase their incomes and reduce mental health crises by improving mental health and wellbeing. The service also aims to reduce demand on mental health and specialist staff so they can concentrate on providing clinical care. | **Setting:** Community  **Modes and format:** NR  **Frequency and intensity:** NR  **Types of treatment provider involved:** NR |
| Mental health and money toolkit | UK | To support people to manage both their mental health and money difficulties | The toolkit has six sections: 1) mental health and money: where to start 2) getting ready to take action, 3) Understanding your finances, 4) navigating the benefit system, 5) support and 6) signposting and appointment planner. | **Setting:** Embedded within mental health consultations and appointments  **Modes and format:** F2F  **Frequency and intensity:** NR  **Types of treatment provider involved:** mental health practitioners |
| Mental health crisis Breathing Space | UK | To allow people in mental health crisis to focus on their recovery without worrying about their debt | If someone meets the definition for receiving mental health crisis treatment and has problem debts, they need to: • Get an Approved Mental Health Professional (AMHP) to complete and sign an evidence form that confirms they are receiving treatment for a mental health crisis. • The form must include details for a Nominated Point of Contact. • Once the form has been completed, they can be referred via the Single Point of Entry’ by their carer, an AMHP, a social worker, case coordinator (or others). When someone has had their debts entered into the Mental Health Crisis Breathing Space scheme, the organisations they owe money to cannot: • Demand payment of the debt. • Charge interest, penalties, or make any other charges to their debt. • Enter their home to take away anything they own. • Try to evict them from their home because they are behind with payments on their rent. Service users can also access debt and money advice whilst in the programme. Advisors can also help check they are claiming all the benefits they are entitled to | **Setting:** NR  **Modes and format:** NR  **Frequency and intensity:** ongoing  **Types of treatment provider involved:** money and debt advisors |
| Payeeship/representative payees | USA (3) | To help patients with a serious mental health condition become more financially autonomous by providing training, offering support, and gradually decreasing the agency’s role | 1) Patients voluntarily consent to the community mental health service or case managers acting as their representative payee. 2) Social security cheques are therefore mailed to the community mental health service and deposited to a state account 3) Initially, patients, their therapists and the money manager collaborate in planning a budget and schedule of funds disbursements. 4) Patients receive disbursements only during scheduled dispensing hours 5) unplanned funds needs are evaluated during a 48 hour hold or waiting period to allow consultation with the treating clinician.  Assistance with locating housing is also an important part of the programme. | **Setting:** Community mental health centre  **Modes and format: F2F**  **Frequency and intensity:** Often the initial budget to include a daily or biweekly allowance, with a gradual shift to a weekly or monthly allowance as clients improve their money management skills  **Types of treatment provider involved:** Money management services provided by a paraprofessional mental health worker who staffs the office from which funds are dispensed. a Primary clinician within a clinical case management team attends to patient's other needs |
| Sheffield money advisors in primary and secondary mental health teams † | UK | NR | This programme provides support for people in primary and community mental health services. Patients are assigned an adviser with mental health, debt and financial capability expertise, who then works with a client’s whole support structure to sustain a more financially secure future. Most of the work relates to debt and welfare benefits. | **Setting:** Community and Primary  **Modes and format:** NR  **Frequency and intensity:** NR  **Types of treatment provider involved:** NR |
| South London IAPT money advice pilot † | UK | NR | Clients are routinely asked about money worries during referral to IAPT HIT services. Where money worries are identified, clients are directly referred to Citizens Advice for a follow-up appointment with a money advisor. The two services, money advice and IAPT - can then be delivered either concurrently, or separately - dependent upon the client's preference. | **Setting:** Community  **Modes and format:** NR  **Frequency and intensity:** NR  **Types of treatment provider involved:** Money advisor and High intensity therapist (referral) |
| *Employment and education models* | | | | |
| Access to Work programme | UK | To help adults with a mental or physical health condition or disability access or stay in employment. | Individuals apply for the access to work grant via an online form or via the phone. Individuals give their contact details, workplace address, name and email address of person to contact to confirm you work there, unique taxpayer reference number (if self-employed), and information about your condition and how it affects your work, and what support you think you might need. After you apply, someone will contact you to discuss support, ask for more details about your application, and confirm person to contact at work. They will then send a letter with the decision and how much money you will receive in the grant. With the money from the grant, the Access to Work scheme can provide mental health support through Able Futures or Remploy, who provide either a tailored plan to help you get or stay in work, or 1:1 mental health support with a MH professional. Access to work can also pay for communication support in interviews, such as interpreters. They do not pay for reasonable adjustments that should be provided by the employer. Access to work can also provide practical support, such as interpreters, vehicle adaptations, or taxi fares to get to work, as well as a support worker or job coach. | **Setting:** community and workplace settings.  **Modes and format:** F2F and phone, individual support  **Frequency and intensity:** ongoing  **Types of treatment provider involved:** government, mental health professionals, support workers |
| BRIDGE supported education/IPS with education focus | USA (2) | To support adults with psychiatric disabilities to pursue education | The Bridge Program consists of 12 classroom modules that are held twice per week over 6 weeks. The Bridge Program incorporates principles of supported education and supported employment, within self-contained classrooms. The Bridge Program has four defining aspects: 1) Mentoring. Each participant has a one-to-one mentor. Mentors are 2nd year master’s level occupational therapy students. Mentoring is the collaborative effort of the mentor and participant to identify client-centred goals and develop and implement occupation-based interventions unique to the client to achieve these goals. Goals are systematically addressed each week through activities developed by the pair, and each weekly session concludes with a reflection on positive aspects and challenges affecting progress toward goals. A faculty member meets with each pair every week to supervise the process. After each session, the mentor completes a progress note detailing the participant’s progress toward the goals and a plan for the next session. Although goals and the activities to meet the goals vary according to each participant, there is a standard procedure to develop, address and evaluate achievement of the goals. 2) Higher education and/or employment goals. Each participant selects an educational and/or employment goal(s) to address in mentoring. If obstacles to the goal make the goal no longer feasible, the participant can change goals and remain in the program (e.g., if higher education is no longer feasible once the participant learns of costs involved, the participant can change to an employment goal). 3) Classroom modules. Modules are weekly presentations of academic and vocational topics for success in higher education and/or employment. They are implemented in a classroom format using lecture and small group experiences. Module topics include the following: 1) An exploration of training programs, degrees, and work options; 2) Study skills for school or work; 3) Time management skills for school or work; 4) Effective reading skills for school and job training; 5) Basic writing skills for school or job seeking; 6) Basic computer skills; 7) Introduction to Internet skills; 8) Basic math skills for school and job placement tests; 9) Use of library resources; 10) Public speaking strategies for school or work; 11) Professional behaviours and social skills; and 12) Stress management skills for school or work. The modules consist of an integration of lecture and lab activities through which participants can practice the skills they are learning. | **Setting:** Community college  **Modes and format:** Computer-based learning in a lab (group-based), and F2F mentoring  **Frequency and intensity:** 13-week program including 2-hour lecture and 1 hour one-to-one mentoring every week  **Types of treatment provider involved:** Occupational therapists and occupational therapy trainees act as mentors |
| Collaborative mental health care | Canada | To investigate the cost, effectiveness, and cost-effectiveness of a CMHC program designed to promote access to specialty care for people on short-term disability leave for psychiatric disorders. | People on short-term disability leave with identified mental health problems who met the criteria for an IME were referred to a CMHC psychiatrist. This psychiatrist used a standardised assessment package to evaluate the severity of the disability. With the employee’s consent, the CMHC psychiatrist contacted the employee’s family physician to discuss the diagnosis and to make treatment recommendations based on the assessment. The attending primary care physician had the option of referring the employee to one of the CMHC consulting psychiatrists for direct treatment through the public health care system. If the employee was referred for treatment, the consulting CMHC physician would provide 2 to 4 sessions with the goal of returning the employee to the care of the primary care physician as soon as possible | **Setting:** Primary care services  **Modes and format:** F2F  **Frequency and intensity:** 2-4 sessions  **Types of treatment provider involved:** psychiatrists |
| Individual Placement and Support (IPS) | Multiple (from our previous systematic review) | To support participants with severe mental illness who would like to work. | Employment specialists are embedded in clinical teams to help support a rapid search for competitive employment, and then provide time-unlimited and individualised support to  participants and employers. |  |
| Individual Placement and Support with education focus [Vocational support with early intervention (VIBE)- Major 2010)] | UK (2) USA (2) | To help clients with first episode of psychosis to gain open employment/mainstream education and focus on retention, helping clients to keep their education/jobs when they first came into contact with the early intervention team. | The Employment Specialist works with up to 25 clients at any one time and engages them on vocational issues, assesses vocational needs, proactively helps them to gain and retain work/education courses, provides welfare benefits advice, and addresses support needs including adjustments to enable people to retain work/education. The Employment Specialist develops good working relationships internally within the team but essentially externally with local employers, employment agencies, youth careers services, colleges and mainstream education and training providers and specialist employment/training services. One study (Nuechterlein 2019) additionally added a workplace fundamentals module which uses a group-based skills training approach to emphasise the social and problem-solving skills necessary to maintain a job/education. Some services are embedded within early intervention teams. | **Setting:** Community mental health teams  **Modes and format:** F2F individual and group learning  **Frequency and intensity:** Ongoing support for up to 3 years. Workplace fundamentals module provided for 6 months  **Types of treatment provider involved:** Employment Specialists and occupational therapists |
| IPS + Augmentation | Multiple (from our previous systematic review) | To support participants with severe mental illness who would like to work. | As IPS (above) with additional augmentation, which involves adding additional intervention components. |  |
| Mind@Work | Canada | To improve job tenure in people with psychosis | This programme is made up of 9 modules. In line with the predictors of job tenure identified in the proposed logic model, modules 1 to 6 focus on the self, while modules 7 and 8 concentrate on others in the work environment (e.g., colleagues, supervisors). The ordering of the modules has been carefully planned so that each one lays the foundation for the next. Modules should be delivered either in someone's existing workplace, or online. Where possible, sessions should be delivered during working hours. if the participants are comfortable disclosing their psychiatric diagnosis, the program is designed to foster collaboration with staff members and participants’ immediate supervisors, thus aiming to optimize knowledge transfer and to develop mental health literacy. Hence, it is recommended in the M@W program to involve a staff member (e.g., onsite psychosocial worker, counsellor) to act as group co-facilitator, and to provide immediate supervisors with a presentation of the program’s themes and strategies. Further, participants are invited to develop a work-related personal objective at the beginning of the program, which will then be discussed every other week to assess progress and deal with barriers. In line with the ‘cognitive behavioural therapy’ framework, participants are invited to complete weekly personalized homework (referred to as ‘challenges’ in the program) wherein they practice the personalized strategies that have been developed during the session. | **Setting:** In people's workplaces or online  **Modes and format:** F2F or online – group  **Frequency and intensity:** 9 modules, length not specified  **Types of treatment provider involved:** None |
| Social firms | England (9) | To create jobs or training for people who are job-disadvantaged due to discrimination. | The European network of Social Firms, (Social Firms Europe - CEFEC) defines Social Firms as follows: A Social Firm is a business created for the employment of people with a disability or disadvantage in the labour market; It is a business which uses its market-oriented production of goods and services to pursue its social mission (more than 50% of its income should be derived from trade); A significant number (minimum 30%) of its employees will be people with a disability or other disadvantage in the labour market; Every worker is paid a market rate wage or salary appropriate to the work, whatever their productive capacity; Work opportunities should be equal between disadvantaged and non-disadvantaged employees. All employees have the same employment rights and obligations. Social firms create jobs for people with disabilities and then promote the physical, social and mental health of their members by building an environment that encourages self-confidence, self-esteem and self-acceptance through fostering awareness and growth. | **Setting:** Workplaces  **Modes and format:** F2F  **Frequency and intensity:** Ongoing  **Types of treatment provider involved:** NR |
| The MENTOR programme | UK | Aims to help workers affected by mental health problems to address these problems and fulfil their potential | Mental health liaison workers are recruited and trained by mind, which deliver a range of 10 sessions-3 with line managers, 3 with employees and 4 jointly, over 3 months, mainly through video-calls but with some flexibility to meet face to face.  First joint session aims to set an action plan around what needs to change for their experience in the workplace to improve, this might involve the employee being supported in some way through reasonable adjustments, which might even include a change in job role. Follow up sessions monitor progress against goals and joint action plan, these sessions aim to help employees and line managers reflect on how they can better handle challenging situations at work and improve their confidence to speak about mental health issues in the workplace. | **Setting:** NR - video call  **Modes and format:** Individual, video-call as well as joint video-calls with line manager  **Frequency and intensity:** NR  **Types of treatment provider involved:** Mental health liaison workers |
| User Employment Programme | UK | To help people with mental illness gain employment in MH services | This is made up of two main elements: a supported employment programme based on the IPS model, and A Charter for the Employment of People who have Experienced Mental Health Problems that is designed to decrease employment discrimination. The User Employment Programme is integrated within the HR department and includes liaison with occupational health services, but is managed by the trust’s vocational services. The supported employment programme involves: assistance with recruitment (such as helping to complete application forms) and the transition to work e.g. any adjustments that need to be made, ongoing support to maintain employment, providing support to line managers. | **Setting:** NHS MH Trust  **Modes and format:** F2F  **Frequency and intensity:** NA  **Types of treatment provider involved:** NA |
| Peer-delivered self-management intervention | UK | To support people moving from crisis to ongoing support. | Peer support workers employed and managed by local NHS trusts. They are recruited through and competitive interviewing process other service users are involved in employment decisions. Peer support workers are fully trained, and provided supervisions by senior clinicians, experienced peer support workers not working in the study and members of staff of the involved crisis resolution team. Peer support workers work with up to five service users over a 6 months period but are free to agree to or decline to work depending on work commitments. The peer provided, self-management support includes: 1) Encouragement and help to complete a written personal recovery plan, 2) a willingness of peer support workers to share their own story and recovery journey where appropriate to model coping strategies and aid recovery, 3) encouragement to consider other supporters e.g. friends and family or other mental health support staff 4) encouragement and support to link with other ongoing sources of support such as local recreation groups. | **Setting:** Community  **Modes and format:** F2F individual  **Frequency and intensity:** 6 months pilot phase, 15 months intervention phase  **Types of treatment provider involved:** peer support workers |
| *Social isolation and inclusion models* | | | | |
| Arts based groups | Australia | To help improve the social networks and social belonging of people who had been referred from community mental health teams | NR | **Setting:** NR  **Modes and format:** F2F, group  **Frequency and intensity:** various  **Types of treatment provider involved:** NA |
| Community Navigator | UK (2) | To increase social connection and support, and reduce loneliness for individuals with severe mental illness and/or complex depression/anxiety | Had three main components. 1) Community Navigator helped participants use a social network mapping tool to map out what is important to the participant and potential areas for new social activities or strengthening existing connections. 2) Community navigator helped participants develop a "connections plan" which identified goals to increase connectedness and social relationships and the community navigator offered practical help or support in achieving these. 3) Community Navigator organised three group meet-up sessions with all participants receiving the program to allow people to meet, initiate friendships and share experiences of programme. | **Setting:** Multiple in-person locations  **Modes and format:** F2F mainly individually, with up to 2 group sessions  **Frequency and intensity:** 10 individual sessions, 3 group sessions  **Types of treatment provider involved:** Intervention providers recruited on basis of excellent interpersonal skills, awareness through personal or work experience of the challenges faced by people with serious mental illness, excellent knowledge of their local community and some previous work experience of supporting social inclusion and helping people develop social connections. |
| Compeer | USA (2) | To increase social support for individuals with a serious mental health condition | Clients were referred to Compeer by their professional mental health providers, then matched to a volunteer. Pairs of volunteers and clients committed to meet for 4 hours a month for 1 year, during which they were willing to engage in social, recreational, and supportive activities together. | **Setting:** N/A  **Modes and format:** F2F  **Frequency and intensity:** 4 hours per month for 1 year  **Types of treatment provider involved:** Volunteer led |
| Connecting People Intervention | UK | To help people recovering from psychosis to generate and mobilise social capital, and promote social interaction and engagement | The CPI (Webber et al., 2016) is an enhancement to usual care as it provides guidance to health and social care practitioners on how to more effectively help service users to develop their social networks. At its heart is a coproductive process of the practitioner and service user setting goals together and identifying opportunities for new social engagement. The focus of the work is on identifying potential new networks to engage the person with and supporting them to make new connections. The CPI requires an organizational commitment from teams to become more fully embedded within their local communities and for team leaders to foreground the CPI model in team meetings and individual supervision. Practitioners are expected to use the model to inform their daily practice with all service users. The CPI model is not prescriptive about the steps the practitioner needs to take, but the practice guidance provides examples and advice to inform the process of engaging service users with their community (geographical, interest, or personal). Two-day training was provided to participating teams in the CPI in sessions led by a team of five trainers (two to three per session) | **Setting:** Multiple in-person locations: National Health Service (NHS) community mental health teams, third sector agencies, and one was a local authority day service for adults with a learning disability.  **Modes and format:** F2F  **Frequency and intensity:** ongoing  **Types of treatment provider involved:** NHS mental health teams, third sector agencies, local authority day service, social workers, OTs and support workers at CMHTs |
| Consumer providers | USA (2) | To help people with a serious mental health condition achieve their recovery goals through providing support | Consumer Providers (CPs) are individuals with a history of a serious mental health condition who draw upon their experience to provide services to others with similar mental health problems. CPs can work in a variety of clinical and rehabilitative settings, performing roles such as outreach and engagement, case management, and a variety of other functions (e.g., job coach). CPs can be either paid or volunteer roles and work to 'model recovery' to patients. | **Setting:** Multiple  **Modes and format:** F2F  **Frequency and intensity:** ongoing  **Types of treatment provider involved:** Consumer providers |
| Creative workshops | Spain | To help people with a serious mental health condition improve their wellbeing and social-connectivity | Bar slight modifications, the structure of the creative workshops, has been the same since 2006. The workshops are structured in six consecutive weekly sessions each of them three hours long (total 18h). Every year the CAAC holds three temporary exhibitions by contemporary artists. These temporary exhibitions drive the topics and themes that are developed within the creative workshops. The workshops were structured in two parts of 90min each. During the first part, participants visited a specific, selected section of the exhibition. An art facilitator provided context and background information about the artwork, artist and other relevant facts and facilitated a discussion with participants, who were encouraged to ask questions and share their opinions and impressions. The second part also took placed at the CAAC and consisted of a hands-on workshop. Participants were invited to gather around a large table and use provided materials for painting, drawing and sculpture. Participants were encouraged to create their own artwork. On occasion the art facilitator suggested a specific topic such as “journey” or “seed” to inspire their artwork. During the first 75min, the participants freely worked on their individual artwork using all the materials available. The artistic techniques employed varied but were largely drawings and acrylic paintings on cardboard or canvases. To close the session, each participant shared with the group their artwork and explained the process and intention. Participants were encouraged to comment on other participants’ artwork, ask questions, make suggestions or new interpretations. | **Setting:** A museum  **Modes and format:** F2f - group (15-30 ppts)  **Frequency and intensity:** The workshops are structured in six consecutive weekly sessions each of them three hours long (total 18h). Within the same week, four different groups of around 15–30 service users participate in these workshops as well as their carers (i.e. keyworkers).  **Types of treatment provider involved:** Clinical psychologist (to manage project), art facilitator |
| Education groups | UK | To improve knowledge in people with schizophrenia living in the community about their condition, leading to positive changes in social functioning and quality of life. | Groups were run in community settings with cafes and funds available for coffee and snacks. Groups ran for 20 weeks with each session lasting an hour and a half. Sessions alternated between an information session (short presentation and discussion) followed by a problem-solving session. Sessions covered were: 1) what does schizophrenia mean to you? 2) what is schizophrenia? 3) problem solving- managing symptoms 4) treatment of schizophrenia 5) problem solving- medication and its side effects 6) rehabilitation; community resources 7) problem solving-employment, leisure 8) early signs of relapse 9) problem solving-managing relapse and symptoms 10) what causes schizophrenia 11) problem solving- using alcohol and other drugs 12) families and schizophrenia 13) problem solving-relations with the family, managing negative feelings 14) relating to other people- family friends and others 15) problem solving- social skills, assertiveness, telling people about your illness 16) stress 17) problem solving- stress management, withdrawal 18) using services 19) problem solving - housing, social work, voluntary agencies, legal aspects and patients’ rights 20) where do you go from here? | **Setting:** Community  **Modes and format:** F2F, group  **Frequency and intensity:** once a week for 20 weeks  **Types of treatment provider involved:** Community psychiatric nurses, one occupational therapist and one registrar who had received training to run the groups |
| Expand your world | UK | To provide a safe space to explore the topic of 'expanding your world', including social, vocation and health activities for people experiencing mental distress | On a practical level the group room in our centre worked well for hosting these workshops. It had a digital projector, stereo speakers, good furniture and the space felt safe, airy, warm and comfortable. It was spacious enough for around 15 people. A variety of different formats were facilitated: whole and small group discussions, presentations, a film made by one of the service users in the team and scenarios. Participants also undertook their own research and reflections on goals and barriers. Content of workshops included: Finding meaning in life; Stigma and Self-Imposed Stigma – how it can shrink your world; Planning your own ways forward; Social Inclusion – what does it mean?; Choice, trying things and saying no; Replacing fear with possibilities – solutions to barriers; Five Ways to well-being – a model; Learning from experiences | **Setting:** Outpatient services  **Modes and format:** F2F, group  **Frequency and intensity:** 5 weeks - 1 workshop a week  **Types of treatment provider involved:** Facilitators included service users and team members (maybe clinicians, not clear from report) |
| Focus, Act, Connect Every-day (FACE) | USA | To support individuals with a history of mental health issues, substance misuse, trauma, homelessness and incarceration to engage with the community in various activities | FACE members (approximately 20 people) come together on a bi-weekly basis to support one another in engaging with the community, including planning activities and events with community partners. These events have included leading mural-making at neighbourhood festivals, packing groceries for people in need, and facilitating conversations with community groups. They have also joined forces with Witnesses to Hunger, and FACE members have recruited new members, attended planning meetings, spoken at roundtables with federal elected officials, and organized rallies and other events to advocate for policies to better address the upstream causes of food insecurity and economic struggle. | **Setting:** outpatient, community setting  **Modes and format:** F2F, group  **Frequency and intensity:** bi-weekly (fortnightly), ongoing  **Types of treatment provider involved:** NR |
| Groups 4 Health | Australia | To improve general health and life satisfaction in people who are socially isolated | G4H's five modules give people the knowledge and skills they need to manage their social group memberships, and the identities that underpin them, effectively. Each module contains a series of exercises and discussions targeting the different aspects of group life identified within SIMIC. The first module, Schooling, raises awareness of the beneficial effects that social group memberships have for health. It highlights the costs of ignoring the social dimensions of health and points out that failure to use all the social resources at our disposal generally leads to suboptimal health outcomes. However, the module also makes the point that it is within people’s power to counter these effects by learning how best to develop, maintain and harness group-based social resources. Module two, Scoping, focuses on the range of group-based resources that people have, or ideally should have at their disposal, to optimize health. This module engages participants in the process of social identity mapping (Cruwys et al., 2015). This tool was developed to explore respondents’ social identities, in order to assess their current social functioning and develop a sense of how they would ideally like to function in the future. The third module, Sourcing, focuses on identifying and strengthening existing valued social identities with a view to optimizing and sustaining these in the longer term. Module four, Scaffolding, uses the G4H group as a model for establishing and embedding new social group connections whilst at the same time exploring strategies to identify which connections to develop and enact through a social plan of action. The goal is to trial these social plans between this and the final module, which takes place at least one month later. The final module, Sustaining, is a booster session held one month later that aims to troubleshoot any difficulties that have arisen in the course of implementing these social plans. It also revisits social identity maps, created in Module 2, to see how they have developed in the course of the program. The social foundations that have been identified and developed in the preceding four modules are also reviewed with a view to encouraging their long-term maintenance. | **Setting:** NR  **Modes and format:** F2F group  **Frequency and intensity:** 5 modules, timeframe not stated  **Types of treatment provider involved:** NR |
| Guided peer support | Netherlands | To provide peer-to peer interaction for adults with schizophrenia or other psychotic disorders to have a positive effect on social life and quality of life | First, people are encouraged to work in pairs to exchange positive experiences from the previous two weeks (ten minutes). Next, all pairs share with the group the stories they just heard (ten minutes). Then the nurse initiates the general discussion by asking, “What have you just heard that could be of interest for the whole group?” Next, the participants choose the theme of the session (five minutes), briefly introduced by the nurse (two minutes). The themes should relate to the illness, for example: living with schizophrenia, telling others about your illness, or resuming your job. After a 15-minute break, they share their experiences about the theme in pairs (15 minutes), participants reconvene for the final plenary session (25 minutes). At the end, the nurse briefly summarizes the session (eight minutes). | **Setting:** Community  **Modes and format:** F2F group-based  **Frequency and intensity:** 16 x 90-minute sessions biweekly over 8 months  **Types of treatment provider involved:** Nurses |
| Hearing Voices Network support groups | Australia | To help people who hear voices improve across several recovery domains | Although they are formed on shared experience, the setup, content and practice within each group is very different. Further, HVGs are run in an open format, which encourages group members to come and go as their needs change. | **Setting:** NR  **Modes and format:** NR (but F2F implied)  **Frequency and intensity:** ongoing  **Types of treatment provider involved:** non-clinical facilitator |
| Occupation and Social Skills Training | Turkey | To strengthen the transfer of skills to the real world and facilitate social participation for people with schizophrenia. | The SST consists of two modules: conversation skills and problem-solving skills. It is a group-based program (6 participants), and the first two sessions were devoted to meeting the group members and introducing the program. The first module comprised four sessions (3–6) and aimed to introduce basic conversation skills and practice the skills with role-playing. The second module covering sessions 7–9 focused on teaching the basic principles of problem-solving skills and reinforced the ways of problem-solving with examples. The final session was devoted to reviewing and evaluating all the sessions. Behavioural techniques were used to help transfer the learned skills to daily life for two interventions. The features of each skill were defined, and the skills were practiced step by step. Scenarios were realized with role-plays, and the skills were learned and practiced with positive reinforcement and corrective feedback. It was also aimed to make the skills permanent with homework assignments. The OT+SST program is a person-centred program addressing the needs and desires of people with schizophrenia and a group-based program (six participants) consisting of the SST (basic conversation skills and problem-solving skills) and OT programs (activity planning skills and recreational skills programs). The Occupational Therapy program was administered on a group basis, giving priority to individual needs. Individual needs were determined according to the problem areas of occupational performance in the COPM at onset and goals set with the client. Encouragement and motivational interviewing were used to generalize the skills learned in the clinical environment to real-life conditions, enhance adherence to therapy, and thereby be able to meet activity needs and to use personal capacity. Participants were monitored throughout the program with homework, and sustainability was encouraged by providing feedback. The themes in this combined intervention program (OT+SST) included preparation for community living, practicing basic conversation skills, identifying desired activity goals and finding strategies to achieve them, problem-solving strategies to deal with daily living challenges, and developing leisure skills. Each session contained a brief education section, a group activity, in vivo exercises, and homework assignments. The first two sessions were devoted to both meeting the group members and introducing the program, and then sessions 3–6 focused on basic conversation skills. Sessions 7–9 engaged the clients in problem-solving skills with examples of daily living difficulties. Sessions 10–12 included activity training and follow-up of individualized occupational therapy programs determined in a one-on-one interview. The importance and benefits of occupational engagement in the recovery from schizophrenia were explained by providing knowledge and awareness about daily living activities with an activity training session. A daily activity schedule was used and followed up according to participants’ intervention plans to practice their performance. So, they became habitual. The activity schedule of the group members was reviewed in each activity session, and their real-life experiences were evaluated and discussed with them. Besides, at the end of each session, the activity plan was reviewed individually with the therapist to help the participants actualize skills in daily life. The last three sessions were administered to develop recreational skills. It was aimed to practice communication skills and transfer those skills into daily life by simulating the selected recreational activity during the recreation skills sessions. The recreational activity in the study was meeting at the patisserie. The activity steps are summarized as (1) selecting the patisserie, (2) meeting friends at the patisserie, (3) asking for the menu, (4) giving an order, (5) chatting with friends and drinking coffee, (6) asking for the bill, and (7) leaving the patisserie and saying goodbye to friends. The final session was devoted to reviewing and evaluating all the sessions. | **Setting:** Community mental health centre  **Modes and format:** F2F group and individual  **Frequency and intensity:** 10 (SST) or 16 (OT+SST) 50-minute weekly sessions  **Types of treatment provider involved:** Occupational therapists |
| Participatory Video | Canada | To help personal recovery for people with a serious mental health condition | Participants were recruited from mental health drop-in centres for people with a serious mental health condition in 3 Canadian cities, leading to 3 workgroups (1 in each city). Each workgroup met approximately twice per week in a spare room in each mental health drop-in centre over a 2-year period (2015–2017). Each workgroup was supported by a videographer-facilitator who gave initial and ongoing training in all aspects of video production, including scripting, filming and editing. Some training sessions were also devoted to critical and analytical thinking. After the initial training, workgroups were tasked with producing educational videos about mental illness, with complete editorial control over topics and themes presented in the videos. | **Setting:** Mental health drop in centres  **Modes and format:** F2F, group  **Frequency and intensity:** twice a week for 2 years  **Types of treatment provider involved:** NA |
| Peer Specialists | USA | To model recovery for service users (veterans with a dual diagnosis of mental illness and substance abuse) and improve social networks, leading to positive changes in social functioning and quality of life. | Peer specialists help to deliver interventions, using a triad approach, where both a Peer Specialist and a Case Manager or other professional work together with each patient to support recovery goals. There are curriculum-based sessions of about one hour in length, and 20 “unstructured” sessions of varying length, to include informal visits, conversation, and activities in the patient's home or in the community. Peer specialists receive weekly supervision | **Setting:** outpatient services, people's homes & community  **Modes and format:** F2F, group and individual activities  **Frequency and intensity:** varied  **Types of treatment provider involved:** Peer support workers |
| Project Connect | USA | To support people with mental illness in creating and sustaining connections in their communities, based on their interests and talents and in ways that they define to be most helpful to them. | Project Connect staff introduce participants as equal partners with other individuals or organizations rather than as people in need of charitable socialization. This introduction reflects and reinforces Project Connect’s status as a supporter, but not a primary support structure, for the participant. Project Connect remains available to support or re-evaluate connections, but participants and the community partner broker their own relationships going forward. Project Connect does not require referrals for participation, nor does it assess “readiness” for it. Although both are generally required for entry into treatment, they are irrelevant and inappropriate to community settings, given that such tools are not part of normative community processes. At times, a community organization might require an application, interview, or background check to allow a participant to do volunteer work, as it would with any other applicant. In this case, Project Connect staff is available to assist the participant in navigating that process. The Project Connect process begins with an introductory meeting with an interested participant at a place and time determined by the participant. Because participation in Project Connect is only partially a function of a person’s status as a client in the mental health system, this first decision is a practical means of allowing participants to exercise their “nonpatient identities.” The site of the meeting can range from traditional mental health settings to public spaces like libraries and coffee shops to private settings, such as participants’ homes. During this initial meeting, participants are asked to share their interests, the types of connections they want, their histories of brokering similar connections in the past, and their transportation options. Participants also talk about how they want the connection to be arranged. This can range from a list with potential connections and contact information that the person will act on to staff-initiated connections in which a Project Connect staff member contacts a potential community partner or arranges a three-way meeting. Staff-participant relationships after the connection also vary. In some cases, staff will take the bus with participants to familiarize them to a new route or to support and ease the person’s anxiety. Project staff may also meet participants near their homes and go with them to new and unfamiliar locations. Some participants request several meetings with project staff to refine their ideas and plans for how connections will happen. | **Setting:** Location decided by participant  **Modes and format:** F2F individual  **Frequency and intensity:** NR  **Types of treatment provider involved:** NR |
| Recovery narrative photovoice | USA | To facilitate recovery, empowerment, community integration and positive identify for people with severe mental illness | Recovery Narrative Photovoice program curriculum was manualised, including psychoeducational content, handouts, writing exercises, and activities to assist participants with con-structing empowering narratives of recovery and identity given the stigma associated with serious mental illnesses. Three “photo missions” were assigned to participants over the course of the 10 sessions to elicit photography and narratives focused on recovery and identity, called: “Who I Am,” “My Story,” and “My Recovery” | **Setting:** psychosocial rehabilitation and education centre  **Modes and format:** F2F, group  **Frequency and intensity:** 10 weeks, meeting once per week for 2 hr each meeting  **Types of treatment provider involved:** Groups were led by 1st author on paper and a peer specialist |
| Role Development programme | USA | To help individuals with schizophrenia develop task and interpersonal skills | NR | **Setting:** NR  **Modes and format:** F2F  **Frequency and intensity:** NR  **Types of treatment provider involved:** Forensic rehabilitation staff |
| Scatter -site & Congregate housing | USA | NR | Independent scatter-site housing included only housing categorised as “supported housing,”(defined as “permanent housing in single and shared apartments in the community...services are provided on an as needed basis”(Centre for Urban Community Services, 2015a, 2015b) while congregate housing included “supervised community residence,” “congregate support,” “dual disorder community residence,” “congregate treatment,” “community residence/single-room occupancy” and “residence for adults”(all of these models share the common elements of group-based housing with 24-h on-site staff). | **Setting:** Residential  **Modes and format:** NR  **Frequency and intensity:** ongoing  **Types of treatment provider involved:** NR |
| Social cognition and interaction training + peer mentoring | Israel | To improve social functioning, attributional biases, theory of mind and emotion recognition in people with serious mental illness. | Participants received various forms and varying degrees of social, leisure, support, and employment services, and all received the same social-mentoring service. This social-mentoring service includes three weekly meetings with a mentor to support in vivo efforts by consumers to take practical steps toward achieving personally meaningful goals, for example, organizing their finances, obtaining needed information, filling in forms, and registering for a course. One of the three weekly mentoring meetings was dedicated to a social cognition and interaction training session | **Setting:** Community  **Modes and format:** F2F with a mentor  **Frequency and intensity:** 3 1hr meetings with mentor per week  **Types of treatment provider involved:** Social mentors- staff at mentoring agencies. Clinicians with training in social cognition and interaction training provided this aspect of the intervention |
| Social network intervention | Italy | To trigger or facilitate relationships to increase the natural social network of patients suffering from schizophrenia. | Usual therapeutic services provided by community treatment teams, alongside identification by staff of possible areas of interest for individual patients which take place outside of the services resources and with members of the community - the intervention is intended to encourage patients to participate in social activities they might have done but for their disability. A member of staff is designated to provide the necessary support for the patients’ integration in the specific activity for 3-6 months, and within this time more than 1 alternative can be suggested. The intervention could be carried out directly by a member of the staff or through natural facilitators such as family members, neighbours or volunteers | **Setting:** Community  **Modes and format:** F2F  **Frequency and intensity:** NR  **Types of treatment provider involved:** Community mental health staff, volunteers, family members |
| Social Prescribing | UK | To improve patients' long-term psychological/social issues and provide a tailored package of support to enable access to community services, groups and activities | New 'holistic' service (Rotherham Social Prescribing Mental Health Service) tailored to needs of secondary mental health services, to augment existing treatment pathways. Funding was provided for social prescribing link workers to provide support, and a 6-month pathway was developed in consultation with a CMHT to help with transition from secondary mental health treatment to community-based activities. CMHTs stay involved for up to 6 months, where they then establish if they can be discharged from the CMHT (applied flexibly, considering personal circumstances). | **Setting:** unclear  **Modes and format:** individual  **Frequency and intensity:** 6-month pathway  **Types of treatment provider involved:** link workers, CMHT staff (psychologist, psychiatrists, social worker etc.) |
| Social recreation programme | Canada | To help adults with SEMI establish and maintain satisfactory social bonds | A multi-level, multi-strategy process is applied to facilitate the process of relationship development. Program delivery is directed at two levels: individual and social or environmental. Furthermore, each level requires the use of a flexible array of strategies. Strategies are derived from two sources: the Ottawa Charter for Health Promotion and the community and social psychology literature. The Charter, in addition to developing personal skills, encourages building healthy public policy, creating supportive environments, strengthening community action, and reorienting health services. The psychology literature identifies strategies for helping the lonely and socially isolated person, that range from social competence training, to altering aspects of the physical environment, to increasing the frequency of friendly interaction between people. Together, these strategies translate into specific program activities. While participation in activities is voluntary, individuals are encouraged to participate in the breadth of activity possibilities. An advisory committee, drawn from people within the program, plays an important role in program planning and review. Other services offered to people include information and referral, (emotional, information, and instrumental) support, outreach, assessment, and goal review, as well as collaborative work with family or professionals. | **Setting:** Community mental health team  **Modes and format:** F2F group and individual  **Frequency and intensity:** NR  **Types of treatment provider involved:** NR |
| Structured social coaching intervention | UK | To expand social networks for people with psychosis | 1) Introduction: The social coach and the patient introduce themselves (usually F2F but can be done over video call) 2) Clarification of the remit of the intervention: The professional explains and discusses the focused remit of the intervention, i.e. that it aims to expand social networks and that all other therapeutic issues have to be addressed elsewhere. (3) Exploration of past and current activities: The social coach explores past activities that involved social contacts; this should be done chrono-logically covering the adult lifetime of the patient from the age of 15 years onwards, and stepwise for periods of 5–10 years. At the end of the exploration, professional and patient go through the list of activities (if any) and discuss to what extent the patient enjoyed each activity. Solution focused therapy techniques (e.g., identifying what went well and what worked) can be used. 4) Motivation for change: The professional explores and discusses the patient’s motivation to change and expand their social networks. Motivational interviewing techniques (e.g., identifying change talk) can be used. (5) Options for activities: Professional and patient discuss which new activities (or expanding existing ones, respectively) the patient considers. (6) Information: The professional provides as much helpful information as possible about options in the given locality for patients’ preferred activities. Professional and patient discuss the practicalities and some-times decide to obtain further information. In this step, the patient is encouraged and supported to find information him/herself. Yet, if this is a substantial hurdle, the professional provides as much direct support as needed. (7) Consideration and decision: Once options have been identified the patient is asked to consider taking it up. If the patient is ambivalent, patients are encouraged to take time, for example, a week until the next meeting or a phone call, to think about it.8) Definition of activity Finally, the patient decides on the type of activity and some specification of the actual steps (e.g., twice per week attending a certain class, but not necessarily on which days), so that professional and patient can assess afterwards whether the activity has been completed or not. The task gets documented for the patient, for example, written on a piece of paper that the patient takes along. The main aim of these initial meetings is to introduce the intervention, explore participants’ social history and discuss preferences and options for activities. The participant then selects one social activity to focus on during the remaining meetings and actions are agreed. The subsequent meetings include discussions around challenges and progress and take place monthly, lasting about 20 min each. The final meeting is face- to-face and is used as both a summary of progress and to plan for future social activities after the intervention. Social coaches are trained in one session lasting 3 hours, normally in a group format (although one- to- one sessions can be arranged). | **Setting:** NR  **Modes and format: F2F or video**  **Frequency and intensity:** Social coaches meet patients at least three times but ideally monthly over the 6-month intervention period. Intervention can be stopped at any time if participant requests it  **Types of treatment provider involved:** The intervention is delivered by clinicians from different backgrounds (e.g., psychologists/assistant psychologists, social workers, nursing staff, occupational therapists and medical doctors), |
| Supported socialisation | USA (1), Ireland (2) | To decrease social isolation for people with severe mental illness by helping them to create social networks | The Volunteer group were matched with a community volunteer partner, undertook social/leisure activities with them and received a stipend of €20 monthly to defray the costs of socialising. The intervention was designed to promote an ‘intentional friendship’ between a volunteer and mental health service user comparable with ordinary social friendships outside the mental health system. Volunteers were recruited from the local area and were aimed to be representative of the study participants. All volunteers attended an interview, were police vetted and received a 1-day training session to prepare them for their role (training components included mental health literacy, stigma awareness, activity planning and problem solving). Solitary participants received the monthly stipend for the 9 months to encourage engagement in self-driven socialisation. Both groups received an activity suggestion pack that detailed free or low-cost socialisation resources in their local community. | **Setting:** Community settings  **Modes and format:** F2F with volunteer partner  **Frequency and intensity:** A few hours a week for 9 months  **Types of treatment provider involved:** NA |
| TREE (Toward recovery, empowerment and experiential expertise) recovery programme | Netherlands | To teach persons with severe mental illness to manage their own lives and counteract marginalisation in society through enabling participants to offer each other mutual support. | i) Self-help working groups: Each group consisted of a maximum of eight participants plus two peer workers who acted as facilitators. The workshop activities were based on recovery and empowerment and organised as a self-help rather than a therapeutic group activity. ii) One-day training course: The seminar programme targeted patients receiving long-term care and their professional mental health care workers. They could only take part if they came as a pair (patient and professional). Mental health care managers facilitated the professionals to attend the seminar during working hours as part of the internal education programme. iii) Training course: “Making a start with recovery” was a familiarisation course on the meaning of the concept of “recovery” for patients using long-term psychiatric care. | **Setting:** Community and residential  **Modes and format:** F2F group based  **Frequency and intensity:** Self-help groups - two-hour meetings every fortnight for 52 weeks for late starters and 104 weeks for early starters  **Types of treatment provider involved:** Senior peer workers |
| Volunteer befriending programmes | Colombia | To support people with severe mental illness | Participants may be grouped with others with similar interests and who live in close proximity to each other (In Botero-Rodriguez, 1-2-1 model was adapted to form groups of participants to spend time with volunteers, traditional model is 1-2-1 (Priebe 2020)). All volunteers receive training which covers information about the programme, the symptoms that the patients may present with, their responsibilities, resources for supervision, support from the research team, and emergency procedures, and also receive a written manual of the intervention. The volunteers contact the patients and organise group/individual meetings. Volunteers are given the opportunity to move to a different group if they feel the original allocation was difficult for them. Volunteers are encouraged to organise different indoor and outdoor activities in their meetings (e.g., go for a coffee/walk/picnic in the park). Following each meeting, volunteers inform the research team about the type of activity, the duration, and the content of discussions. Monthly social events including food and/or an activity (for example a picnic in a park, an art workshop) are organised to provide opportunities for different volunteers and patients to meet and interact. | **Setting:** In the community  **Modes and format:** F2F group based  **Frequency and intensity:** 12 bi-monthly meetings for 6 months (Botero-Rodriguez) or weekly meetings for 12 months (Priebe 2020)  **Types of treatment provider involved:** volunteers |
| *Family models* | | | | |
| Family support services (Parents, advocacy, coordination, education; PACE) | USA | To help parents with a serious mental illness build on their strengths and provide the services and supports needed to avoid losing custody of their children. | The services offered by FSS.PACE include 1) case management, 2) individual therapy for both adults and children, 3) medication and illness management, 4) parenting skills training and child development education, 5) problem solving skills training, 6) strength identification for the entire family, and 7) advocacy. FSS.PACE also offers 24-hour on call support, emergency assistance, transportation, a monthly social support group for mothers, tenant-landlord mediation, financial assistance, crisis planning, transitional planning (for example, after hospitalisation), and housing assistance. The primary site of service provision (the family home) is integral to the program success. Collaborative arrangements are also made with other service providers e.g. schools, juvenile justice, housing, mental health and court services. | **Setting:** Patient's homes  **Modes and format:** F2F  **Frequency and intensity:** ongoing  **Types of treatment provider involved:** Clinical case managers, alongside centre staff who have expertise in areas related to the care of FSS/PACE and who provide both formal and informal consultation. |
| Integrated family treatment | USA | To help parents with severe psychiatric disabilities improve parenting skills and the home environment | Components of integrated family treatment include 1) engagement into treatment, 2) assessment, 3) linkage to environmental supports, 4) education about child development and 5) parenting skills training, modelling and coaching. | **Setting:** NR  **Modes and format:** F2F  **Frequency and intensity:** NR  **Types of treatment provider involved:** Family specialist clinician |
| Mothers and children’s project | USA | To teach mothering skills to mothers with schizophrenia and provide early intervention | 1) Mothers are referred to the service from public social service agencies, domestic courts, mental health clinics and maternity wards. 2) A mother is visited at home by the program director who reviews their diagnosis and assesses their level of functioning to offer a realistic view of the home environment and any special difficulties the mother may be facing. 3) The main program consists of 2.5-hour meetings held once a week in a community church-transportation is provided. 4) Mothers and children spend the first 45 minutes in small groups led by a volunteer supervised by a child specialist. the groups focus on developmental education, directed play and role modelling. These small groups give mothers the opportunity to discuss problems with staff and peers and to observe other mother child interactions. 5) Each week one mother and her child meet with the child specialist to discuss individual issues. 6) in the afternoon the mothers meet in a separate group while children are in a therapeutic nursery. The group is led by a social worker and a volunteer and provides a forum to discuss parenting | **Setting:** Community  **Modes and format:** F2F, group  **Frequency and intensity:** weekly  **Types of treatment provider involved:** Staff consist of a psychologist, a child development specialist and a social worker, and five volunteers, two research trainees, one domestic assistant who provides lunches during meetings. |
| Thresholds mothers project | USA | To support homeless mothers with mental illness | A problem-solving approach to psychosocial rehabilitation and intensive case management- practical problems in daily living are a focus of the model, and mothers are helped to meet their basic needs, stabilize their living arrangements, and begin addressing psychiatric symptoms. Case managers help to secure entitlements, find independent apartments, and function as representative payees when needed. Care managers also assist with enrolling children in regular or special education. | **Setting:** NR  **Modes and format:** F2F  **Frequency and intensity:** NR  **Types of treatment provider involved:** Care managers |
| *Victimisation models* | | | | |
| BRAVE (Better Reduction and Assessment of Violence) | Netherlands | To improve detection and referrals on DVA in mental health care, using a gender sensitive, system-level program that targeted community mental health (CMH) teams, and also provided mental health training for DVA professionals. | The BRAVE intervention consists of three parts: (a) a training course for CMH teams to increase knowledge, attitudes, and skills in managing DVA, (b) a knowledge, attitudes, and skills training course on mental illness for DVA professionals, and (c) the provision and implementation of a direct care referral pathway between CMH services and DVA services for victims of DVA. The BRAVE intervention is based on the successful LARA intervention (Trevillion et al., 2014) | **Setting:** Community  **Modes and format:** Group training, individual referral pathways  **Frequency and intensity:** 8-hour training course, divided into two four-hour sessions (CMH staff). DVA professionals received four 3-hour workshops about mental illness. Referral pathway ongoing.  **Types of treatment provider involved:** CMH team staff, DVA professionals |
| Health Pathfinder project | UK | To improve the awareness, knowledge and skills of health professionals and the systems within which these professionals work, in order to increase professionals’ ability to routinely and sensitively enquire about domestic violence and abuse and to increase system ability to support professionals in doing this work effectively and consistently. This is expected to increase the confidence of victim survivors to disclose, and to receive a professional response that in turn leads to a timely referral to specialist services. | Individuals experiencing domestic violence and abuse and presenting at one of the eight intervention sites were asked by trained healthcare professionals, including, where relevant, the named safeguarding lead, about whether they were experiencing domestic violence and abuse. The healthcare professional or safeguarding lead is then meant to refer this individual to the local specialist domestic abuse service, Independent Domestic Violence Advisor (IDVA) or Advocate Educator associated with that particular site following the intervention protocol. The provider then follows their standard operating procedures for managing referrals for domestic violence and abuse. | **Setting:** outpatient, community and inpatient mental health services  **Modes and format:** various  **Frequency and intensity:** ongoing  **Types of treatment provider involved:** community mental health teams, inpatient teams, and acute teams. |
| LARA (Linking Abuse and Recovery through Advocacy) | UK | To support psychiatric service users who experience domestic violence | (1) Four hours domestic violence training for clinicians (on entry to study), illustrating how to identify and respond to domestic violence (delivered by a senior clinical psychologist (author RAD) who specializes in supporting victims of domestic violence). (2) Domestic violence manual for clinicians (developed by the research team), incorporating good practice guidance and local/national domestic violence services. (3) Six hours mental illness training for domestic violence advisors (who provided domestic violence advocacy), including definitions of disorders, treatments and service provision (delivered by a senior psychiatrist (author LH)). (4) Direct referral pathway to domestic violence advocacy (developed by the research team) for service users experiencing past year violence. (5) Provision of integrated domestic violence advocacy for service users, modified for this study and delivered by domestic violence advisors (seconded from a local voluntary sector organization). Advocacy comprised emotional and practical support, including domestic violence education, facilitation of support groups, safety planning and legal/housing support. In this study, each CMHT had a named advisor who was available to discuss/take referrals and feed outcomes back to the team and to regularly attend clinical meetings to discuss cases and provide domestic violence education. (6) Information campaign (posters and leaflets in waiting rooms and toilets) highlighting the problem of domestic violence and support available. | **Setting:** Community (CMHT), domestic violence advocacy teams  **Modes and format:** F2F, individual and group training  **Frequency and intensity:** NR  **Types of treatment provider involved:** CMHT staff-care coordinators, psychologists |
| LINKS project | UK | To increase identification of patients experiencing domestic abuse, increase referrals of patients into specialist domestic abuse services, and increase awareness, knowledge and confidence of community and inpatient mental health staff in responding to domestic abuse. | The Mental Health Independent domestic violence advisor (MH-IDVA) was responsible for providing training to Barnet staff working for Barnet, Enfield and Harringay Mental Health Trust (BEH-MHT), giving ongoing support for health professionals, and providing a proactive and culturally sensitive support service for women and men over 16 years of age who are experiencing domestic abuse who were referred by Barnet staff in BEH-MHT. The job description included the following expectations, but was not limited to: To support ongoing training for health professionals based in Barnet from BEH-MHT to increase their understanding, identification and response to domestic abuse, to promote awareness of the experiences and needs of women and men living with or experiencing abuse, particularly in relation to their mental health, to encourage health professionals to ask women and men about their experience of abuse and respond, record, safety check and refer to Solace Women’s Aid or other appropriate specialist services, to build and maintain effective relationships with general practice teams and agencies in the borough, to carry out risk assessments in a timely manner and refer high risk cases. To work with survivors referred to the MH-IDVA to agree safety and support plans working to reduce risk and meet identified needs including through making referrals to other services to keep them and their children safe, to provide an effective and well managed casework service working to targets agreed in the safety and support plan and to act as an advocate for survivors keeping their safety at the centre of all coordinated and community based responses, to advocate on behalf of service users with external agencies where appropriate, to attend case review meetings and effectively communicate across both teams, and to keep and maintain accurate and confidential records of all work undertaken. The introduction of the MH-IDVA did not create a new referral pathway into services. Referrals to the MH-IDVA were made by staff on an ad hoc basis. Referrals were intended to be informal to facilitate a greater number of discussions with staff about how to approach patients who disclose domestic abuse. Once a patient had been discussed with the MH_IDVA, she would typically include the patient as part of her caseload. | **Setting:** community and inpatient  **Modes and format:** F2F settings with staff individually and in groups, and 1:1 with patients referred to the pathway  **Frequency and intensity:** training on domestic violence delivered by the MH-IDVA in two 3-hour sessions  **Types of treatment provider involved:** mental health team staff, community and inpatient service users who have experienced/are experiencing domestic abuse |
| Self-wise, Other-wise, Streetwise (SOS) training | Netherlands | To prevent total, violent and property victimization as well as reduction of psychiatric symptom severity in dual-diagnosis patients | The SOS training comprises three modules: self-wise, other-wise and street-wise - each of which consist of four sessions. Self-wise: involves a newly developed emotion regulation skills training, inspired by the principles of existing emotion regulation skills training. Comprises various interactive group exercises to practice with recognising one's own emotions, interpreting emotional expressions of others and coping with feelings of anxiety and feeling of anger. Other-wise module: Newly developed conflict resolution skills training specifically focusing on preventing and resolving interpersonal conflicts. Participants jointly compose a list of important resolution skills which are practiced in role-playing exercises categorised by relevant themes. The specific role-playing exercises are based on input from dual-diagnosis patients who participated in the pilot phase to ensure the exercises are relevant to the population and applicable to real-life situations of dual-diagnosis patients. Streetwise module - Newly developed street skills training which builds on the idea that teaching patients about behavioural factors that contribute to their risk of victimisation may be effective in reducing victimisation. | **Setting:** Outpatient and inpatient  **Modes and format:** F2F group based  **Frequency and intensity:** 12 twice-weekly 90-minute sessions including a 15-minute break, within a 6-week time frame. Sessions were delivered by any 2 of 14 trained therapists with a maximum of 8 participants per session  **Types of treatment provider involved:** Therapists with a master's level university degree and 8 nurses with immediate or higher vocational education. They all received training to deliver the SOS training which included studying the treatment manual and participating in 2 4-hour training sessions. The first 12 sessions of SOS training per location were supervised and supervision was provided at least once a month at each location. Additional supervision by telephone or email was provided on request. |
| The Victoria intervention | The Netherlands | To help people with a serious mental health condition and severe limitations in social functioning participants safely get involved in social contexts, following a victimisation experience | The intervention has four steps as follows: exploring, analysing, clarifying the context, and future steps. The first step is to explore the victimization experience together in a one-on-one meeting with the client alone. This involves evaluating the client’s satisfaction and activity related to life domains, such as housing, contacts, education, and work. The other steps of the Victoria intervention are indicated when there are signals that a client is avoiding activities or if desired progress on these domains is stagnating. If this is the case, the possible role of recent victimization experiences in this is assessed. The second step discusses the most relevant negative experiences related to social participation. The third step is to clarify the context of the experience. The fourth and final step involves determining future steps based on the results of the previous steps. | **Setting:** Home or F-ACT office  **Modes and format: F2F**  **Frequency and intensity:** The Victoria intervention is personalised: One client may need and want several conversations, while another may be satisfied with one or two conversations. Most conversations last 15-60 minutes.  **Types of treatment provider involved:** Staff from Flexible Assertive Community Treatment (F-ACT) teams, generally consisting of a psychiatrist, employment specialist, psychologist, mental health nurses, and experts-by-experience |
| *Offending models* | | | | |
| Forensic peer support service | USA | To model recovery for the forensic population with a serious mental health condition | Peerstar employed 77 peer specialists who provided peer support services to an active caseload of 429 individuals. All peer specialists self-disclosed as present or former consumers of behavioural health services with a minimum education level of high school diploma or equivalent. They received 80h of peer specialist certification training from one of two training vendors approved by the state. They also receive weekly supervision from individuals who have received state-approved peer support supervisor training. All peer specialists working in the FPSS received an additional 16 h of specialised forensic peer support training. Inmates are paired with peer mentors 30-90 days before release. Inmates are screened for risk of recidivism. An 'Intake and Re-entry transition plan' is developed by the inmate and peer specialist, and implementation of this plans starts before the inmate is released. On release, entering citizens are paired with community-based forensic peer support mentors (who are not typically the same FPS who worked with the individuals inside the jail), and they create an 'Individual Service Plan', which is implemented and then reviewed/monitored every 6 months. This plan contains activities to help the individual meet their recovery goals. an individual is discharged from the FPSP upon request or disengagement, or when he or she has achieved the goals in the ISP and there is a reasonable expectation that discharge from the program will not result in loss of gains or goals attained and that services are not expected to provide additional benefits to the individual. | **Setting:** Prisons, then in the community (exact location not specified)  **Modes and format:** F2F, individual  **Frequency and intensity:** ongoing  **Types of treatment provider involved:** peer supporters |
| Liaison and diversion schemes | England (1), Australia (1) | To improve the early identification of people with a range of vulnerabilities — such as mental health issues and learning disabilities — who come into contact with the youth or adult criminal justice system (CJS). | Liaison and diversion schemes are a model used to divert people with certain needs or vulnerabilities away from the criminal justice system and refer them referred to appropriate interventions or treatment in the healthcare system. 1) the Liaison service becomes involved when a person comes into the custody centre of the magistrates’ court and is referred by the offender's lawyer or by the magistrate 2) after referral the service ensures the offender receives appropriate medication and that other custodial management issues are addressed 3) an assessment is conducted and a report is presented to the court regarding the person's mental health. | **Setting:** NR  **Modes and format:** F2F  **Frequency and intensity:** NR  **Types of treatment provider involved:** NR |
| Mental Health Court | USA (19) Australia (1) Canada (1) | To reduce recidivism, increase compliance with outpatient treatment (and other court-ordered conditions), reduce emergency room visits and reduce hospital time. Also aims to increase public safety and the quality of life of offenders with a serious mental health condition. | The main aspects of mental health courts are as follows:  1) Participation is voluntary with clear terms for participation. Informed choice is a key part of voluntary participation, including evaluation of competency to make an informed choice regarding participation. Mental health courts also aim to identify potential participants in a timely fashion 2) Preliminary assessment in which a clinical advisor (registered psychologist) assesses whether the person understands the program and consents to being involved. A written report is provided to a magistrate who makes the final decision on whether the person is accepted onto the program. Mental health courts are organised so that there is corroboration between agencies involved including both criminal justice and mental health professionals  3) Bi-monthly court reviews aimed at motivating the offender to continue treatment. Complexities of the problems faced determine the length of the program- sufficient time to engage in relevant treatment programs to gain a benefit is required  4) ongoing monitoring to allow treatment plans to be reviewed and updated where necessary. Connection to individualised and inclusive treatment services and support systems within the community is provided, and specially trained teams assist the participant to accomplish established treatment objectives. 5) In the event of non-compliance, sanctions are not used, but court attendance is required where the magistrate encourages the person to engage in the treatment. there is a focus on reintegration rather than judgment and disapproval, however if the program appears unsuitable, participation may be terminated and the person return to the normal court (poor performance or lack of satisfactory progress is not relevant to sentencing). | **Setting:** Community and forensic treatment services  **Modes and format:** F2F individual court appearances  **Frequency and intensity:** Eligible defendants are required to participate in the MHC for varying lengths of time that depend on their level of criminal charge at entry as well as their progress throughout the program, which was based on compliance with terms of participation.  **Types of treatment provider involved:** Various, depending on the mental health court conditions. Also involved are a clinical advisor (usually a registered psychologist) who assesses the suitability of the participant for the mental health court magistrate. The mental health court magistrate decides whether to accept the person into the program and encourages treatment adherence at court appearances. A team of clinical liaison officers are responsible for the implementation of the treatment plans. |
| RESET | UK | To support people with a serious mental health condition to transition into the community and gain secure housing post release from prison. | referrals made by prison InReach team. Pre-release work focused on building rapport, and developing individuals' motivation and engagement with services. RESET team would offer to meet with prisoner on day of release to support them with crucial appointments (e.g. probation/local authority housing). They also ensured correct medication, prescriptions, and planned appointments. Support continued for 12 weeks to help the participant gain safe and appropriate accommodation, access benefits, engage with health services, and strengthen links with family and community services/support (holistic care). RESET workers did not need a professional qualification although all had degrees/prior relevant experience, and all had training to level 4 of IAG training, as well as training in motivational interviewing, housing law, advocacy, and dynamic risk management. | **Setting:** in prison initially, then community settings  **Modes and format:** individual  **Frequency and intensity:** 12 weeks  **Types of treatment provider involved:** RESET team, relevant housing/welfare services |
| Serious and persistent mental illness (SPMI) Release Planning service | USA | To reduce recidivism in those being released from jail with a history of serious mental illness | Delivery of release planning services (which are voluntary), addressing offenders housing, vocational, chemical dependency, psychiatric, disability, medical, medication, and transport needs, with special 'release planners' providing this support rather than case managers. | **Setting:** Correctional facilities  **Modes and format:** F2F, individual  **Frequency and intensity:** meet with release planner several times over the last few months prior to release  **Types of treatment provider involved:** release planner', and then different staff depending on services involved in release planning- e.g. psychiatry |
| Thinking for a Change (T4C) | USA | To address the dynamic, malleable criminogenic risk factors associated with justice-involved adult males with a serious mental health condition | T4C is a highly structured, 25-session, manualized intervention that is delivered in a closed-group format to 8 to 12 people at least twice a week over a 3-month time period. The intervention curriculum includes three modules: (a) social skills training that teaches participants cognitive skills to interpret and respond positively to social situations that involve potential conflict, (b) cognitive restructuring activities that teach participants a concrete process for self-reflection, and (c) a structured problem-solving method that builds on the skills taught in the other two modules to integrate the skills from the modules to teach participants skills to manage interpersonally challenging situations. | **Setting:** County jail or community mental health centre  **Modes and format: F2F, group**  **Frequency and intensity:** 25-sessions delivered in a closed-group format to 8 to 12 people at least twice a week over a 3-month time period.  **Types of treatment provider involved:** Community mental health practitioners, social workers |
| *Multidomain models* | | | | |
| Clubhouses | USA (3), Sweden (1), Australia (1), Canada (1) | To support transition from hospital to community for people with severe mental illness and address social isolation, readjustment to society, and community integration | Membership of clubhouses is voluntary, and members must have a current or past psychiatric diagnosis. Clubhouses emphasise social interaction as an essential aspect for rehabilitation, and clubhouses are intentional communities that emphasise development of social connections that create a psychological sense of community. Clubhouses follow a 'work-ordered' 9-5 workday schedule, and members and staff work together on tasks in operating the clubhouse (e.g. making lunch). There is low staff: member ratio with a non-hierarchical style to promote peer support. | **Setting:** Community  **Modes and format:** F2F, Group  **Frequency and intensity:** ongoing  **Types of treatment provider involved:** Clubhouse staff members trained in recovery practices |
| Community-Enhanced Social Prescribing | UK | To enhance community and individual wellbeing | Each primary care network employs a link worker who is responsible for social prescribing. The link worker works closely with the citizens’ panel to fully understand the assets of the local community, and also supports it and feeds into the mapping process. They utilise a model of social prescribing informed by Connecting People, which requires full and active engagement with the community with whom they are working. Link workers engage with people within primary care settings who are seeking to improve their wellbeing by engaging with local groups, networks, resources, activities or assets. They follow the Connecting People steps of establishing readiness; mapping the individuals’ existing networks and access to local community assets; setting goals for enhancing their wellbeing and planning with which local resources might assist; supporting them to engage with community resources; reviewing with them their progress towards their goals and supporting them to overcome barriers to community engagement. Link workers’ engagement with local people, the citizens’ panel and the wider primary care network gives them an important role in ensuring that the local asset map is a dynamic resource that is kept up to date, is relevant and fit for purpose. As well as using it in their daily work, they will also continually update it and promote its use in the wider community. Working with the citizens’ panel, they will also help to identify gaps in local provision. It is hypothesised that increased knowledge and use of local assets, resources and networks will bring benefits for both individuals and communities. | **Setting:** NR  **Modes and format:** NR  **Frequency and intensity:** NR  **Types of treatment provider involved:** Link workers citizens panel- members of the community |
| EMILIA | UK (1), Finland (1) | To improve the way in which service users with schizophrenia, schizoaffective or bipolar disorder can experience greater participation and inclusion in the delivery of services, training, employment and unpaid meaningful activities, as well as to enable the organization to provide such opportunities. | The EMILIA Project developed and produced 11 training programs as the main intervention to be delivered to the users participating in the project. These training programmes were Dual Diagnosis, Empowering People in Recovery, Family Network Support, Personal Development Plan, Post Traumatic Stress Disorder (PTSD) Intervention, Powerful Voices, Social Competences (work related), Social Network, Strengths Support, Suicide Intervention and User Research Skills. The overall structure of the training programs was similar. Each program started with an introductory phase including a “getting to know each other” or “breaking the ice” exercise. Then an introduction was followed by the main topic of that particular program. This agenda usually filled the first day’s training. The following days’ training had different themes but a similar structure, opening with an introduction to an exercise and detailed instructions to the trainer and finishing with an evaluation. The training programs for the EMILIA intervention were chosen and developed in collaboration with mental health service users and mental health professionals. Each program was of varying length and module based. The minimum requirement of the eight demonstration sites was that they delivered at least three training programs to the participating users. Only a couple of the sites chose to deliver the original programs as they were presented. The majority chose a set of modules, picking from several programs, to tailor them to their own particular local needs. | **Setting:** 7 mental health services and 1 university across 8 European countries  **Modes and format:** F2F, group  **Frequency and intensity:** NR  **Types of treatment provider involved:** Peer support workers (mental health service users as trainers) |
| Horyzons Project | Australia | Improve social and vocational functioning, as well as prevent relapse in individuals with first episode psychosis | Horyzons is based on the moderated online social therapy (MOST) model, which integrates interactive online therapy (“pathways” and “steps”), peer-to-peer online social networking (“the café”), peer moderation, and expert support. Therapy content focused on understanding psychosis, preventing relapse, making connections with others, and managing depression and anxiety | **Setting:** Online  **Modes and format:** Digital platform  **Frequency and intensity:** Logged onto platform either weekly or fortnightly, depending on individual  **Types of treatment provider involved:** registered mental health clinicians |
| Hospital discharge service | England | To help support people as they transition from inpatient care to the community | The support offered may include: Help finding alternative accommodation.  Help maintaining the safety and security of the home. Help in managing finances and benefit claims.   Support with long term tenancy management.  Developing domestic or life skills.  Support to access other appropriate services and support with filling in any required paperwork.  Increasing social engagement and reducing isolation.  The key aim of our HDS is to resolve any housing issues and deescalate any associated mental health distress. Subsequently, reducing pressures on crisis teams and preventing avoidable admissions, delayed discharges and readmissions. The teams also reduce the pressure on existing crisis or ward-based staff to resolve housing issues, freeing up more clinical time. Simply put, we allow clinical teams to do what they do best, whilst we use our expertise as a social landlord to ensure patients have access to safe and secure accommodation. This helps remove any barriers to discharge or providing preventative support to those already living in the community. One of the key elements in the success of our mental health services is the use of our knowledge as a housing association to find solutions for residents with more complex needs. This may include those with dual diagnosis, forensic history or financial arrears. We understand the systems in place and developed partnerships with other housing providers as well as offering our own general needs stock. All of which helps us get the best results for the residents we support. | **Setting:** inpatient wards then community  **Modes and format:** NR  **Frequency and intensity:** NR  **Types of treatment provider involved:** NR |
| Intensive Psychiatric Rehabilitation (IPR) based on Choose-get-keep (CGK) | USA | To help individuals with severe mental illness achieve housing or employment goals | Firstly, participants were aided in choosing a meaningful rehabilitation goal relating to either housing or employment. Then, they were then offered support 2 to 4 times a month while they worked towards their goals. There are 4 phases: assessing and developing readiness for rehabilitation, goal setting (named the “Choosing” phase), goal achievement (named the “Getting” phase), and goal keeping (the “Keeping” phase), with a particular focus on the goal setting phase. | **Setting:** Multiple in-person locations  **Modes and format:** F2F  **Frequency and intensity:** After goal-setting, received support 2-4 times a month, in person  **Types of treatment provider involved:** IPR practitioner |
| Intentional recovery communities | USA | To help improve the psycho-social needs of people with a serious mental health condition | Women’s Empowerment Centre: 15 – 25 members, Open Monday – Friday 10am– 4pm for women only, with activities, staff support and peer support specialists. Residential Community: 13 members, Supported apartments in a converted building, with tenant council and other social activities. Day Program: 20 members, Open Monday – Friday 8.30am–3pm with structured social and learning activities, Adolescent Group: 8 – 12 members Open twice weekly after school and during school holidays with structured and unstructured activities. | **Setting:** Depends on community - day centres or supported apartments  **Modes and format:** F2F, group  **Frequency and intensity:** NR  **Types of treatment provider involved:** NR |
| Look Ahead's mental health pathways | UK | To provide integrated mental health services and housing support | Crisis and recovery houses: - a short-term community-based alternative to in-patient psychiatric treatment. Those who receive support through these services are less likely to be re-admitted to hospital, while the NHS and social care agencies benefit from lower costs per-bed-per-day than in-hospital services. Rehabilitation services: - accommodation-based services providing medium or short-term support to develop mental health stability and daily living skills. Forensic step-down: for people to step-down safely from secure in-patient settings into the community Supporting inpatient discharge: Look Ahead’s Housing and Advice Workers (HAWKs) support mental health patients in acute wards to ensure they have suitable housing options following discharge. Community-based support: The Independent Living Community Support provided in Tower Hamlets currently supports 186 individuals with varying needs to live independently and to maintain their tenancies. | **Setting:** NR  **Modes and format:** NR  **Frequency and intensity:** NR  **Types of treatment provider involved:** NR |
| Mental health day centres | USA | NR | Mental health day centres accept clients by referral only (predominantly from mental health teams). They are open 6 days a week and some evenings, offering patients a drop-in facility, cheap food, and therapeutic groups and members meetings. These groups can differ between sites, e.g. some offer culture specific groups and outreach for patients from ethnic minority groups, some also involve art music and dance therapists. | **Setting:** Community  **Modes and format:** F2F, group or individual  **Frequency and intensity:** at least weekly meetings are encouraged, although the service functions as a drop-in service so patients can use it as many times as needed  **Types of treatment provider involved:** Staff have a wide variety of backgrounds including social services, psychiatric nursing, art, music or dance therapists |
| Mental health inclusion service † | UK | NR | The service offers 3-6 months’ worth of one-to-one support with an inclusion coach, which centres around goal setting and identifying hobbies and interests, and also barriers to these (e.g. living rurally with no transport). Inclusion coaches also deliver mental health drop-in hubs around the county, with support from volunteers, and we have a tailored platform online called Wiltshire Clic. Anyone can use the hubs or the website at any time; you don’t need to be accessing 1-1 support. The service also supports transition into adult mental health support for ages 16+, and there is a close link with CAMHS to support young people leaving their services. Additionally, there are Digital Tech volunteers to support people to learn about getting online and how to access support virtually, and travel training volunteers to support with getting on public transport. These are supported by two members of staff (A digital officer and a peer and volunteer co-ordinator). | **Setting:** Community  **Modes and format:** NR  **Frequency and intensity:** NR  **Types of treatment provider involved:** Digital officer and peer and volunteer coordinator |
| Mental Health Navigation | UK | To help people with mental illness access support for their unmet non-clinical needs | The first appointment with a new service user is an opportunity to get to know them and conduct an assessment on their needs from a whole-person perspective. This sets the tone for the work that they will do with their Mental Health Navigator. Each of the Mental Health Navigators uses a tool that covers eight different topics to guide the initial appointment and subsequent work. The first appointment is a minimum of an hour and a half, with Mental Health Navigators often allowing for two hours in their calendar for this initial meeting. A holistic needs assessment tool is recommended to be undertaken at this point (rethink provide a template form that can be adapted to local populations). The holistic needs assessment tool is used as a starting point for the Mental Health Navigator to understand the unmet non-clinical needs of the service user. Using it as a baseline they work together to set realistic goals and steps to achieve them. The Mental Health Navigator will use their local community knowledge to recommend and support the service user achieve these goals. Each service delivering Mental Health Navigation has formed their own referral pathways. This has been done to ensure that the service aligns appropriately with the healthcare systems and pathways already in place. It allows for flexibility in approach to meet the needs of the local population. It also allows referral pathways to be put in place where there is pressure on healthcare services where people are presenting with unmet social rather than clinical needs. | **Setting:** Navigators setting at the service users’ home or out in the community.  **Modes and format:** F2f or telephone  **Frequency and intensity:** appointments usually last between 30 mins and an hour. The support offered is not time limited, and the length of time supporting a service user can vary depending on the complexity and level of need presented. Support can vary as widely as being provided for a few weeks to six months.  **Types of treatment provider involved:** NR |
| Peer-delivered centres for independent living | USA | To improve empowerment, recovery, quality of life, participation and getting needs met for people with psychiatric disabilities | Participants were contacted by phone by the CPS on 3 different days at 3 different times to schedule the first session. If the CPS was unable to schedule a meeting, a letter was sent to the participant to reiterate the services they were eligible to receive at the CIL. First session - an overview of the philosophy and history of Centres for Independent Living and the score services the CIL offers e.g. information and referral, independent living skills training, peer support and advocacy. Second session - A standardised script was used to assess unmet needs and assist the CPS and participant in identifying where they would like support. The list included 25 areas such as housing, transportation and education and was used as a tool throughout the intervention as the CPS would start each session by checking in on progress within each unmet need that was previously identified. | **Setting:** Community  **Modes and format:** F2F, individual  **Frequency and intensity:** The amount and frequency of sessions was driven by the participant as they were encouraged to work with the CPS.  **Types of treatment provider involved:** Individuals who received certification as a peer specialist (CPS) from a well-known CPS training organisation with a 75-hr training curriculum and performance standards that need to be achieved. The CPS received training and supervision in the CIL core services philosophy and model from peers with disabilities in the organisation |
| Psychologically Informed Environments | UK | To update and improve the flexibility of the principles of the therapeutic community, thereby meeting the psychological and emotional needs of residents. | 5 components of PIEs: 1. A psychological framework, explicitly committed to as the therapeutic approach underlying the project. 2. The physical environment managed in a way which promotes psychological safety. 3. Staff supported to make consistent changes to approaches to clients by means of reflective practice groups, which also provide ongoing learning and reflection. 4. Managing relationships should be considered the principal tool for therapeutic change, rather than controlling behaviour. 5. Outcomes should be evaluated at service and individual levels | **Setting:** A hostel for homeless people  **Modes and format: F2F**  **Frequency and intensity: NR**  **Types of treatment provider involved:** Hostel staff and psychotherapists |
| Recovery Colleges | Australia (2), UK (2) | To assist people with mental health difficulties in their recovery through education, develop life skills and move towards recovery-focussed life goals | Supports people through their individual recovery journey and support transition from dependency--> self-management in the longer term. Through an educational approach, the college uses psychoeducational and vocational training as well as social and peer support tools to promote mental health recovery. examples of courses are those related to ‘Mental Health and Self‐Management’ (e.g. Anxiety Management, Confidence Building), ‘Life Skills’ (e.g. Preparing to Volunteer, Work Preparation), and ‘Creative and Wellbeing’ (e.g. Healthy Living, Writing for Well‐being), exploring emotional resilience, creative wellness, dialectical behaviour therapy skills, understanding the impact of a difficult childhood, and a de-stigmatising mental health art workshop. Course lengths vary. | **Setting:** NR  **Modes and format:** F2F, group  **Frequency and intensity:** Courses varied in the number of hours or sessions, but each course completed within 3 months  **Types of treatment provider involved:** Mental health professionals and lived experience staff/peer support workers |
| Rethink mental illness community services (Brent mental health service) † | UK | NR | Provides support to adults with mental health needs aged 18+, within the Brent borough alongside statutory services. The service has 4 main aspects of delivery which are Befriending service utilising volunteers; Peer Navigators Support; Substance Misuse Support delivered via Change, Grow, Live (CGL) staff; Mental Health and Recovery Workshops. | **Setting:** Community  **Modes and format:** NR  **Frequency and intensity:** NR  **Types of treatment provider involved:** NR |
| Shared Lives | UK | The pilot aimed to reduce the significant challenges young people face when leaving care such as mental health and wellbeing, poor educational outcomes and housing insecurity, and achieve increased independence for young people leaving care. | Shared Lives (formerly Adult Placement) is a regulated form of social care delivered by Shared Lives carers who are trained and approved by a registered Shared Lives scheme. In Shared Lives arrangements, adults (or sometimes young people aged 16 – 17) who need support or accommodation are matched with compatible Shared Lives carers and families who support and include the individual in their family and community life. Support is provided in three types of arrangement: 1) Long-term accommodation and support: an arrangement where the person moves into the home of the Shared Lives carer. These arrangements can be an alternative to residential care, supported accommodation, or living with their own family. This can be a step towards independent living, or could be a longer-term arrangement for individuals who are unlikely to be able to live independently. 2) Short breaks: an arrangement where the person stays in the home of the Shared Lives carer for a limited time. This can be utilised as a form of intermediate care, or for family carer respite. 3) Day support: involving the sharing of home and family (and/or community) life of the Shared Lives carer during the day. This can be used as an alternative to other forms of day support, such as day centres. | **Setting:** community/supported living  **Modes and format:** F2F, individual  **Frequency and intensity:** average of 8.5 months  **Types of treatment provider involved:** supported living staff, Shared Lives carer |
| Social cafes | UK (1), Denmark (1) | The purpose of the cafes is to support people, to provide some consistency for those transitioning from inpatient settings to the community and give them somewhere to meet and socialise with people | The cafes are operated by a paid member of staff supported by trained volunteer recovery coaches. They operate a mix of planned and optional drop-in activities to enable people to access help and resources to support their mental health recovery. The cafes offer advice and support to promote good mental and physical health. A locally based project worker offers project participants the opportunity to challenge their problems and gradually prepare for a job by taking part in the daily work in the café. At the social cafe, people can go and take part in games, gardening, music, learn about self-care and access online resources such as My - Toolkit - an online space to support mental health and wellbeing. Meanwhile, the participants and the project worker together work on reducing or removing other problems standing in the way of improving the participants’ life situation. | **Setting:** Community settings e.g. church, library  **Modes and format:** F2F, group  **Frequency and intensity:** ongoing  **Types of treatment provider involved:** NA |
| Social Welfare Legal services in health settings | UK | To save time and reduce pressure on healthcare services by providing specialist legal advice, and also improving access for those who would otherwise not seek legal advice. | Wide range of services, which include advice Charities (e.g. Citizens Advice), Health charities (e.g. Macmillan), and mental health charities (e.g. Mind), local authorities/health services (welfare rights advice teams), independent services such as law charities, and collaborations of local providers. Some of these, such as Citizens Advice, Hospices, and Macmillan, have a replicable model across the country. They also connected with healthcare services in different ways, such as being completely integrated, direct and indirect links to services, and more of a 'social prescribing' function. | **Setting:** community setting  **Modes and format:** various  **Frequency and intensity:** varied  **Types of treatment provider involved:** various – depending on the service: mental health staff, charity workers, support workers, social workers, or lawyers |
| STAR | UK | To deliver face-to-face support to people with complex needs, including those in rough sleeping hotspots, and to those at risk of losing their tenancy due to anti-social behaviour, poor mental health, or drug and alcohol issues. | NR | **Setting:** NR  **Modes and format:** NR  **Frequency and intensity:** NR  **Types of treatment provider involved:** mental health practitioners, charity workers, housing services |
| The Citizens/Citizenship Project | USA (7), Canada (1) | To address and enhance the key elements of citizenship for people with mental illness and criminal justice records. | Participants are matched with a Peer Mentor, and meet the Program Director before starting. The course shares similarities with social rehabilitation and social skills programs but also embodies an emphasis on both group support and community contacts. Students are treated as individuals with unique strengths and skills who are capable of exercising rights, roles, and responsibilities and developing personal identities as valued members of society. The Project Director, with help from an assistant, Peer Mentors, and community and student presenters, facilitates twice-weekly two-hour classes. Each class, as well as each meeting of the valued role project that follows the course, begins with a “What's up?” discussion. Students talk about their activities during the preceding week, including to their struggles and successes in recovery from behavioural health disorders, efforts to find or maintain housing, educational goals, and other topics. The curriculum includes a wide range of topics (e.g. problem solving, citizenship, anger management) with sessions delivered by internal or external speakers. Following the “What's up?” discussion, students work with a group facilitator to plan and complete an education-focused project in the community that embodies the goal of establishing meaningful social roles for themselves as contributing members of society. The Peer Mentor component offers ongoing mentorship, counselling, and support to students as they make their way through the group component and, sometimes, after they leave the intervention. All peer mentors complete an 8-week training course prior to starting. This intervention has also been examined online- these were similar except they had a fixed duration. | **Setting:** Community venue (local church)/online  **Modes and format:** F2F - group, or online  **Frequency and intensity:** 5-month programme with twice-weekly two-hour classes  **Types of treatment provider involved:** Peer mentors who completed training on confidentiality, the client engagement process, cultural competence, and the distinctive roles of criminal justice and mental health treatment system personnel |

*NR: Not reported. NA: Not applicable. F2F: Face to face. MHC: Mental health court*

### Appendix 4: Evaluation outcomes

#### Appendix 4.1: Housing

| Model | Author ID  (country) N=sample size | Study design | Outcomes | Author stated conclusions regarding model’s usefulness |
| --- | --- | --- | --- | --- |
| Critical time intervention | Herman et al., 2011 (USA)  N= 150 | Quantitative-RCT | At 18 months, the odds ratio of experiencing homelessness in the intervention group compared to the control group during the final three follow up intervals was 0.22 (95% CI:0.06, 0.88) when controlling for baseline homelessness, indicating that the intervention group were less likely to experience homelessness. Among those assigned to Critical Time Intervention there were 1,812 total homeless nights, and among those assigned to the control group, there were 2,403 homeless nights. Poisson regression to control for baseline homelessness showed that this difference was statistically significant at p<.001. | The Critical Time Intervention had a substantial, lasting impact on reducing the risk of recurrent homelessness among persons with severe mental illness after their re-entry to community living |
| Peer-enhanced case management programme | Weissman et al., 2005 (USA)  N= 32 | Quantitative-RCT | No individuals lived independently in their own residence at baseline, 6 (46%) did so at one or more follow-up.  3 (23%) were employed at baseline while 9 (69%) were employed at one or more follow-up (N= 13). Individuals reported no changes for social inclusion, subjective social inclusion, or social acceptance. | Peer case management can be a useful adjunct to standard treatment for homeless mentally ill individuals during the transition to independent housing, but implementation can be challenging. |
| Sheltered housing (supported housing) | Roos 2016 (Norway)  N= 14 | Qualitative-interview study | Interviewees expressed feeling safe in the accommodation, liking the combination of private accommodation and shared social space, some concerns about being able to stay in the accommodation after the 3 year tenancy, some mentioned becoming friends with other residents (different from inpatient stays), taking part in more social and physical activities, some resident conflict (e.g. at weekends when there are less activities), staff treated them with respect and saw them as humans and not patients, and were able to recognise when they were struggling and offer support at an early stage. Most also mentioned liking meeting up with friends outside of the accommodation. Overall, main finding is feelings of safety, high levels of satisfaction in this accommodation. | All residents in the study highlighted the importance of access to the service providers and their skills in observing symptoms at an early stage. They also emphasized the shared accommodation room as important to establish a relationship with other residents. This study indicates private accommodation has many advantages for the residents. Having a short tenancy agreement made some informants feel insecure, but if the goal of the sheltered housing is to help some residents become capable of living in their own self-contained apartment, giving this group a permanent tenancy might be counterproductive |
| Group homes (residential care) | Nelson 1997 (Canada)  N= 107 | Mixed methods- observational pre-post | Residents of Supported Accommodation and Group Homes were more likely to have their own room, to spend less of their income on rent, and to have more control in decision-making in the residences than people living in Board and Care Homes. Residents of group facilities (Group Homes and Board and Care Homes) had more staff support and less emotional abuse than residents of Supported Accommodation. Residents in all three types of housing increased their involvement in work/education and the community. However, none of the groups showed improvement independent functioning, meaningful activity, positive and negative affect, and life satisfaction. | Residents of Supported Accommodation and Group Homes appear to more growth oriented, while the residents of Board and Care Homes appeared to less concerned with change. Group Homes tend to be used to assist people early in their recovery, while Supported Accommodations tend to be used at a later stage after people have made significant growth. |
| Supported accomodation (supported housing) | Nelson 1997 (Canada)  N= 107 | Mixed methods- observational pre-post | Residents of Supported Accommodation and Group Homes were more likely to have their own room, to spend less of their income on rent, and to have more control in decision-making in the residences than people living in Board and Care Homes. Residents of group facilities (Group Homes and Board and Care Homes) had more staff support and less emotional abuse than residents of Supported Accommodation. Residents in all three types of housing increased their involvement in work/education and the community. However, none of the groups showed improvement independent functioning, meaningful activity, positive and negative affect, and life satisfaction. | Residents of Supported Accommodation and Group Homes appear to more growth oriented, while the residents of Board and Care Homes appeared to less concerned with change. Group Homes tend to be used to assist people early in their recovery, while Supported Accommodations tend to be used at a later stage after people have made significant growth. |
| Board and care homes (floating outreach) | Nelson 1997 (Canada)  N= 107 | Mixed methods- observational pre-post | Residents of Supported Accommodation and Group Homes were more likely to have their own room, to spend less of their income on rent, and to have more control in decision-making in the residences than people living in Board and Care Homes. Residents of group facilities (Group Homes and Board and Care Homes) had more staff support and less emotional abuse than residents of Supported Accommodation. Residents in all three types of housing increased their involvement in work/education and the community. However, none of the groups showed improvement independent functioning, meaningful activity, positive and negative affect, and life satisfaction. | Residents of Supported Accommodation and Group Homes appear to more growth oriented, while the residents of Board and Care Homes appeared to less concerned with change. Group Homes tend to be used to assist people early in their recovery, while Supported Accommodations tend to be used at a later stage after people have made significant growth. |
| Los Angeles’ Honeless Oppurtunity Providing Employment (LA’s HOPE) | Burt 2012 (USA)  N= 56 | Quantitative- observational 2-group retrospective | Housing: 50% of participants had lived in permanent supportive housing since enrolment on the programme, compared to 1% of comparators, and took fewer days to move into housing and spent far more days housed (mean+/-SD=79+/-97 days) than comparators (3+/-35 days).  Employment: 57% of participants took part in employment (27% comparators) and were more likely to have worked in a part time or full-time job, less likely to have no employment at all, took fewer days post-enrolment to take part in a job and worked significantly more days in competitive employment that comparators. Half the days worked were in competitive rather than transitional/sheltered employment.  Author stated predictors of outcomes:  Race-ethnicity affected outcomes- African American and Hispanic clients were less likely to have been in supportive housing since enrolment in the programme, took longer to receive placement, and were in permanent supportive housing for less time than White participants. The more time a client spent homeless in the 12 months before enrolment the less likely they were to move into supportive housing, and the less time they spent there. Also, those with substance use disorder were more likely to have lived in supportive housing since enrolment and stayed there longer than those without a co-occurring substance use disorder. | LA's HOPE programme fulfilled its purpose and demonstrated what providing housing and employment support can mean for individuals with severe mental illness who are chronically homeless. This research also adds to the evidence base on the effectiveness of supportive housing and supported employment. |

*RCT: Randomised controlled trial*

#### Appendix 4.2: Money and basic needs

| Model | Author ID  (country) n=sample size | Study design | Outcomes | Author stated conclusions regarding model’s usefulness |
| --- | --- | --- | --- | --- |
| Payeeship/representitive payees | Rosen et al., 2001 (USA)  N= 28 | Mixed methods- interview/survey | Percentages reporting program-related improvement were high for not having people take advantage of the patients by taking their money (63%)  78% reported more stable housing | Patients in money management programs are satisfied with this service and find it to be helpful. For some patients, these feelings of satisfaction are diminished by some feelings of being coerced. |
| Payeeship/representitive payees | Conrad et al., 2006 (USA)  N= 184 | Quantitative- RCT | Six months follow up: After controlling for baseline determination of need, representative payeeship score baseline outcome score, and baseline severity of psychiatric illness score, being in the intervention group was associated with significantly lower scores on the money management measure (B=-0.3, SE=0.1, p=0.02). 12 months follow up: (B=-0.5, SE=0.15, p<0.001). | Even with rather modest implementation, psychiatric care coordinated with representative payeeship was found to be effective in reducing money mismanagement, decreasing alcohol and drug use, and improving quality of life over 12 months. |
| Representitive payee programme | Luchins et al., 1998 (USA)  N= 56 | Quantitative- observational pre-post | During the year of participation in the representative payee program, the mean number of days spent in state hospitals decreased markedly compared with the year before enrolment, from 68 days to seven days. A similar reduction was noted in the number of days spent in state and private hospitals, from 97 days to 15 days | The results suggest that the representative payee program is quite effective in reducing hospital stays |

*RCT: Randomised controlled trial. SE: Standard error*

#### Appendix 4.3: Employment/Education

| Model | Author ID  (country) n=sample size | Study design | Outcomes | Author stated conclusions regarding model’s usefulness |
| --- | --- | --- | --- | --- |
| Social Firms | Morant et al., 2021 (UK)  N= 51 | Qualitative-interview study | A method back into paid employment "I think it’s a starting place for people that have been out of work for some time. It’s a gentle way back into the workplace" | People with mental health problems employed in UK social firms provided generally very positive views of their experiences in these contexts. |
| Social Firms | Svanberg et al., 2010 (UK)  N= 16 | Qualitative-Interview study | Individuals recovering from a mental illness experience social firms as providing a flexible environment which promotes feelings of belonging, success, competence, and individuality through meaningful activities, accepting social groups, and inclusive leadership | Social firms provide a secure base for recovery and are an important model for ‘recovery-oriented services’ |
| Assertive Community Treatment (ACT)/clubhouse | Macias et al., 2006 (USA)  N= 121 | Quantitative-RCT | Outcomes for Assertive Community Treatment (ACT) and clubhouse participants met or exceeded most published outcomes for supported employment programmes. Compared with the clubhouse program, the ACT programme had significantly (p<.05) better service engagement (ACT, 98 percent; clubhouse, 74 percent) and retention (ACT, 79 percent; clubhouse, 58 percent) over 24 months, but there was no significant difference in employment rates (ACT, 64 percent; clubhouse, 47 percent). Compared with ACT participants, clubhouse participants worked significantly longer (median of 199 days versus 98 days) for more total hours (median of 494 hours versus 234 hours) and earned more (median of$3,456 versus $1,252 total earnings). | ACT and certified clubhouses can achieve employment outcomes similar to those of exemplary supported employment teams |
| BRIDGE Supported Education | Gutman et al., 2009 (USA)  N=38 | Quantitative-RCT | The Bridge participants were significantly more likely to successfully enrol in education at 6 months from baseline compared to the treatment as usual (TAU) group- 43% vs 6%. In their Overall Satisfaction Scale responses, participants expressed that the Bridge Program had prepared them for further education and job pursuit by teaching them needed skills and helping them gain the confidence to test their abilities in the school and work environments. | The results support the effectiveness of the Bridge Program and suggest that the program helped participants to increase their skill level in basic academic areas, improve professional behaviours and social skills needed for school and work settings, and gain the confidence to test their skills in the larger community. |
| BRIDGE Supported Education | Schindler & Sauerwald, 2013 (USA)  N=48 | Mixed methods- quantitative one group pre/post-test survey, and post-test qualitative focus group | At the time of starting in the program, five of the 48 participants (10%) reported active employment, seven reported current enrolment in higher education (15%), and 36 reported neither being enrolled in higher education or employed (75%). Upon completion of the follow-up questionnaire 19 of the 48 participants (40%) reported active employment, 11 reported current enrolment in higher education (23%), and 18 reported neither being enrolled in higher education nor employed (37%). Combined employment and higher education totals were 12 at pre-test (25%) and 30 at post-test (63%) an increase of 18 participants (38%). All the participants enrolled in higher education at the start of the program (n=7) were either still enrolled or completed a degree at the time of the post-test. | Quantitative and qualitative results support that the Bridge Program, an occupational therapy program that incorporates principles of supported education and supported employment, can assist participants to achieve these goals. |
| Collaborative Mental Healthcare | Dewa et al., 2009 (Canada)  N= 126 | Quantitative- Quasi-experimental trial | There was a significantly higher proportion of the Collaborative Mental Healthcare group who returned to work (85%, compared with 63%) and a lower proportion that transitioned to long-term disability leave (7%, compared with 31%). In addition, the average number of days on short-term disability leave was significantly shorter for the Collaborative Mental Healthcare group (62 days, compared with 76 days). | Supporting a Collaborative Mental Healthcare model of disability management may be a worthwhile investment. Our research suggests the model in our study was a less costly and more effective way of providing mental health treatments for people who work. |
| Individual Placement and support (IPS)- education focus | Rinaldi et al., 2010 (UK)  N= 166 | Quantitative-quasi-experimental trial | At the start of the intervention, 40% of the clients were working/studying in a mainstream setting but by 6 months, this had risen to 71%. The proportion in open employment rose from 13–35% while the proportion who were unemployed and had no structured daytime activity fell from 27–6%. At 12 months, 75% were working/studying in a mainstream setting. The proportion in open employment rose from 35% at 6 months to 44% while the proportion who were unemployed and had no structured daytime activity remained stable at 6%. At 18 months, 82% were working/studying in a mainstream setting. The proportion in open employment rose from 44% at 12 months to 48% while the proportion who were unemployed and had no structured daytime activity fell from 6% at 12 months to 3%. At 24-months, 76% were working/studying in a mainstream setting. The proportion in open employment remained stable at 48% while the proportion who were unemployed and had no structured daytime activity increased slightly. The clients who were in open employment at baseline all successfully retained employment during the time periods up to 24 months. Only 16% dropped out of education into other structured and unstructured activity. | Our study suggests that the Individual Placement and Support approach adapted to include support to fulfil educational goals appears to have been effective in enabling people with a first episode of psychosis to gain and retain open employment and mainstream education in a UK context. |
| Individual Placement and Support (IPS)- with workplace fundementals module | Nuechterlein et al., 2019/2020 (USA)  N=69 | Quantitative-RCT | Education: Individual Placement and Support patients had a substantially greater likelihood of returning to school during the initial 6 months than control patients [68% (28 of 41) v. 32% (7 of 22), logistic regression correcting for non-significant group differences in school status at baseline, Wald χ2 = 4.64, df = 1, p = 0.03], although the mean duration of periods of schooling was similar between the groups (17.8 weeks, SD = 17.6 vs 14.8 weeks, SD = 9.2), the IPS groups had more school periods (1.7 v 1.0, SD = 1.7 and 1.2, t=2.06, df=57.1, p<0.05). Return to employment: Although the groups did not differ in return to employment at 6 months, at 1 year the Individual Placement and Support showed notable advantages (69% (25/36) v 33% (5/15), p=0.02) | This study demonstrates the substantial benefits of the combined Individual Placement and Support–with workplace fundamentals module treatment for helping first-episode psychosis patients return to competitive work or school and to extend work/school participation over time. |
| Supported Education programmes | Unger et al., 2000 (USA)  N= 124 | Mixed methods- single-armed trial | Education: Twenty-one students (20%) of the 105 students who remained in the study for three years completed programs of study and during the project attained academic certificates or degrees. Of these students, 11 individuals received certificates, 8 individuals received bachelor’s degrees, and 2 individuals received master’s degrees. There was a 90% completion rate of credits students enrolled in. However, 78% dropped out before the end of the 3-year programme (potentially due to external factors e.g., financial constraints).  Employment: 71% of students reported being able to perform their jobs better because of their education and 50% reported that their job fit their education level. | People with psychiatric disorders can attend post-secondary education and complete their courses. |
| Vocational Support Within Early Intervention (VIBE)- IPS | Major et al., 2010 (UK)  N= 114 | Quantitative- quasi-experimental trial | 36% of Camden residents achieved competitive employment, compared with 19% in Islington; the baseline employment rate was 14% in both boroughs. A concurrent drop in unemployment occurred in Camden residents (from 70% at baseline to 43% throughout follow-up). Not all participants categorised as in education during the subsequent 12 months achieved the primary outcome as defined for the study; a minority were engaged in educational activities not leading to a nationally recognised vocational qualification or degree and were therefore not included as having achieved vocational recovery. | Following first episode psychosis, having access to a specialist vocational intervention service predicted vocational recovery during the subsequent 12 months. This effect was demonstrated after adjusting for baseline factors that we knew to be potential confounders. |

*RCT: Randomised controlled trial*

#### Appendix 4.4: Social isolation and Connectedness

| Model | Author ID  (country) n=sample size | Study design | Outcomes | Author stated conclusions regarding model’s usefulness |
| --- | --- | --- | --- | --- |
| Arts based groups | Williams et al 2020  (Australia)  N=48 | Qualitative study-Interview study | The most prominent benefit that participants reported was connecting with the group and other participants. Participants described feeling “part of the group”, or even in the choir “part of a family”. This provided a sense of belonging, where many members had previously felt isolated. Furthermore, over time, many group members developed friendships with others in the group which helped them to overcome isolation outside of the group context. | The thematic analysis demonstrated that participation in the choir and creative writing group activities allowed participants to meet psychological needs of belonging, support, self-efficacy, purpose, and positive emotions. |
| Community Navigator | Frerichs 2020 (UK)  N=19 | Qualitative study-Interview study | Six themes were identified that explained facilitators and challenges to participating in the programme: desire to connect with others; individual social confidence; finding something meaningful to do; the accessibility of resources locally; the timing of the programme; and the participant’s relationship with the Community Navigator. | Factors at the individual, interpersonal and structural level, that enable or hinder an individual’s participation should be identified early, so that people are able to make the best use out of the Community Navigator or other similar programmes |
| Community Navigator | Lloyd-Evans 2020 (UK)  N=40 | Quantitative study-Pilot feasibility trial | Loneliness in the intervention group fell from a median De Jong Gierveld Scale score of 11 at baseline to 9 at follow up, and from 10.5 to 10 for the control group participants. The median perceived social capital (social network resourcefulness) also changed from 7.0 to 7.5 in the intervention group and 11.5 to 11.0 in the control group. | A socially-focused Community Navigators Programme designed to reduce loneliness can be delivered as intended for people with complex depression or anxiety, in secondary mental health services as an addition to standard care. Qualitative feedback and session logs indicate that the programme focused as planned on social relationships. Support from a Community Navigator to review their social world, set goals to develop social connections and then to act on them appeared to help participants to increase knowledge about opportunities for social interaction, access new social groups and activities, and develop or regain positive social identities and connections with others. |
| Compeer | McCorkle 2008 (USA)  N=154 | Quantitative study-Quasi-experimental trial | Compeer clients reported significant improvements in social support and a trend towards improved subjective well-being. After 6 months, social support increased to 13%, increasing to 23% at 12 months. | Compeer is an effective way to increase social support for people with serious mental illness, and findings are consistent with qualitative studies on the topic |
| Compeer | McCorkle 2009 (USA)  N=20 | Qualitative study-Interview study | Participants reported numerous benefits to participating in Compeer. Clients and volunteers spoke enthusiastically about the benefits of gaining a friend. Many intentional relationships deepened over several years into mutually beneficial friendships. Most clients became more outgoing, sociable and active, with increased self-esteem, self-worth and self-confidence. Volunteers who had experienced mental illness themselves provided unique added benefits to the relationship. | Intentional friendships can be an effective and cost-effective way to help people with serious mental illness develop social skills, expand their social networks, and improve their quality of life |
| Connecting People Intervention | Webber et al 2016 (UK)  N=34 | Qualitative study-Focus group | Findings suggest that the intervention leads to an enhancement in the individual’s social network and the service user may experience an increased social confidence and participate in more social activities, which may also improve their well-being. These activities are ideally activities based on shared interests within the local community rather than being confined to health or social care services. The person may also deepen their existing relationships, more closely align their activities to their talents, and increase their own contribution to the lives of others | The Connecting People intervention can achieve outcomes of social connectedness for service users |
| Connecting People Intervention | Webber et al 2019 (UK)  N=155 | Quantitative study-Single arm trial | The Connecting People intervention was associated with participants’ enhanced ability to obtain resources from their networks. The other main finding was an increase in perceived social inclusion in those experiencing high-fidelity Connecting People intervention. | These preliminary results suggest that when fully implemented, the Connecting People intervention can improve social outcomes for people with a mental health problem or learning disability. |
| Connecting People Intervention | Webber et al 2021 (UK)  N=151 | Quantitative study-Quasi-experimental trial | implementation of Connecting People in this study is evaluated as not having shown an effect. | In summary, this study found that outcomes for mental health service users of a practitioner-led implementation of Connecting People did not improve in comparison with teams not implementing the model. Given the pre-defined criteria, there was no statistically significant effect on access to social capital in the implementation group. |
| Creative workshops | Saavedra 2018 (Spain)  N=19 | Qualitative study-Interview Study | Social connection was unanimously described by all interviewees, which has increased through the participation in the creative workshops. The creative workshops are considered to offer a social space for personal knowledge between users and keyworkers outside the clinical contexts where they spend most of their time. Participants explain how, sometimes, the creative workshops in the CAAC have helped them to meet new people and make new friends that they continue to meet and join in activities with outside the museum. | The creative activity we have described enables people diagnosed with serious mental illness to participate in various social practices away from clinical contexts, and helps them to increase the quantity and quality of their social activities. |
| Education groups | Atkinson 1996 (UK)  N=146 | Quantitative study-RCT | Those who attended the educational groups had a significant increase in the total number of contacts (t=2.0, p=0.05). There was no significant increase in contacts in the control group. There was also a significant difference in the mean change between group and non-group attenders (mean change attenders +4, waitlist -1, t=4.4, p=0.0001) and at follow up (mean change attenders +4.5, waitlist -4 t=3.6, p=0.0001). | The improvements in quality of life come mostly from improvements in interpersonal relations and expanding social networks (factor 1) and would seem to be supported by the significant differences shown in the Social Functioning Schedule and the Social Networks Schedule. The social networks data would seem to indicate that the quality of relationships was changing, not just the number of contacts, as group attenders reported significantly more confidants. |
| Groups 4 Health | Haslam et al 2016 (Australia)  N=51 | Quantitative study-Quasi-experimental trial | Significant improvements in loneliness for the intervention group, no changes in the control group | Supporting H1, there was consistent evidence (a) that, relative to a non-treatment control, participation in G4H led to significantly improved mental health, well-being and social connectedness |
| Guided peer support | Castelein 2008 (Netherlands)  N=106 | Quantitative study-RCT | There was no significant difference between groups in the extent of discrepancies between desired and received social support at 8 months (Hedges g adjusted for baseline values: =-0.09, 95% CI: -0.29, 0.47).  Those with higher distress from negative symptoms had significantly less chance of improving on social relations (p = 0.01). | This intervention is effective in improving their social network by encouraging mutual relationships and in enhancing their appraisal support. Contact with peers can play an important role in the prevention of social and emotional isolation. |
| Hearing Voices Network support groups | Beavan 2017 (Australia)  N=29 | Quantitative study-observational pre-post | Social isolation: 89% of participants agreed that their HVG promoted social connection at least quite a lot. | The overall findings of this study are encouraging in that the members who chose to participate reported varied but positive benefits of attendance. |
| Occupation and Social Skills Training | Ercan Doğu 2021 (Turkey)  N=60 | Quantitative study-RCT | The increase in the total score of the Community Integration Questionnaire was found to be higher in the occupational therapy + social skills training group compared to the Social skills training only group (Wilksλ= 0.907,F(2.116) = 3.921;p= 0.027). | The combined Occupational Therapy and Social Skills Training program is a beneficial intervention for people with schizophrenia to address occupational and community needs. |
| Participatory Video | Whitely 2021 (Canada)  N=20 | Qualitative study-Interview study | Almost all participants reported that project involvement gave them a strong sense of connectedness. In fact, many referred to other workgroup members as “family” and “friends” throughout the interviews. Others talked positively about a “sense of camaraderie,” indicating that the project provided helpful peer support and social support | The theme of “connectedness” overlaps considerably with the notion of social recovery, with project involvement leading to new friendships, peer support and a richer social life |
| Peer Specialists | McCarthy 2019 (USA)  N=31 | Qualitative study-Interview study | Case Managers and Peer Specialists commented that the program sessions helped to reduce isolation for Veterans. The unstructured sessions gave the Veterans a chance to have an event to look forward to, and time to talk and interact with the Peer Specialists. | Compared to unstructured peer support and structured curricula delivered by Peer Specialists the program provided a middle level of program structure, and was generally well received by those involved—the Veterans, the Peer Specialists, and the Case Managers |
| Peer Specialists | Rivera 2007 (USA)  N=203 | Quantitative study-RCT | There was no significant difference between peer assisted case management and standard case management in the subjective quality of social relations at either 6-months post baseline or 12 months post baseline, although measures did improve significantly over time | A fairly robust randomized trial of real world treatment delivery with a well-defined role for consumers that capitalizes on their theoretical strengths has not found evidence that consumers enhance case management. |
| Role Development programme | Schindler 2005 (USA)  N=84 | Quantitative study-Quasi-experimental trial | Participants in the experimental group improved significantly in interpersonal skills compared to the control group (p=.020). | Findings suggest that there was statistically significant improvement in task skills, interpersonal skills, and roles among participants involved in the role development programme. |
| Scatter -site & Congregate housing | Yanos 2018 (USA)  N=343 | Quantitative study-Quasi-experimental trial | Participants living in independent scatter site housing with low independent living-skill had significantly lower social community participation than participants in congregate housing with comparable levels of independent living-skill, while the reverse was true for participants with high in-dependent living-skill, who had higher social community participation when living in independent scatter-site housing. Negative symptoms and active coping were the only personal capacity variables which significantly predicted “Social Community Participation” | The degree of community participation demonstrated was moderate, with most participants engaged in social contacts with friends and family members and typically engaged in activities in their communities 1–2 times per week. |
| Social cognition and interaction training | Hasson-Ohayon 2014 (Israel)  N=55 | Quantitative study-RCT | Participants who completed social cognition and interaction training alongside peer mentoring showed significant improvement between baseline and post-assessment in mean scores for social engagement (10.162.3 and 14.362.5, respectively), compared with participants in the control group, whose scores decreased (10.962.1 and 10.862.2, respectively). | The results suggest that participation in the SCIT intervention was beneficial in terms of increasing engagement in social interactions |
| Social network intervention | Terzian 2013 (Italy)  N=357 | Quantitative study-RCT | A social network improvement—defined as an increase in number, frequency, importance, or closeness of relationships—was observed at year 1 follow-up in 25% of the patients allocated to routine treatment and in 39.9% of those allocated to the experimental arm (OR 2.0, 95% CI 1.3 to 3.1; AOR 2.4, 95% CI 1.4 to 3.9). An overall social network improvement—including an improvement in intimate or working relationships—was reported at year 1 for 30.8% and 44.5% of the routine and experimental treatment patients, respectively (OR 1.8, 95% CI 1.2 to 2.8; AOR 2.1, 95% CI 1.3 to 3.4). The results were still statistically significant at year 2 (31.5% in the routine group and 45.5% in the experimental group, OR 1.8, 95% CI 1.1 to 2.8; AOR 2.1, 95% CI 1.3 to 3.5; overall social network improvement 33.3% and 47.9%, respectively, in the 2 groups, OR 1.8, 95% CI 1.2 to 2.9; AOR 2.2, 95% CI 1.3 to 3.5). | The implementation of a social network cannot, and should not, be imagined as a sophisticated technique increasing the burden (and the costs) of care. It would be reasonably practicable if conceived as an opportunity not only to increase the patients’ autonomy but also to decrease the burden of repetition and frustration of the service, thus opening up new opportunities and the expression of new competences. |
| Social Prescribing (secondary MH) | Dayson 2020 (UK)  N=20 | Qualitative study-Interview Study | Interviewees (patients) reported improved emotional wellbeing and quality of life, improved psychological wellbeing (feelings of a sense of purpose in life and opportunities for personal growth, helped to develop confidence leading to self-acceptance, activities providing a 'lifeline' and helping cope with difficult experiences, helped them to make friends and create support networks outside of mental health services, enabled environmental mastery, enabled development of positive relationships with others). They also reported improvements in social wellbeing (developing social contribution, coherence, and integration, and motivation to get out and join a group/activity). | Social prescribing appears to be beneficial for patients in a secondary mental health setting as well as a primary care setting, and shows similar findings in emotional, psychological and social wellbeing. |
| Social recreation programme | Petryshen 2001 (Canada)  N=36 | Quantitative study-Single-arm trial | There were lower levels of loneliness at the one-year follow-up than at intake. People rated their overall social functioning and their satisfaction with social relations higher at the one-year follow-up than at the time of intake.  There were significant associations between previous hospitalisation and improved loneliness, social functioning and satisfaction with social relations. Married individuals improved more than those who were single, but had lower social connectedness scores at intake. | Results from the various outcome measures confirmed a positive impact on people's lives with respect to their loneliness, self-esteem, social functioning, satisfaction with social relations, satisfaction with leisure activities, and general life satisfaction. |
| Supported socialisation | Davidson 2001 (USA)  N=21 | Qualitative study-Interview study | Participants in the supported socialisation conditions reported expanded social networks and reduced social isolation. | The most consistent finding across all of those in­terviewed was the degree to which par­ticipants desired, and responded to opportunities for, friendship. |
| Supported socialisation | Sheridan 2018 (Ireland)  N=70 | Qualitative study-Thematic analysis of diary entries | Achieving belonging and reducing social isolation: volunteer partnered participants reported that having a volunteer partner offered belonging and buffered against the alienation and social exclusion that they had experienced at social events previously & helped them identify increased possibilities to socialise | A key finding was that all participants were willing and more than capable of engaging in social activities irrespective of whether or not they had a volunteer partner to assist them to do so. |
| Supported socialisation | Sheridan 2015 (Ireland)  N=107 | Quantitative study-RCT | Overall social functioning positively changed throughout the three time points from a mean of 99·7 (standard deviation (SD) = 15.1) at baseline, to a mean of 106.0 (SD = 27.0) at the endpoint for the control group, and from a mean of 100·4 (SD = 15.0) at Time 1 for the intervention group, to a mean of 104.1 (SD = 23.4) at the endpoint for the intervention group. No significant differences between groups. | The intervention showed no statistical differences between the control and intervention groups on primary or secondary outcome measures. The stipend and the stipend plus volunteer partner led to an increase in recreational social functioning; a decrease in levels of social loneliness, in depression and in the proportion living within a vulnerable social network. |
| TREE (Toward recovery, empowerment and experiential expertise) recovery programme | Boevink 2016 (Netherlands)  N=163 | Quantitative study-RCT | After 1 year, the patients in the TREE recovery programme did not have significantly lower loneliness scores compared to treatment as usual. | User-developed and user-run recovery programmes may bring about small but reliable changes in recovery and community outcome after two years. More research is required to examine how such programmes can become more successful within the context of disability-focused mental health services. |
| Volunteer befriending programme | Priebe 2020 (UK)  N=124 | Quantitative study-RCT | Patients in the intervention group increased their activity from 20 to 81 min per day in the primary analysis with imputed data, which represents a larger difference than the one that was considered to be clinically meaningful for the sample size calculation. However, a similar increase (from 17 to 70 min) was found in the control group. The analyses comparing the groups at the 6-month follow-up showed that patients in the intervention group still had significantly more social contacts (adjusted difference 0.73, 95% CI 0.05–1.40, P = 0.04), and better scores on the Social Outcomes Index (adjusted difference 3.05, CI 1.13–8.20, P = 0.03). | In this trial of a befriending programme for patients with schizophrenia, time spent in activities increased substantially in both arms but with no differential benefit for befriending. |
| Group volunteer befriending programme | Botero-Rodriguez 2021 (Colombia)  N=23 | Mixed methods-Exploratory non-controlled study | During the intervention the Social Outcomes Index score increased, showing improvement in objective social situation. Improvements were seen in the items on friendships (mean = 1.39; SD = 0.5 to mean = 1.65; SD = 0.49; p=0.05) and as a statistical trend on employment (mean =2.78; SD = 3.07 to mean = 3.45; SD = 3.08; p= 0.10). Qualitative findings also reported an improvement in social isolation: "New, special bonds were formed between the participants. The trust generated within these ties led them to have honest and open conversations about their personal and emotional lives." | Participating in the programme was associated with significant improvements of the objective social situation, whilst we did not find significant changes on scales assessing subjective quality of life, symptom levels and self-stigma |

*RCT: Randomised controlled trial. AOR: Adjusted Odds Ratio*

#### Appendix 4.5: Family and relationships

| Model | Author ID  (country) n=sample size | Study design | Outcomes | Author stated conclusions regarding model’s usefulness |
| --- | --- | --- | --- | --- |
| Integrated Family Treatment | Brunette 2004 (USA)  N= 7 | Quantitative-observational single group pre-post | Of seven parents, one parent lost and did not regain custody of her children but improved on parental skills, another parent who had lost custody of her children improved and regained custody. The only parent who didn’t improve relapsed to alcohol use, was hospitalised, became homeless and lost custody of her children. Details of remaining participants not given. | Integrated family treatment was highly acceptable to parents with severe psychiatric disabilities. Most of the parents, even those with severe psychiatric symptoms, difficult environmental circumstances, and previous neglectful behaviour, improved parenting skills. |
| Mothers and childrens project | Waldo 1987 (USA)  N= 31 | Quantitative- observational | Ten children were in protective custody when the mothers entered the program. After the mothers had attended for 6 months, six of the children were returned to their mothers, 2 were placed in adoptive homes, and 2 remained in protective custody. | Overall, the program has had considerable success in teaching mothering skills to schizophrenic women. |
| Thresholds mothers project | Hanrahan 2005 (USA)  N= 24 | Quantitative- retrospective chart review | At intake, 43 children were living with their mothers. After 6 months, 79% of these children were still living with their mothers and 8 were in foster care. After 1 year, 77% or 27 children remained with their mothers, (excluding children whose mothers left the program).  Literal homelessness declined considerably after enrolment in the program. 6 months after intake, the proportion of families living in shelters for homeless persons declined by 73%. After 12 months in the program, no families were living in shelters (p<0.001), instead families lived in supportive housing, 42% or independent apartment, 58%. It was not possible to determine the residential status of the 17% who had left the project. | The findings suggest that the mothers project benefited both homeless mothers with mental illnesses and their children- there was a relatively high proportion of intact families at 1 year. |

#### Appendix 4.6: Victimisation and exploitation

| Model | Author ID  (country) n=sample size | Study design | Outcomes | Author stated conclusions regarding model’s usefulness |
| --- | --- | --- | --- | --- |
| BRAVE (Better Reduction and Assessment of Violence) | Ruijne et al. 2022  (Netherlands)  N=214 | Quantitative-RCT | There was no increase in detection and referral rates of domestic violence and abuse despite an overall increase in knowledge, attitudes, and skills related to domestic violence and abuse. This suggests that the increase in knowledge, attitudes and skills after the training was not accompanied by changes in clinicians’ behaviour. There was an increase in knowledge, skills and attitudes about domestic violence and abuse, but this did not affect the main outcome (no increased detection). Also, clinicians' readiness to manage domestic violence and abuse was high at all 3 time points, but in the intervention group and control group increased. This increase remained in the intervention group, whereas this increase in readiness stagnated in the control group after 6 months. | The trial showed that a training program on domestic violence and abuse knowledge, attitude, and skills in community mental health teams can increase knowledge, attitudes, and skills toward domestic violence and abuse. However, the intervention does not appear to increase the detection or referral rate of domestic violence and abuse in serious mental illness patients. |
| BRAVE (Better Reduction and Assessment of Violence) | Ruijne et al. 2020  (Netherlands)  N=16 | Qualitative-Focus groups | Despite the fact that all participants who participated in the focus groups found the intervention highly acceptable, topic relevant and important, participants had trouble maintaining the knowledge and skills they had acquired during the domestic violence and abuse training. This indicates that the effects of the intervention were not sustainable. The main reason for this was that their focus on domestic violence and abuse declined in the first few months after the training. They also mentioned possible reasons for not maintaining enquires about domestic violence and abuse, such as fear of disrupting therapeutic alliance, fear of making symptoms worse, knowing when/where to ask, worries about having to breach confidentiality to protect patients and affects on patients' autonomy. Other reasons included fear of safety of patient and professional, and lack of mandate to intervene/frustration when patients do not press charges against perpetrator etc. | The BRAVE intervention was acceptable but not feasible or sustainable. Personal, institutional, and public barriers make it not feasible for CMH professionals to detect domestic violence and abuse in mental healthcare. To increase the detection of domestic violence and abuse, professional standards should be combined with training, feedback sessions with peers and domestic violence and abuse counsellors, and routine enquiry about domestic violence and abuse. |
| Health Pathfinder project | Melendez-Torres et al. 2021  (UK)  N=Not reported | Quantitative-Observational pre-post | First, Health Pathfinder significantly increased the rate of cases discussed. Specifically, we found a 10.9% increase in those sites where Health Pathfinder was implemented, which continued to significantly increase each quarter after implementation by 10.1%. Second, Health Pathfinder projects improved detection of domestic violence and abuse across a wider spectrum of risk. Further evidence of an improved ‘whole health response’ is provided from data indicating a substantial, additional number of Health Pathfinder contacts with victim-survivors who were not yet ready to progress with linkage into specialist services, as well as the provision of specialist advice to health professionals regarding the management of domestic violence and abuse. | Health Pathfinder provided a safe context for people, mostly women, to disclose experiences of domestic violence and abuse, resulting in sensitive professional responses, and access to timely support from specialist agencies. In short, the research showed that Health Pathfinder helped more victim-survivors to safety, and sooner. |
| LARA (Linking Abuse and Recovery through Advocacy) | Trevillion et al. 2014  (UK) N=63 | Quantitative-Quasi-experimental | At follow up, clinicians reported improved knowledge, attitudes and behaviours towards domestic violence. Service users at follow up reported reductions in total violence on both the quantitative measure and in qualitative interviews. Service users described how DV advisors supported them to take actions to reduce their risk of harm. They also reported a reduction in the amount of unmet needs (e.g. childcare, finance). | This study provides preliminary evidence that multi-faceted domestic violence intervention in community mental health settings may be effective in improving services user outcomes. |
| LINKS project | Safe Lives  (UK)  N=Not reported | Quantitative-Observational pre-post | There was a 660% increase in referrals (5-38 within 2016/2017 year), after staff training evaluation forms filled in by staff suggested there was an increase in understanding about how to record safeguarding concerns, staff felt more confident talking about domestic abuse, and their knowledge of referral pathways increased. These improvements meant improved outcomes for the victims of domestic abuse. They also reported improved knowledge in referrals for perpetrators of abuse. In interviews, staff found referring cases to the idva helpful, that the training was useful, and discussed that the training and role would help to save lives, and the importance of remembering to ask about domestic violence. Despite positive feedback, it was felt that movement was slow, lots of staff did not take up the training due to staff restructure and other issues. | The evaluation concluded that the intervention was successful and recommended a longer multi-site evaluation of the project. Other recommendations include ensuring that staff training in domestic abuse is mandatory, reviewing domestic abuse policies and clarifying referral pathways, ensuring clinical supervision, providing top-up domestic abuse training for staff, and clear advertisement of the service. |
| Self-wise, Other-wise, Streetwise (SOS) training | de Waal 2019  (Netherlands)  N=250 | Quantitative-RCT | SOS training was more effective in preventing victimisation than care as usual alone but the results were inconclusive: significantly more participants in the experimental group (67.6%) achieved treatment response for total victimization compared to the control group (54%) at 14 months post baseline, a significant difference (OR= 1.78, 95% CI: 1.02-3.11, P=0.042), however, when examining only violent victimisation, the difference did not reach statistical significance (OR= 1.75, 95% CI: 0.91-3.34, P=0.092).  None of the tested baseline characteristics were significant predictors of treatment response for total victimization. Regarding violent victimization, significantly more participants with a middle or high level of education achieved treatment response compared to participants with a lower level of education [82.6% in middle/high versus 67.3% in lower; OR = 2.46; 95% CI= 1.26–4.83; P = 0.009]. | In dual-diagnosis patients, adding SOS training to usual care is more effective in preventing victimization compared to usual care alone. SOS training can be implemented in addiction–psychiatry services in order to prevent future victimization in these patients. |
| The Victoria Intervention | Albers 2021 (Netherlands)  N= 400 | Quantitative- RCT | No intervention effect was found. No effect for social functioning or victimisation. Also, no intervention effect for overcoming stigmatisation, avoiding social participation, or perceived unsafety. | The Victoria Intervention had no effects on victimisation and societal participation. However, there were indications that the intervention was successful in moderating experienced discrimination. Small but significant effects were also found on the feelings of acknowledgment and recovery support. These findings may reduce the reluctance to discuss victimisation experience that hampers the client’s social participation. |

*RCT: Randomised controlled trial.*

#### Appendix 4.7: Offending

| Model | Author ID  (country) n=sample size | Study design | Outcomes | Author stated conclusions regarding model’s usefulness |
| --- | --- | --- | --- | --- |
| Liaison and Diversion Schemes | Disley et al., 2021  (UK)  N=36,491 | Quantitative Evaluation | The likelihood of service users receiving a custodial sentence is halved following involvement with Liaison and Diversion services. The Liaison and Diversion scheme may reduce the proportion of offences resulting in custodial sentences and thus increase diversion from the criminal justice system. | Not reported |
| Serious and persistent mental illness (SPMI) Release Planning service | Duwe et al., 2015  (USA)  N=1,325 | Quantitative-Quasi-randomised trial | The release planning group had a slightly lower rates of rearrests (66%) and reincarceration for new offence (28%) than comparators (67%, 29%). Comparators had slightly lower rates of reconviction (53%) compared to release planning group (54%). This provides little support for the theory that release planning reduces recidivism. | There is very little difference between the release planning group and comparator group on rates of recidivism, and providing offenders with release planning did not significantly reduce recidivism. |
| MHC | Eckburg et al., 2006  (USA)  N=272 | Quantitative-observational pre-post | 16 (8%) had at least one new conviction compared to 35 (18%) prior to MHC. (p<0.01), 44 (23%) out of 191 disposed defendants had a new charge in 4 months, compared with 70 (37%) who had at least one prior charge in the 4 months preceding their start date. | The preliminary data suggest that MHCs are solving problems for this population of defendants. The fact that the number of new offenses for these defendants is half of what it was prior to the start of MHC tells us that we are meeting one of our goals. In addition, the fact that two-thirds of MHC defendants are currently in compliance with court orders is also a promising finding |
| MHC | Canada et al., 2020  (USA)  N=35 | Qualitative-interview study | Half of participants described the MHC as a 'second chance' or 'get out of jail free card'. Some said it felt like a chance to learn responsibility and skills such as taking part in society and self-care that they wouldn't get to in jail. Second chance for people with mental illness as they can be seen as someone who needs help, rather than punishment. Some chose MHC to stay out of prison, but then saw how it could help them with other needs (e.g. treatment) | As interventions like MHCs continue to expand and utilize diminishing mental health resources, it is important to understand how people enter this system, if bias exists in referral, and how the population currently served by MHCs compares to the population of people who may be eligible but never referred or accepted |
| MHC | O’Keefe 2006  (USA)  N=262 | Quantitative-observational pre-post | 27% had been arrested at least once in last 12 months prior to the MHC, whereas only 6 participants re-offended during the first 12 months of the MHC (statistically non-significant.). | During the first 28 months of operations, the Brooklyn MHC achieved its implementation goals of improving the court system’s ability to identify, assess, and monitor offenders with mental illness; and using the authority of the court to link offenders with mental illness to appropriate mental health treatment services |
| MHC | Steadman et al., 2011  (USA)  N=1047 | Quantitative-two-group prospective | In the 18 months following treatment, defined as entry into MHC, the MHC group has a lower annualized rearrest rate, fewer post–18-month arrests, and fewer post–18-month incarceration days than the treatment as usual group. The MHC graduates had lower rearrest rates than participants whose participation was terminated both during the MHC supervision and after supervision ended. Factors associated with better outcomes among the participants include lower pre–18-month arrests and incarceration days, treatment at baseline, not using illegal substances, and a diagnosis of bipolar disorder rather than schizophrenia or depression. | MHCs meet the public safety objectives of lowering posttreatment arrest rates and days of incarceration. |
| MHC | Ray 2014  (USA)  N=449 | Quantitative-two-group retrospective | There was a significant difference between those defendants who completed the MHC and those who did not. Non-completers were almost twice as likely to have recidivated (74.5% vs. 39.6%, χ2 = 59.03, p b .001) and to recidivate with a felony arrest (68.4% vs. 31.6%, χ2 = 12.93, p b .001) than those who completed the MHC. Among non-completers, 52.7% of those who recidivated did so in year one compared to only 20.0% of the completers. Non-completers recidivated significantly sooner than completers (17.15 months and 12.27 months respectively, t = 2.67, p< .01). | this study adds to the accumulating evidence on the effectiveness of MHCs in reducing recidivism among offenders with mental illness by looking at the long-term and continued impact of a MHC post-exit. |
| MHC | Bagwell et 2013 [Thesis]  (USA)  N=610 | Quantitative-Quasi-randomised trial | Participants who completed the MHC were less likely to be rearrested in the future compared to traditional court participants (χ² (1) 47.003, p <.001). However, participants who started but did not complete the MHC had a higher percentage of rearrests than traditional court participants. | The emergence of MHCs offers a new perspective for the management of mentally ill offenders based in therapeutic justice and not punitive punishment. This study found that the use of an integrated approach of collaboration and accountability between mental health services and the Riverside County criminal justice system demonstrated a higher rate of success in preventing participants’ re-arrest. The process of recovery is continuous and complex. Individuals accepted may have idiosyncratic or non-recovery-oriented reasons for participating, and may also encounter situational factors that make it difficult to enter or graduate from MHC. |
| MHC | Neiswender 2005 [Thesis]  (USA)  N=194 | Quantitative-Quasi-randomised trial | MHC participants were significantly less likely to spend time in jail during the first year as well as the second year of the program than the opt outs when controlling for age, race, and whether or not the current offense was drug related. | The research efforts of this study point to MHC having a significant impact on the recidivism rates of mentally ill defendants |
| MHC | Roman 2011  [Thesis] (USA)  N=89 | Quantitative-Quasi-randomised trial | After controlling for the number of convictions prior to MHC, there was a statistically significant difference on the number of convictions after the MHC- traditional court participants had significantly more convictions than MHC participants (F ( 1, 83) = 4.32, p = .041, n2 = .05). However, there were no significantly differences between the severity of offenses between the two groups. (Mann Whitney U=606.50, z=-0.168, p=.867) | MHCs reduce recidivism and severity over time with respect to its population. In addition, when compared to a similar group of mentally ill offenders, MHCs reduce recidivism. |
| MHC | Lindhorts et al., 2020  (USA)  N=1465 | Quantitative-Quasi-randomised trial | Defendants who completed the MHC supervision had a lower rate of rearrest (15%) compared to those who did not completed the course (53.7%) and eligible defendants who did not participate (55.9%). | Defendants who assaulted family members and had a positive program termination had a much lower rate of rearrest post-program completion compared with those who did not complete the program or did not participate despite being eligible, a finding that held when controlling for other factors. |
| MHC | Campbell et al., 2015  (Canada)  N=196 | Quantitative-Quasi-randomised trial | There was a tendency for completers to have the lowest rate of recidivism (28.6%) relative to partial-completers (50%) and nonstarters (32.6%), but these differences were not statistically significant,2(2,N178)5.22,p.074, | Small but significant improvements were found for criminogenic needs and some indicators of mental health recovery for MHC completers relative to participants who were prematurely discharged or referred but not admitted to the program |
| MHC | Trupin & Richards 2003  (USA)  N not available | Quantitative-observational pre-post | Statistically significant evidence from both courts suggested impacts on relevant criminal justice and mental health indicators of effectiveness. In most cases, the measured effects were in the direction that would be expected for programs intended to reduce crime and criminal justice sanctions while increasing treatment for the mentally ill. Some of the evidence was suggestive of an important prevention role, or at least negative predictive function, for participation in MHCs, in that individuals who chose not to participate fared worse after MHC referral, not only compared to those who did participate, but when compared to their own pre-referral history. | Although causality cannot be clearly attributed from studies of this type, we find it more reasonable than not to conclude that both MHCs described here made significant impacts on both the participants and non-participants referred to them. Both interviewees and quantitative data indicators point to a criminal justice system in that has been positively impacted by a new ecological presence, the MHC. |
| MHC | McNiel & Binder 2007  (USA)  N=170 | Quantitative-two-group retrospective | MHC participation predicted a longer time to any new charge and longer time to a new violent charge. Additional analyses showed that persons who graduated from the MHC program maintained reduced recidivism after they were no longer under supervision of the court, in contrast to comparable persons who received treatment as usual. | These results support the effectiveness of a MHC in reducing the involvement of persons with mental disorders in the criminal justice system. |
| MHC | Lowder et al., 2016  (USA)  N=98 | Quantitative-two-group retrospective | MHC participants had fewer jail days, but not charges or convictions, in the 1-year period following exit from the MHC relative to TAU offenders. MHC graduates, specifically, had even fewer jail days and significantly fewer convictions relative to TAU participants. Finally, we found a significant interaction between time in program and changes in pre- and post-program jail days. Specifically, each day spent in the MHC was associated with a 1.01 times greater decrease in jail days served from pre-program to post-program, even after controlling for whether or not a participant graduated from the MHC. | This study adds to the growing body of literature suggesting MHCs are effective, but also that effectiveness varies as a function of individual and MHC-specific factors. To the extent that evidence-based factors associated with reduced recidivism can be implemented, MHCs may better achieve the ultimate goal of reducing criminal justice contact among adults with mental illnesses. |
| MHC | Hidday et al., 2016; Hidday 2013  (USA)  N=1,095 | Quantitative-Quasi-randomised trial | MHC participants were significantly less likely than control participants to be rearrested in the 6 month follow-up period (38% versus 48% ;x2=10.99, df=1, p=.001) and the year follow-up period than persons in the control group (27.5% vs. 37.3%; 𝑥2 = 11.04,p <.001, Cramer’s V = 0.10). | This study adds to the accumulating evidence that MHCs meet their major goal, reduction of criminal recidivism, and that reductions can extend beyond court exit and may result from more than the provision of treatment and services. |
| MHC | Han & Redlich 2016; Han 2020  (USA)  N=741 | Quantitative-Quasi-randomised trial | Both groups improved in the post enrolment period, such that their arrests decreased. After the analysis controlled for a priori time and group differences, a between-group difference was not found.  MHC participants received more mental health and substance abuse services of all types (except substance abuse case management) compared to comparators, but lived in more disadvantaged neighbourhoods than comparators. However, MHC participants receiving substance abuse related treatments were more likely to recidivate compared to comparators. It was also found that those MHC participants who were living in more disadvantaged neighbourhoods were more likely to recidivise compared to those living in less disadvantaged neighbourhoods. | A key finding of this study was the significant impact of mental health treatment on arrests for MHC participants only.  Mechanisms of success regarding recidivism through MHC are unclear due to involvement of many different services. MHC ppts had significant improvements compared to comparators for variety of behavioural health services. Understanding factors related to recidivism for offenders with MI can help more efficiently target research. |
| RESET | MacInnes et al., 2021 (UK)  N= 62 | Quantitative- observational 2-group prospective | The RESET group were significantly more likely to have secure housing at all timepoints and housed longer than comparison, and more RESET participants were in independent housing.  RESET group had significantly greater level of contact with GPs, more were in receipt of benefits, and in contact with mental health services. Few of either group had entered employment/education. RESET group had greater level of engagement and none in RESET group reoffended in the timeframe, compared to 4 in the comparison group. This was significantly lower (*χ*2 4.42(1) *p* = 0.04). Although not significant, less reoffending was also reported at the following two time points (five at 3 months and seven at 9 months) compared to the comparison group (six at 3 months and nine at 9 months). | The RESET intervention was successful in achieving its main objective, accommodating participants in permanent housing, and reducing the likelihood of homelessness. There is also support for the view that secure housing is important in ensuring a positive transition from prison to the community for prisoners with mental health needs with those receiving RESET services significantly more likely to be in contact with other services (for the receipt of state benefits, accessing GPs and engaging with mental health services), having a lower rate of reoffending and a greater level of service engagement. |

*MHC: Mental Health Court*

#### Appendix 4.8: Multidomain and citizenship

| Model | Author ID  (country) n=sample size | Study design | Outcomes | Author stated conclusions regarding model’s usefulness |
| --- | --- | --- | --- | --- |
| EMILIA | Ramon 2011 (UK)  N=27 | Qualitative- interview study | Users gained new skills and knowledge and were thus empowered to train others by reaffirming their own skills. After 10 months an improved social life was clearly visible among the interviewees. However, difficulties in social relationships, such as lack of confidence or problems with family relationships, continued to exist among the users. The findings at time 2 indicated that overall, there was a slight increase in the number of those who entered employment, from eight at time 1 (33%) to 10 at time 2 (48%). At time 2 being employed remained one of the users’ main goals yet to be achieved by the majority. Key relationships remained a central unresolved issue.  The respondents reported having scarce financial resources due to unemployment, a situation that did not change over the 10-month period. | Although the project has another 25 months to continue from the 10-month follow-up, it has already demonstrated worthwhile achievements of its main aims in relation to the service users who continue to participate in its activities. The high degree of similarity of the findings across the sites, regardless of the socioeconomic and cultural differences that exist between them, indicates the viability of the strategy used in the EMILIA project towards enhancing social inclusion, as well as the validity of the qualitative measurements used. |
| EMILIA | Nieminen 2012 (UK)  N= 23 | Qualitative- interview study | Mental health service users perceived work as an important factor in social inclusion and well-being already at the beginning of the project. The findings show that involvement in work and meaningful activities increased during the EMILIA training. Employment opportunities improved for several participants at the 10-month follow-up point, although without further improvement taking place at the 20-month point. Attending a course prevented social isolation. The project gave the needed confidence for this purpose, along with the creation of one’s own social group. EMILIA helped people to meet, and these social contacts were seen as beneficial. However, some of the users had only a few friends, and difficulties in family relations continued. Maintaining social relationships was perceived to be difficult. There was an improvement in the key informants’ financial state at the end of the project, but financial support was still needed for everyday life. Users at one site were more satisfied with their housing at the end of the project compared to the beginning. Otherwise, the results show no clear changes in the housing situation. Authors reported that social support helped users to achieve goals, find and keep a job, to function, to develop, and to be part of the community | The EMILIA project improved the mental health service users’ social inclusion and employment. |
| Intensive Psychiatric Rehabilitation (IPR) based on choose-get-keep (CGK) | Anthony 2014 (USA)  N= 238 | Quantitative- observational pre-post | Residential functioning for those who set residential goals increased significantly, independent of level of participation. The intervention enrolees group began with a mean residential rating of 4.59 (±1.19) suggesting that they were residing, on average, in supervised facilities, or in other supervised settings. At endpoint, they were more likely to be residing in supervised non-facilities or assisted/supported living environments with a mean of 5.32 (±1.40). The 18-month study completers with residential goals also changed significantly and positively on employment status with their mean score being 1.96 (±1.52) at baseline com-pared with a mean of 2.60 (±1.91) at endpoint. For people who had set employment goals, only graduates showed significant changes in employment status going from a mean score of 2.74 at baseline (±1.94) to a mean of 4.21 (±1.96) at endpoint. Furthermore, employment functioning changed significantly when a goal was set in the residential area; in contrast, residential functioning did not change significantly when goals were set in the employment area. | The present study reiterates the need for rehabilitation counselling staff to be trained in goal setting to improve quality, frequency, and achievement of vocational and residential goals. It also underscores the benefit of goal achievement in one area (i.e., residential) to promote goal outcomes in another area (i.e., employment). Therefore, rehabilitation counsellors might consider setting goals with clients in multiple domains to enhance goal outcomes |
| Intentional Recovery Communities | Whitley et al., 2008 (USA)  N= 38 | Qualitative-focus groups | It appeared that the ongoing constructive social interactions in the recovery community were prompting better coping strategies, a positive change in personal values and an increasingly optimistic view of other people, the future and life in general | Indeed, participants seemed to frame their recovery in these wider social terms: they were recovering not only from a mental illness (a biological disease), but also from baseline troubles of a social and cultural nature. The intentional recovery community thus appeared to positively diminish the loneliness and despair so often identified as concomitant with serious mental illness |
| Look Ahead | Europe Economics 2021(UK)  N= unknown | Quantitative-other (report) | Supported housing helps prevent readmission and has positive outcomes for patients –These services not only help to prevent readmission to hospital or clinical settings, but also help individuals achieve further positive outcomes, for example living independently, securing work, or training, or reconnecting with family or friends. | This report illustrates both the economic and the social care cases for an integrated model of mental health and supported housing services. As the government develops social care reform, Look Ahead believes that integrated mental health pathways, working alongside supported housing, should be a key component. |
| Mental Health Day Centres | Catty 2001 (USA)  N= 109 | Qualitative- survey | The most attractive aspects were the company (59.6%), food (13.8%) and the groups (11.9%).  Negative social aspects- tension or aggression- were mentioned by 11% as their least favourite aspect.  Many called their centre "a lifeline" and reported that it "makes [them] feel wanted" | Not reported |
| Psychologically Informed Environments | Phipps et al., 2017 (UK)  N= 24 | Qualitative- interview study | Most participants spoke about making the hostel a 'home' and the importance of safety. | The data suggest Psychologically Informed Environments are broadly meeting their aims by supporting staff to promote positive experiences of relationships in an environment valued by residents. Nevertheless, this is a challenging task and translating the model into practice is not straightforward. |
| Mental Health Navigation | Mental Health UK (UK)  N= N/A | Online report | One case study reported mental health navigation helped someone develop a social network and reduce social isolation. There is a huge demand for this service across all four pilot areas. This has meant that a key role for the management team is supporting the Mental Health Navigators with their capacity. In some cases, this has been done by setting clear geographical boundaries for the service, such as only taking referrals from certain GP practices. In other areas the delivery teams have not been able to advertise the service to as many clinicians as they had initially planned, as the Mental Health Navigator has consistently reached capacity with only a few clinicians being aware of the service in the area. | Not reported |
| Shared Lives | Shared Lives website, 2022 (UK)  N= Not reported | Mixed methods- interview study | Shared lived reported to provide stability during housing transition, monitoring data suggested little improvement in education, employment, or training, but individual responses show improvement for 10 young people, either increased engagement with voluntary roles, or improvements in education. All were in education or training who responded to the survey, and evaluation reports were positive about effect of support from Shared lives. Several young people showed improvements in relation to integration with family and with the community, involvement in community activities and improved friendships and social networks, and carers helped to support with difficult relationships.  Interviews also indicated increased independence and quality of life. | Carers facilitated greater community connection, as well as relationships with their own and the young people’s birth families. Young people reported that this enhanced their sense of belonging. |
| Clubhouses | Pernice-Duca 2008 (USA)  N= 221 | Qualitative- interview study | Staff and members reported benefits related to community and consistency, participation and opportunity, and respect and autonomy | The study revealed that recovery practices were implemented by using social environment to facilitate a sense of community, participation, and respect. |
| Clubhouses | Hultqvist 2018 (Sweden)  N= 185 | Qualitative- focus groups and questionnaires | Clubhouses promote closeness through work, repeated interaction between members, a non-judgmental environment, evening and weekend activities, social skill enhancement, sharing of similar experiences, equal power between staff and members, flexibly structured activity, and staff outreach after absence. | Clubhouses are suited for supporting their users in maintaining quality of life. |
| Clubhouses | Raeburn 2016 (Australia)  N= 18 | Qualitative- observation and interviews | Found that clubhouses were involved in the reduction of stigma, reduced isolation, and enhanced sense of belonging, and acquiring skills. In addition, feelings of personhood and feeling better and at peace. | The study provides insight into the clubhouse model of recovery and fills the gap in the literature on the processes and outcomes that occur for individuals who utilise this type of psychosocial rehabilitation program. |
| Clubhouses | Prince et al., 2018 (USA)  N= 20 | Qualitative- focus groups | Six themes were identified that explained facilitators and challenges to participating in the programme: desire to connect with others; individual social confidence; finding something meaningful to do; the accessibility of resources locally; the timing of the programme; and the participant’s relationship with the Community Navigator. | Factors at the individual, interpersonal and structural level, that enable or hinder an individual’s participation should be identified early, so that people are able to make the best use out of the Community Navigator or other similar programmes |
| Clubhouses | Rouse 2017 (Canada)  N= 39 | Mixed- pilot feasibility trial, group interviews and focus groups | Loneliness in the intervention group fell from a median De Jong Gierveld Scale score of 11 at baseline to 9 at follow up, and from 10.5 to 10 for the control group participants. The median perceived social capital (social network resourcefulness) also changed from 7.0 to 7.5 in the intervention group and 11.5 to 11.0 in the control group. | A socially focused Community Navigators Programme designed to reduce loneliness can be delivered as intended for people with complex depression or anxiety, in secondary mental health services as an addition to standard care. |
| Clubhouses | Macais et al. 2006 (USA)  N= 121 | Quantitative- RCT | Outcomes for Assertive Community Treatment (ACT) and clubhouse participants met or exceeded most published outcomes for supported employment programmes. Compared with the clubhouse program, the ACT programme had significantly (p<.05) better service engagement (ACT, 98 percent; clubhouse, 74 percent) and retention (ACT, 79 percent; clubhouse, 58 percent) over 24 months, but there was no significant difference in employment rates (ACT, 64 percent; clubhouse, 47 percent). Compared with ACT participants, clubhouse participants worked significantly longer (median of 199 days versus 98 days) for more total hours (median of 494 hours versus 234 hours) and earned more (median of $3,456 versus $1,252 total earnings). | Certified clubhouses (with ACT) can achieve employment outcomes similar to those of exemplary supported employment teams |
| Recovery Camp | Moxham 2017 (Australia)  N= 27 | Mixed methods- qualitative survey | The theme of connectedness was prominent within the dataset. The goals such as ‘make new friends’ and ‘meet new people’ were attained. A small portion of goals within this theme were ranked as being only partially attained and these were related to making a new friend. | Various goals, although individually identified, were commonly shared and many were attained. Therapeutic recreation activities embedded within the Recovery Camp program were the catalyst for individuals to connect with others and enhance their own strengths. A sense of connectedness and the identification of healthy habits—in both the physical and mental domain—were developed. |
| Recovery College | Wilson 2019 (UK)  N=25/19/17 | Mixed methods- observational pre-post | Social inclusion- social inclusion scores increased from baseline to 3-month follow up (non-significant), but a significant increase at 6-month follow-up, particularly on social relations subscale. Qualitative focus group- 10 out of 11 participants said social relationships had improved at 3 month follow up. Additional comments included gaining confidence in meeting others and being around people and speaking in groups and socialising more. Noted increased social inclusion/peer support, new friendships, and a sense of belonging/understanding. | In summary, attendance at the South East Essex Recovery College coincides with significant improvements to mental well‐being and social inclusion, with attenders enjoying their courses and making future plans as a result of attending (such as volunteering or obtaining paid employment). |
| Recovery College | Ebrahim et al., 2018 (UK)  N= 89 | Mixed methods- qualitative survey | Students reported feeling less alone, making friends and being able to meet others with similar problems. Feedback around the theme of social support included: “kept my mind busy, made friends”, “I have found that there are other like-minded people out there”, “I don’t feel so isolated”. Students felt the Recovery College gave them an opportunity to develop their social skills, e.g., “It helped me to appreciate the views of others”.  Students felt inspired to take up volunteering opportunities or find paid work. Feedback around the theme of work/volunteering included: “I am going forward as a paid peer support worker”, “The possibility of becoming a volunteer at the college came up and seems like something I could do”. Students felt the Recovery College helped by encouraging them to leave the house and to feel a part of something. | Findings suggest improvements in wellbeing and personal resources as a result of recovery college courses. Recovery Colleges are seen as unique accepting and enabling. There is a need for further evaluation of the unique contribution of Recovery Colleges to mental wellbeing and the mechanisms involved in promoting the process of recovery. |
| Recovery College | Hall 2018 (Australia)  N= 51 | Mixed methods- document review, interviews, survey and focus groups | The most reported impact for students was in education and learning. The third most common area of impact reported by students was on employment. Several students had taken up the opportunity to participate in the formulation and facilitation of college courses either on a paid or voluntary basis. Other students reported learning about career options such as peer support positions. The college also provides opportunities for social interaction as it reduces isolation. | The impacts discussed by students demonstrate positive outcomes in relation to achievement of educational qualifications, employment, lifestyle and self-care, and social engagement within the community |
| Horyzons Project | Alvarez-Jimanez 2021 (Australia)  N= 170 | Quantitative- RCT | Social functioning remained high and stable in both groups from baseline to 18-month follow-up, with no evidence of significant between-group differences. Participants in the Horyzons group had a 5.5 times greater increase in their odds to find employment or enrol in education compared with those in treatment as usual, with evidence of a dose-response effect. Moreover, participants in treatment as usual were twice as likely to visit emergency services compared to those in the Horyzons group (39% vs 19%). There was a non-significant trend for lower hospitalizations due to psychosis in the Horyzons group vs. treatment as usual (13% vs 27%) | Although no significant effect of Horyzons on social functioning compared with TAU was found, the intervention was effective in improving vocational or educational attainment, a core component of social recovery, and in reducing usage of hospital emergency services, a key aim of specialized first-episode psychosis services |
| Peer-delivered centres for independent living | Salzer 2016 (USA)  N=99 | Quantitative-RCT | Both groups reporting a decrease in the number of unmet needs over time, however the community independent living group showed no differences over time compared to the control (F(2, 172)=1.60, p=0.21). The intervention group had significantly more participation days than the control group at 12 months (d=21.94, SE=9.83, t=2.23, p=0.03). Qualitative findings: Almost 50% obtained some type of resource because of the support that was offered. Participants also frequently commented (31%) on the value of the peer connection they made with the peer specialist as a positive outcome. Less commonly, participants reported not following through on supports that were offered or even less frequently reported that the supports did not meet their needs. | The quantitative results from this study did not support our hypotheses but do offer some useful information. However, the qualitative data suggest that Centres for Independent Living do offer substantial supports in plausibly unique, unmet areas, with important results. |
| The Citizens/Citizenship project | Clayton et al. 2013 (USA)  N= 114 | Quantitative-RCT | Of those who were working within the 6-months prior to the baseline interview, part- or full-time (n=42, 37 %), citizenship intervention participants had significantly higher increases in satisfaction with work from baseline to both 6 and 12 months (B=.85, p=.01; B=5.19, p≤.001, respectively). A higher increase in satisfaction with ones’ finances from baseline to 6 months was found for those who received the intervention (B=.75, p=.01). No significant differences were found relating to social capital as measured by Hogan and Owen’s (2000) adaptation of the World Values Survey. Those in the intervention group also reported enhancements in amount of and satisfaction with social activity. | When controlling for baseline covariates, individuals in the citizenship intervention had reductions in alcohol and drug use as well as enhancements in amount of and satisfaction with social activity, satisfaction with finances, satisfaction with work, and overall quality of life. These findings suggest that the citizenship intervention may have facilitated, to some degree, participants’ efforts to build a life in the community. |
| The Citizens/Citizenship project | Pelletier et al., 2021 (Canada)  N= 15 | Quantitative- RCT | The only outcome measure which indicated a statistically significant difference between groups was the Citizenship Measure (i.e., p≤.05, 95% confidence interval) were found between the experimental group and the control group (p=.04). | Access to peer support groups facilitated by peer support workers is likely to allow members of these small groups to cope better with their personal daily challenges by maintaining meaningful relationships of mutual respect. In addition, and as its name suggests, the Citizen Project was initially designed as an intervention promoting the exercise of citizenship for people living with Severe mental illness. In this regard, this study may not have resulted in clear and statistically observable improvements, but in a maintenance that could be empirically observed with the use of a tool specifically designed for this purpose of assessing citizenship, namely the Citizenship Measure. |
| Peer Support Group Citizenship Intervetion | Rowe et al., 2007  (USA)  N=41 | Quantitative-RCT | The intervention showed no main effect for criminal justice involvement, although in both groups the number of new criminal charges reduced. | The research suggests that the citizenship intervention may be effective in reducing alcohol use among persons with severe mental illness and a criminal history. Findings did not support our hypothesis that the intervention group would have significantly less nonalcohol drug use and fewer criminal justice charges than the control group. |

RCT: Randomised controlled trial

### Appendix 5: References of Included sources

#### Housing

Burt, M. R. (2012). Impact of housing and work supports on outcomes for chronically homeless adults with mental illness: LA's HOPE. Psychiatric services, 63(3), 209-215.

Herman, D. B., Conover, S., Gorroochurn, P., Hinterland, K., Hoepner, L., & Susser, E. S. (2011). Randomized trial of critical time intervention to prevent homelessness after hospital discharge. Psychiatric services, 62(7), 713-719.

Nelson, G., Hall, G. B., & Walsh‐Bowers, R. (1997). A comparative evaluation of supportive apartments, group homes, and board‐and‐care homes for psychiatric consumer/survivors. Journal of community psychology, 25(2), 167-188.

Roos, E., Bjerkeset, O., Søndenaa, E., Antonsen, D. Ø., & Steinsbekk, A. (2016). A qualitative study of how people with severe mental illness experience living in sheltered housing with a private fully equipped apartment. BMC psychiatry, 16(1), 1-9.

Weissman, E. M., Covell, N. H., Kushner, M., Irwin, J., & Essock, S. M. (2005). Implementing peer-assisted case management to help homeless veterans with mental illness transition to independent housing. Community Mental Health Journal, 41(3), 267-276

##### Survey responses:

| Model | Additional information on this model |
| --- | --- |
| Rethink Mental Illness Accommodation  SURVEY RESPONSE 1 | **Staffordshire Supported Housing** Staffordshire Supported Housing provides high quality accommodation and low-level housing management support for people affected by mental illness aged 18 years and above, who are at risk of not accessing or sustaining a home because of their mental health, vulnerability and/or a history of housing related problems. The service has 57 accommodation units, located across four local authority areas and requires minimum stay of 6 months. There is also an out-of-hours housing related on-call service. The data broadly indicates various positive outcomes for service users. Since being in the accommodation, service users felt they were better able to manage their mental health, felt their living arrangement was more secure and found daily tasks easier to manage. Service users also reported an improved ability to manage their finances, felt more connected to local support networks and felt more confident about eventually living unsupported.  **Herbert House**  Herbert House is a registered care home (CQC) providing accommodation and personal care to 13 people with support needs related to their mental health. The service can support up to 15 people, with the other 2 beds allocated as “crisis beds” for short-term (1-2 week) stays. Service users have extremely positive perceptions of service delivery; they feel staff are knowledgeable, trustworthy and respectful and feel the right kind of support is provided by the service. The data also indicates various positive outcomes for service users: since being in the accommodation service users feel they are better able to manage their mental health, feel their living arrangement is more secure and feel more optimistic about their future. |
| Nottinghamshire supported accommodation  SURVEY RESPONSE 2 | Supported Accommodation is intended to achieve a phased step-down to the least supported housing option possible for people with severe and enduring mental health conditions and additional support needs. In Nottingham(shire) this includes specialist forensic mental health support, support to people with coexisting substance use, with learning disabilities and supporting autistic people with mental health illnesses. Usually, the first step down from inpatient psychiatric stays, people stay at the accommodation under licence and access intensive personalised support over 6-24 months from a 24/7 on-site team. The model is trauma and psychologically informed and is delivered by experienced highly-skilled mental health recovery workers with as many of these as possible being people with lived experience. They apply strengths-based assessment and support approaches to reinforce and build skills and confidence to live as independently as possible in the community. This includes becoming registered with primary healthcare providers and improving physical health, being registered to vote, accessing employment and education, (re)building relationships, optimising income, building budgeting skills, understanding, and sustaining a tenancy. The outcome is to support move-on to supported living or general housing options with personalised transition plans and informed ongoing support plans designed to further and sustain recovery and self-determination outcomes. |
| Stafford Intensive Supported Accommodation  SURVEY RESPONSE 7 | High Intensity Supported Accommodation is designed to support people with coexisting conditions who are rough sleeping or homeless. They are using substances and/or have poorly diagnosed or treated mental health illnesses. This is a cohort who have disengaged with previous services and have little faith or trust in “the system”. A specialist dual diagnosis team provides street support to individuals, encouraging them to access supported accommodation that is less restrictive. The team also engages with current and potential professional and personal supporter networks to build their confidence, skills and understanding in supporting each person, designing bespoke strategies that tackle many years of disengagement, disenfranchisement and of being seldom heard and seldom effectively supported in services. Accepting that recovery is not a linear journey, people may be supported in a housing setting or at street level for times when they relapse. The model is intended to become increasingly peer-led to improve credibility with people supported and to sustain their engagement. Further underpinning this aim is for evaluation and development of the model to be peer-led and actions coproduced and co-delivered with the support team. There is no time limit attached to support. It is available for as long as required. |

#### Money

Conrad, K. J., Lutz, G., Matters, M. D., Donner, L., Clark, E., & Lynch, P. (2006). Randomized trial of psychiatric care with representative payeeship for persons with serious mental illness. Psychiatric services, 57(2), 197-204.

Luchins, D. J., Hanrahan, P., Conrad, K. J., Savage, C., Matters, M. D., & Shinderman, M. (1998). An agency-based representative payee program and improved community tenure of persons with mental illness. Psychiatric services, 49(9), 1218-1222.

Mental Health and Money Advice. Mental Health & Money Toolkit. Retrieved from https://www.mentalhealthandmoneyadvice.org/media/1444/mhma-money-toolkit-a5-february-2022.pdf

Mental Health and Money Advice. Mental Health Crisis Breathing Space: Guide for Approved Mental Health Professionals. Retrieved from https://www.mentalhealthandmoneyadvice.org/media/1440/a-guide-for-amhps.pdf

Rosen, M. I., Desai, R., Bailey, M., Davidson, L., & Rosenheck, R. (2001). Consumer experience with payeeship provided by a community mental health center. Psychiatric Rehabilitation Journal, 25(2), 190.

##### Survey responses:

| Model | Additional information on this model |
| --- | --- |
| Hertfordshire & Sheffield Mental Health Project  [SURVEY RESPONSE 3] | Hertfordshire Mental Health Project - co-located services. The Mental Health Project takes referrals directly from community-based mental health clinical staff. Money advisors, who are located in mental health settings including CMHTs and Crisis Assessment and Treatment Teams, work with patients to increase their incomes and reduce mental health crises by improving mental health and wellbeing. The service also aims to reduce demand on mental health and specialist staff so they can concentrate on providing clinical care. The service, which is now approaching its ninth year, has received more than 7,500 referrals from community-based mental health clinical staff. Advisors have achieved around £32 million in extra benefits of debt-write off and clinical staff estimate that in a single year the money advice service helped prevent: 40 hospital admissions, 30 episodes of homelessness and 32 episodes of service harm.  Mental health transformation project -This programme provides support for people in primary and community mental health services. Patients are assigned an adviser with mental health, debt and financial capability expertise, who then works with a client’s whole support structure to sustain a more financially secure future. Most of the work relates to debt and welfare benefits and in 2021/22, the service assisted 68 clients. |
| A combined money advice and psychological therapy intervention in IAPT  [SURVEY RESPONSE 8] | Where money advice clients have the capabilities and self-efficacy to act on advice it can be incredibly effective: 80% of people who receive money advice feel more in control of their finances. Building on the need for integrated money advice and mental health services, below we present details of a pilot we’ve partnered on with King’s College London and Citizens Advice. A pilot currently underway in South London and Maudsley (SLaM) NHS Foundation Trust, trialling a combined IAPT and money advice intervention to increase recovery rates in IAPT. The pilot is being delivered to people with common mental disorders, such as anxiety and depression, who are receiving High-Intensity Therapy (HIT) in IAPT. Clients are routinely asked about money worries during referral to IAPT HIT services. Where money worries are identified, clients are directly referred to Citizens Advice for a follow-up appointment with a money advisor. The two services, money advice and IAPT - can then be delivered either concurrently, or separately - dependent upon the client's preference. Initial statistical modelling has shown that co-located money advice in IAPT supports improved recovery outcomes for service users. This model of intervention has also been found to be cost-effective. A simple cost-benefit analysis, drawing on economic modelling undertaken at LSE, suggested that such an intervention would generate a small surplus of healthcare savings (£2.4m) and a more substantial economic benefit (£105m) by reducing barriers to work and increasing productivity. The initial findings of the pilot are encouraging, and form a firm foundation for scaling-up a national pilot of the combined mental health and money advice intervention. To further investigate the effectiveness of co-located and integrated services, the DHSC should fund a national pilot to understand and evaluate the effectiveness of co-located debt advice in the IAPT setting. |

#### Work and education

Camden and Islington NHS Foundation Trust. Choice and Control Peer Coaching Service. Retrieved from <https://www.candi.nhs.uk/our-services/choice-and-control-peer-coaching-service>

Crisis Resolution Team Optimisation and Relapse Prevention (CORE UCL). "A peer-delivered self-management intervention to bridge the gap between crisis and continuing care." from <https://www.ucl.ac.uk/core-study/workstream-2>

Dewa, C. S., Hoch, J. S., Carmen, G., Guscott, R., & Anderson, C. (2009). Cost, effectiveness, and cost-effectiveness of a collaborative mental health care program for people receiving short-term disability benefits for psychiatric disorders. The Canadian Journal of Psychiatry, 54(6), 379-388.

Gutman, S. A., Kerner, R., Zombek, I., Dulek, J., & Ramsey, C. A. (2009). Supported education for adults with psychiatric disabilities: Effectiveness of an occupational therapy program. The American Journal of Occupational Therapy, 63(3), 245-254.

Indistrial Services Group. Social Firms England. Retrieved from https://industrialservicesgroup.co.uk/social-enterprise/social-firms-england/

Kordsmeyer, A.-C., Efimov, I., Lengen, J. C., Harth, V., & Mache, S. (2022). Workplace Health Promotion in German Social Firms—Offers, Needs and Challenges from the Perspectives of Employees, Supervisors and Experts. International journal of environmental research and public health, 19(2), 959.

Macias, C., Rodican, C. F., Hargreaves, W. A., Jones, D. R., Barreira, P. J., & Wang, Q. (2006). Supported employment outcomes of a randomized controlled trial of ACT and clubhouse models. Psychiatric services, 57(10), 1406-1415.

Major, B. S., Hinton, M. F., Flint, A., Chalmers-Brown, A., McLoughlin, K., & Johnson, S. (2010). Evidence of the effectiveness of a specialist vocational intervention following first episode psychosis: a naturalistic prospective cohort study. Social psychiatry and psychiatric epidemiology, 45(1), 1-8. doi:10.1007/s00127-009-0034-4

Morant, N., Milton, A., Gilbert, E., Johnson, S., Parsons, N., Singh, S., & Marwaha, S. (2021). Vocational rehabilitation via social firms: a qualitative investigation of the views and experiences of employees with mental health problems, social firm managers and clinicians. BMC psychiatry, 21(1), 1-11.

Mental Health and Productivity Pilot. (2021). Helping People in Work with Mental Health Problems to Perform their Best. Retrieved from https://mhpp.me/helping-people-in-work-with-mental-health-problems-to-perform-at-their-best/

Nuechterlein, K. H., Subotnik, K. L., Ventura, J., Turner, L. R., Gitlin, M. J., Gretchen-Doorly, D., . . . Liberman, R. P. (2020). Enhancing return to work or school after a first episode of schizophrenia: the UCLA RCT of Individual Placement and Support and Workplace Fundamentals Module training. Psychological medicine, 50(1), 20-28.

Perkins, R., Rinaldi, M., & Hardisty, J. (2010). Harnessing the expertise of experience: increasing access to employment within mental health services for people who have themselves experienced mental health problems. Diversity in Health & Care, 7(1).

Resources, D. I. (2011). Social Firms in Europe. Retrieved from https://www.dinf.ne.jp/doc/english/resource/employment/Social_Firms_in_Europe.html

Rinaldi, M., Perkins, R., McNeil, K., Hickman, N., & Singh, S. P. (2010). The Individual Placement and Support approach to vocational rehabilitation for young people with first episode psychosis in the UK. Journal of Mental Health, 19(6), 483-491.

Sauvé, G., Buck, G., Lepage, M., & Corbière, M. (2021). Minds@ Work: A New Manualized Intervention to Improve Job Tenure in Psychosis Based on Scoping Review and Logic Model. Journal of Occupational Rehabilitation, 1-14.

Schindler, V. P., & Sauerwald, C. (2013). Outcomes of a 4-year program with higher education and employment goals for individuals diagnosed with mental illness. Work, 46(3), 325-336.

Social Firms England. Retrieved from socialfirmsengland.co.uk

Svanberg, J., Gumley, A., & Wilson, A. (2010). How do social firms contribute to recovery from mental illness? A qualitative study. Clinical Psychology & Psychotherapy, 17(6), 482-496.

UK Government. Access to work Retrieved from https://www.gov.uk/access-to-work

Unger, K. V., Pardee, R., & Shafer, M. S. (2000). Outcomes of postsecondary supported education programs for people with psychiatric disabilities. Journal of Vocational Rehabilitation, 14(3), 195-199.

#### Social Isolation

Atkinson, J. M., Coia, D. A., Gilmour, W. H., & Harper, J. P. (1996). The impact of education groups for people with schizophrenia on social functioning and quality of life. The British journal of psychiatry, 168(2), 199-204.

Beavan, V., de Jager, A., & dos Santos, B. (2017). Do peer-support groups for voice-hearers work? A small scale study of Hearing Voices Network support groups in Australia. Psychosis, 9(1), 57-66.

Boevink, W., Kroon, H., van Vugt, M., Delespaul, P., & van Os, J. (2016). A user-developed, user run recovery programme for people with severe mental illness: A randomised control trial. Psychosis, 8(4), 287-300.

Botero-Rodríguez, F., Hernandez, M. C., Uribe-Restrepo, J. M., Cabariqe, C., Fung, C., Priebe, S., & Gómez-Restrepo, C. (2021). Experiences and outcomes of group volunteer befriending with patients with severe mental illness: an exploratory mixed-methods study in Colombia. BMC psychiatry, 21(1), 1-10.

Bromage, B., Kriegel, L., Williamson, B., Maclean, K., & Rowe, M. (2017). Project Connect: A community intervention for individuals with mental illness. American Journal of Psychiatric Rehabilitation, 20(3), 218-233.

Castelein, S., Bruggeman, R., Van Busschbach, J. T., Van Der Gaag, M., Stant, A., Knegtering, H., & Wiersma, D. (2008). The effectiveness of peer support groups in psychosis: a randomized controlled trial. Acta Psychiatrica Scandinavica, 118(1), 64-72.

Chinman, M., Young, A. S., Hassell, J., & Davidson, L. (2006). Toward the implementation of mental health consumer provider services. The Journal of Behavioral Health Services & Research, 33(2), 176-195.

Citizens community collaborative. FACE. Retrieved from https://citizens.collaborative.yale.edu/programs/community-action/face

Davidson, L., Haglund, K. E., Stayner, D. A., Rakfeldt, J., Chinman, M. J., & Kraemer Tebes, J. (2001). " It was just realizing… that life isn't one big horror": A qualitative study of supported socialization. Psychiatric Rehabilitation Journal, 24(3), 275.

Dayson, C., Painter, J., & Bennett, E. (2020). Social prescribing for patients of secondary mental health services: emotional, psychological and social well-being outcomes. Journal of Public Mental Health, 19(4), 271-279.

Ercan Doğu, S., Kayıhan, H., Kokurcan, A., & Örsel, S. (2021). The effectiveness of a combination of Occupational Therapy and Social Skills Training in people with schizophrenia: A rater-blinded randomized controlled trial. British Journal of Occupational Therapy, 84(11), 684-693.

Frerichs, J., Billings, J., Barber, N., Chhapia, A., Chipp, B., Shah, P., . . . Evans, B. L. (2020). Influences on participation in a programme addressing loneliness among people with depression and anxiety: findings from the Community Navigator Study. BMC psychiatry, 20(1), 1-15.

Giacco, D., Chevalier, A., Patterson, M., Hamborg, T., Mortimer, R., Feng, Y., . . . Priebe, S. (2021). Effectiveness and cost-effectiveness of a structured social coaching intervention for people with psychosis (SCENE): protocol for a randomised controlled trial. BMJ open, 11(12), e050627.

Haslam, C., Cruwys, T., Chang, M. X.-L., Bentley, S. V., Haslam, S. A., Dingle, G. A., & Jetten, J. (2019). GROUPS 4 HEALTH reduces loneliness and social anxiety in adults with psychological distress: Findings from a randomized controlled trial. Journal of consulting and clinical psychology, 87(9), 787.

Haslam, C., Cruwys, T., Haslam, S. A., Dingle, G., & Chang, M. X.-L. (2016). Groups 4 Health: Evidence that a social-identity intervention that builds and strengthens social group membership improves mental health. Journal of affective disorders, 194, 188-195.

Hasson-Ohayon, I., Mashiach-Eizenberg, M., Avidan, M., Roberts, D. L., & Roe, D. (2014). Social cognition and interaction training: preliminary results of an RCT in a community setting in Israel. Psychiatric services, 65(4), 555-558.

Lambeth Vocational Services. Expand your world: A serries of co-designed workshops. Retrieved from https://www.lambethcollaborative.org.uk/wp-content/uploads/2020/09/Expand-your-world-.pdf

Lloyd-Evans, B., Frerichs, J., Stefanidou, T., Bone, J., Pinfold, V., Lewis, G., . . . Chipp, B. (2020). The Community Navigator Study: Results from a feasibility randomised controlled trial of a programme to reduce loneliness for people with complex anxiety or depression. Plos one, 15(5), e0233535.

McCarthy, S., Chinman, M., Mitchell-Miland, C., Schutt, R. K., Zickmund, S., & Ellison, M. L. (2019). Peer specialists: Exploring the influence of program structure on their emerging role. Psychological services, 16(3), 445.

Mccorkle, B. H., Dunn, E. C., Wan, Y. M., & Gagne, C. (2009). Compeer friends: a qualitative study of a volunteer friendship programme for people with serious mental illness. International Journal of social psychiatry, 55(4), 291-305.

McCorkle, B. H., Rogers, E. S., Dunn, E. C., Lyass, A., & Wan, Y. M. (2008). Increasing social support for individuals with serious mental illness: Evaluating the compeer model of intentional friendship. Community Mental Health Journal, 44(5), 359-366.

Mizock, L., Russinova, Z., & DeCastro, S. (2015). Recovery narrative photovoice: Feasibility of a writing and photography intervention for serious mental illnesses. Psychiatric Rehabilitation Journal, 38(3), 279.

Petryshen, P. M., Hawkins, J. D., & Fronchak, T. A. (2001). An evaluation of the social recreation component of a community mental health program. Psychiatric Rehabilitation Journal, 24(3), 293.

Priebe, S., Chevalier, A., Hamborg, T., Golden, E., King, M., & Pistrang, N. (2020). Effectiveness of a volunteer befriending programme for patients with schizophrenia: randomised controlled trial. The British journal of psychiatry, 217(3), 477-483.

Rivera, J. J., Sullivan, A. M., & Valenti, S. S. (2007). Adding consumer-providers to intensive case management: does it improve outcome? Psychiatric services, 58(6), 802-809.

Saavedra, J., Arias, S., Crawford, P., & Pérez, E. (2018). Impact of creative workshops for people with severe mental health problems: art as a means of recovery. Arts & Health, 10(3), 241-256.

Schindler, V. P. (2005). Role development: an evidenced-based intervention for individuals diagnosed with schizophrenia in a forensic facility. Psychiatric Rehabilitation Journal, 28(4), 391.

Sheridan, A., O’Keeffe, D., Coughlan, B., Frazer, K., Drennan, J., & Kemple, M. (2018). Friendship and money: A qualitative study of service users’ experiences of participating in a supported socialisation programme. International Journal of social psychiatry, 64(4), 326-334.

Sheridan, A. J., Drennan, J., Coughlan, B., O’Keeffe, D., Frazer, K., Kemple, M., . . . Kow, V. (2015). Improving social functioning and reducing social isolation and loneliness among people with enduring mental illness: Report of a randomised controlled trial of supported socialisation. International Journal of social psychiatry, 61(3), 241-250.

Terzian, E., Tognoni, G., Bracco, R., De Ruggieri, E., Ficociello, R. A., Mezzina, R., & Pillo, G. (2013). Social network intervention in patients with schizophrenia and marked social withdrawal: a randomized controlled study. The Canadian Journal of Psychiatry, 58(11), 622-631.

Webber, M. (2014). From ethnography to randomized controlled trial: an innovative approach to developing complex social interventions. Journal of Evidence-Based Social Work, 11(1-2), 173-182.

Webber, M., Morris, D., Howarth, S., Fendt-Newlin, M., Treacy, S., & McCrone, P. (2019). Effect of the connecting people intervention on social capital: a pilot study. Research on Social Work Practice, 29(5), 483-494.

Webber, M., Ngamaba, K., Moran, N., Pinfold, V., Boehnke, J., Knapp, M., . . . Morris, D. (2021). The implementation of connecting people in community mental health teams in England: a quasi-experimental study. The British Journal of Social Work, 51(3), 1080-1100.

Webber, M., Reidy, H., Ansari, D., Stevens, M., & Morris, D. (2016). Developing and modeling complex social interventions: Introducing the connecting people intervention. Research on Social Work Practice, 26(1), 14-19.

Whitley, R., Sitter, K. C., Adamson, G., & Carmichael, V. (2021). A meaningful focus: Investigating the impact of involvement in a participatory video program on the recovery of participants with severe mental illness. Psychiatric Rehabilitation Journal, 44(1), 63.

Williams, E., Dingle, G. A., Calligeros, R., Sharman, L., & Jetten, J. (2020). Enhancing mental health recovery by joining arts-based groups: a role for the social cure approach. Arts & Health, 12(2), 169-181.

Yanos, P. T., Stefancic, A., Alexander, M. J., Gonzales, L., & Harney-Delehanty, B. (2018). Association between housing, personal capacity factors and community participation among persons with psychiatric disabilities. Psychiatry Research, 260, 300-306.

#### Family

APA achievement awards. (2003). 2003 APA Gold Award: Helping Parents With Serious Mental Illness Retain Custody of Their Children: FSS/PACE Program. Psychiatric services, 54(11), 1526-1528. doi:10.1176/appi.ps.54.11.1526

Brunette, M. F., Richardson, F., White, L., Bemis, G., & Eelkema, R. E. (2004). Integrated family treatment for parents with severe psychiatric disabilities. Psychiatric Rehabilitation Journal, 28(2), 177.

Hanrahan, P., McCoy, M. L., Cloninger, L., Dincin, J., Zeitz, M. A., Simpatico, T. A., & Luchins, D. J. (2005). The mothers' project for homeless mothers with mental illnesses and their children: a pilot study. Psychiatric Rehabilitation Journal, 28(3), 291.

Waldo, M. C., Roath, M., Levine, W., & Freedman, R. (1987). A model program to teach parenting skills to schizophrenic mothers. Psychiatric services, 38(10), 1110-1112.

#### Victimisation

Albers, W. M., Roeg, D. P., Nijssen, Y. A., Deen, M. L., Bongers, I., & van Weeghel, J. (2021). Intervention to prevent and manage the effects of victimization related to social participation for people with severe mental illness: Results from a cluster randomized controlled trial. Psychiatric Rehabilitation Journal.

De Waal, M. M., Dekker, J. J., Kikkert, M. J., Christ, C., Chmielewska, J., Staats, M. W., . . . Goudriaan, A. E. (2019). Self‐wise, Other‐wise, Streetwise (SOS) training, an intervention to prevent victimization in dual‐diagnosis patients: results from a randomized clinical trial. Addiction, 114(4), 730-740.

Melendez-Torres, G. J., Pell, B., Buckley, K., Couturiaux, D., Trickey, H., Young, H., . . . Robinson, A. Health Pathfinder: A Full Technical Report. Retrieved from https://safelives.org.uk/sites/default/files/resources/Health%20Pathfinder%20Full%20Technical%20Report%20-%20March%202021.pdf

Ruijne, R., Kamperman, A. M., Trevillion, K., Garofalo, C., Van der Gaag, M., Zarchev, M., . . . Mulder, C. L. (2020). Assessing the acceptability, feasibility and sustainability of an intervention to increase detection of domestic violence and abuse in patients suffering from severe mental illness: A qualitative study. Frontiers in psychiatry, 11, 581031.

Ruijne, R., Mulder, C., Zarchev, M., Trevillion, K., van Est, R., Leeman, E., . . . Bogaerts, S. (2022). Detection of domestic violence and abuse by community mental health teams using the brave intervention: a multicenter, cluster randomized controlled trial. Journal of interpersonal violence, 37(15-16), NP14310-NP14336.

Safe Lives. LINKS: Towards a better response to domestic violence & abuse in mental health. Retrieved from https://safelives.org.uk/sites/default/files/resources/BEH-MHT%20LINKS%20pilot%20evaluation.pdf

Trevillion, K., Byford, S., Cary, M., Rose, D., Oram, S., Feder, G., . . . Howard, L. (2014). Linking abuse and recovery through advocacy: an observational study. Epidemiology and psychiatric sciences, 23(1), 99-113.

#### Offending

Bagwell, M. (2013). Does Riverside Mental Health Court Reduce Re-arrest Among Mentally Ill Offenders? : Alliant International University.

Bellamy, C., Kimmel, J., Costa, M. N., Tsai, J., Nulton, L., Nulton, E., . . . O’Connell, M. (2019). Peer support on the “inside and outside”: building lives and reducing recidivism for people with mental illness returning from jail. Journal of Public Mental Health.

Bonfine, N., Ritter, C., Teller, J. L., & Munetz, M. R. (2018). A comparison of participants in two community-based programs: Assisted outpatient treatment and a mental health court. Psychiatric services, 69(9), 1001-1006.

Campbell, M. A., Canales, D. D., Wei, R., Totten, A. E., Macaulay, W. A. C., & Wershler, J. L. (2015). Multidimensional evaluation of a mental health court: Adherence to the risk-need-responsivity model. Law and Human Behavior, 39(5), 489.

Canada, K. E., Trawver, K. R., & Barrenger, S. (2020). Deciding to participate in mental health court: Exploring participant perspectives. International journal of law and psychiatry, 72, 101628.

Disley, E., Gkousis, E., Hulme, S., Morley, K. I., Pollard, J., Saunders, C. L., . . . Sutherland, A. (2021). Outcome Evaluation of the National Model for Liaison and Diversion. Santa Monica, CA: RAND Corporation.

Duwe, G. (2015). Does release planning for serious and persistent mental illness offenders reduce recidivism? Results from an outcome evaluation. Journal of Offender Rehabilitation, 54(1), 19-36.

Eckberg, D., Podkopacz, M. R., Zehm, K., & Kubits, G. (2006). Evaluation of the Hennepin County mental health court. In: Hennepin County, Minnesota. Fourth Judicial District Research Division.

Han, W. (2020). Effect of behavioral health services and neighborhood disadvantages on recidivism: a comparison of mental health court and traditional court participants. Journal of experimental criminology, 16(1), 119-140.

Han, W., & Redlich, A. D. (2016). The impact of community treatment on recidivism among mental health court participants. Psychiatric services, 67(4), 384-390.

Hiday, V. A., Ray, B., & Wales, H. (2016). Longer-term impacts of mental health courts: Recidivism two years after exit. Psychiatric services.

Hiday, V. A., Wales, H. W., & Ray, B. (2013). Effectiveness of a short-term mental health court: Criminal recidivism one year postexit. Law and Human Behavior, 37(6), 401.

Linhorst, D. M., Dirks-Linhorst, P., Stiffelman, S., Gianino, J., Bernsen, H. L., & Kelley, B. J. (2010). Implementing the essential elements of a mental health court: The experiences of a large multijurisdictional suburban county. The Journal of Behavioral Health Services & Research, 37(4), 427-442.

Linhorst, D. M., Kondrat, D., Eikenberry, J., & Dirks-Linhorst, P. (2020). The role of mental health courts in mitigating family violence. Journal of interpersonal violence.

Lowder, E. M., Desmarais, S. L., & Baucom, D. J. (2016). Recidivism following mental health court exit: Between and within-group comparisons. Law and Human Behavior, 40(2), 118.

Luskin, M. L. (2013). More of the same? Treatment in mental health courts. Law and Human Behavior, 37(4), 255.

MacInnes, D., Khan, A. A., Tallent, J., Hove, F., Dyson, H., Grandi, T., & Parrott, J. (2021). Supporting prisoners with mental health needs in the transition to RESETtle in the community: the RESET study. Social psychiatry and psychiatric epidemiology, 56(11), 2095-2105.

McNiel, D. E., & Binder, R. L. (2007). Effectiveness of a mental health court in reducing criminal recidivism and violence. American Journal of Psychiatry, 164(9), 1395-1403.

Molyneaux, E., Vera San Juan, N., Brown, P., Lloyd-Evans, B., & Oram, S. (2021). A pilot programme to facilitate the use of mental health treatment requirements: Professional stakeholders’ experiences. The British Journal of Social Work, 51(3), 1041-1059.

Neiswender, J. R. (2005). An outcome evaluation of the mental health court of King County, Washington: An affirmation of the specialty court system: Washington State University.

O’Keefe, K. (2006). The Brooklyn mental health court evaluation. Planning, implementation, courtroom dynamics, and participant outcomes. New York, NY: Center for Court Innovation.

Ray, B. (2014). Long-term recidivism of mental health court defendants. International journal of law and psychiatry, 37(5), 448-454.

Richardson, E., & McSherry, B. (2010). Diversion down under—Programs for offenders with mental illnesses in Australia. International journal of law and psychiatry, 33(4), 249-257.

Roman, D. E. (2011). Examining Offender Recidivism and Severity of Offenses in a Mental Health Court: Northcentral University.

Steadman, H. J., Redlich, A., Callahan, L., Robbins, P. C., & Vesselinov, R. (2011). Effect of mental health courts on arrests and jail days: A multisite study. Archives of general psychiatry, 68(2), 167-172.

Trupin, E., & Richards, H. (2003). Seattle's mental health courts: Early indicators of effectiveness. International journal of law and psychiatry, 26(1), 33-53.

Wilson, A. B., Farkas, K., Bonfine, N., & Duda-Banwar, J. (2018). Interventions that target criminogenic needs for justice-involved persons with serious mental illnesses: a targeted service delivery approach. International journal of offender therapy and comparative criminology, 62(7), 1838-1853.

#### Citizenship and Multi-domain

Alvarez‐Jimenez, M., Koval, P., Schmaal, L., Bendall, S., O'Sullivan, S., Cagliarini, D., . . . Penn, D. L. (2021). The Horyzons project: a randomized controlled trial of a novel online social therapy to maintain treatment effects from specialist first‐episode psychosis services. World Psychiatry, 20(2), 233-243.

Anthony, W. A., Ellison, M. L., Rogers, E. S., Mizock, L., & Lyass, A. (2014). Implementing and evaluating goal setting in a statewide psychiatric rehabilitation program. Rehabilitation Counseling Bulletin, 57(4), 228-237.

Atterbury, K., & Rowe, M. (2017). Citizenship, community mental health, and the common good. Behavioral sciences & the law, 35(4), 273-287.

Catty, J., & Burns, T. (2001). Mental health day centres: their clients and role. Psychiatric Bulletin, 25(2), 61-66.

Clayton, A., O’Connell, M. J., Bellamy, C., Benedict, P., & Rowe, M. (2013). The citizenship project part II: Impact of a citizenship intervention on clinical and community outcomes for persons with mental illness and criminal justice involvement. American Journal of Community Psychology, 51(1), 114-122.

Ebrahim, S., Glascott, A., Mayer, H., & Gair, E. (2018). Recovery Colleges; how effective are they? The Journal of Mental Health Training, Education and Practice.

Eurodiaconia. (2013). A mapping of work integration social enterprises: Efforts on the outskirts. Retrieved from https://eurodiaconia.org/wordpress/wp-content/uploads/2015/09/misc_23_13-mapping_of_wise_eurodiaconia_members.pdf

Europe Economics. Look Ahead: The financial case for integrated mental health services and supported housing pathways. Retrieved from https://www.lookahead.org.uk/app/uploads/2021/02/Look-Ahead-Report-The-financial-case-for-integrated-care-2021.pdf

Hall, T., Jordan, H. L., Reifels, L., Belmore, S., Hardy, D., Thompson, H., & Brophy, L. (2018). A process and intermediate outcomes evaluation of an Australian recovery college. Journal of Recovery in Mental Health, 1(3), 7-20.

Hultqvist, J., Markström, U., Tjörnstrand, C., & Eklund, M. (2018). Quality of life among people with psychiatric disabilities attending community‐based day centres or Clubhouses. Scandinavian Journal of Caring Sciences, 32(4), 1418-1427.

Luck, D. (2018). Retrieved from http://democracy.sheffield.gov.uk/documents/s29526/Mental%20Health%20Social%20Cafes.pdf

Lynch, R. (2022). Home Group's hospital discharge and mental health services. Retrieved from https://www.housing.org.uk/news-and-blogs/blogs/rosie-lynch/home-groups-hospital-discharge-and-mental-health-services/

Macias, C., Rodican, C. F., Hargreaves, W. A., Jones, D. R., Barreira, P. J., & Wang, Q. (2006). Supported employment outcomes of a randomized controlled trial of ACT and clubhouse models. Psychiatric services, 57(10), 1406-1415.

Morris, D., Thomas, P., Ridley, J., & Webber, M. (2020). Community-Enhanced social prescribing: integrating community in policy and practice. International Journal of Community Well-Being, 1-17.

Moxham, L., Taylor, E. K., Patterson, C., Perlman, D., Brighton, R., Heffernan, T., & Sumskis, S. (2017). Goal setting among people living with mental illness: a qualitative analysis of recovery camp. Issues in mental health nursing, 38(5), 420-424.

Nieminen, I., Ramon, S., Dawson, I., Flores, P., Leahy, E., Pedersen, M. L., & Kaunonen, M. (2012). Experiences of social inclusion and employment of mental health service users in a European Union project. International Journal of Mental Health, 41(4), 3-23.

Pelletier, J.-F., Houle, J., Goulet, M.-H., Giguère, C.-É., Morin, C.-A., Juster, R.-P., . . . Denis, F. (2021). The Online Citizens Project: Effects oftransitional peer support groups facilitated by peer support workers for persons living with severe mental illness in times of the Covid-19 pandemic. Cadernos Brasileiros de Saúde Mental/Brazilian Journal of Mental Health, 13(36), 97-116.

Pernice‐Duca, F. M. (2008). The structure and quality of social network support among mental health consumers of clubhouse programs. Journal of community psychology, 36(7), 929-946.

Phipps, C., Seager, M., Murphy, L., & Barker, C. (2017). Psychologically informed environments for homeless people: resident and staff experiences. Housing, Care and Support.

NHS England Mental Health Secure Care Programme. (2021). Case Studies from the Specialist Community Forensic Team SCFT Pilot. Retrieved from

Prince, J. D., Mora, O., Ansbrow, J., Benedict, A., DiCostanzo, J., & Schonebaum, A. D. (2018). Nine ways that clubhouses foster interpersonal connection for persons with severe mental illness: lessons for other types of programs. Social Work in Mental Health, 16(3), 321-336.

Raeburn, T., Schmied, V., Hungerford, C., & Cleary, M. (2016). The use of social environment in a psychosocial clubhouse to facilitate recovery-oriented practice. BJPsych open, 2(2), 173-178.

Ramon, S., Griffiths, C. A., Nieminen, I., Pedersen, M., & Dawson, I. (2011). Towards social inclusion through lifelong learning in mental health: analysis of change in the lives of the EMILIA project service users. International Journal of social psychiatry, 57(3), 211-223.

LSE Housing and Communities. (2020). Tackling Homelessness: Case Studies. Retrieved from https://sticerd.lse.ac.uk/dps/case/cr/casereport132.pdfRethink. Mental Health Navigation: An established method for reducing pressure on primary and secondary care by supporting people to address their unmet non-clinical needs. A good practice guide. Retrieved from https://www.rethink.org/media/5466/mental-health-navigation.pdfRouse, J., Mutschler, C., McShane, K., & Habal-Brosek, C. (2017). Qualitative participatory evaluation of a psychosocial rehabilitation program for individuals with severe mental illness. International Journal of Mental Health, 46(2), 139-156.

Rowe, M., & Baranoski, M. (2011). Citizenship, mental illness, and the criminal justice system. International journal of law and psychiatry, 34(4), 303-308.

Rowe, M., Bellamy, C., Baranoski, M., Wieland, M., O'Connell, M. J., Benedict, P., . . . Sells, D. (2007). A peer-support, group intervention to reduce substance use and criminality among persons with severe mental illness. Psychiatric services, 58(7), 955-961.

Rowe, M., & Benedict, P. Citizenship, Mental Health, and the Citizens Project. Retrieved from <https://slidetodoc.com/citizenship-mental-health-and-the-citizens-project-michael/>

Rowe, M., & Davidson, L. (2016). Recovering citizenship. Israel Journal of Psychiatry and Related Sciences, 53(1), 14-21.

Rowe, M., & Pelletier, J.-F. (2012). Citizenship: A response to the marginalization of people with mental illnesses. Journal of Forensic Psychology Practice, 12(4), 366-381.

Salzer, M. S., Rogers, J., Salandra, N., O'Callaghan, C., Fulton, F., Balletta, A. A., . . . Brusilovskiy, E. (2016). Effectiveness of peer-delivered Center for Independent Living supports for individuals with psychiatric disabilities: A randomized, controlled trial. Psychiatric Rehabilitation Journal, 39(3), 239.

Shared Lives Plus. What is shared lives care? Retrieved from https://sharedlivesplus.org.uk/

Thames Reach. STAR. Retrieved from https://thamesreach.org.uk/what-we-do/prevention/star/

UCL Centre for Access to Justice. (2018). The Health Justice Landscape in England and Wales: Social welfare legal services in health settings. Retrieved from https://www.ucl.ac.uk/access-to-justice/sites/access-to-justice/files/lef030_mapping_report_web.pdf

Whitley, R., Harris, M., Fallot, R. D., & Berley, R. W. (2008). The active ingredients of intentional recovery communities: Focus group evaluation. Journal of Mental Health, 17(2), 173-182.

Wilson, C., King, M., & Russell, J. (2019). A mixed‐methods evaluation of a Recovery College in South East Essex for people with mental health difficulties. Health & social care in the community, 27(5), 1353-1362.

##### Survey responses:

| Model | Additional information on this model |
| --- | --- |
| Rethink Mental Illness Community Services  [SURVEY RESPONSE 6] | Our range of community support services give people severely affected by mental illness access to personalised help that can rebuild their confidence, help them stay in or return to work, and take part in social activities they enjoy. No community service is the same as we look to provide bespoke community services for different communities and their needs which can look to address a variety of life domains. https://www.rethink.org/help-in-your-area/services/community-support/  Here are a few examples of our work: Brent Mental Health Service  The Brent Mental Health service provides support to adults with mental health needs aged 18+, within the Brent borough alongside statutory services. The service has 4 main aspects of delivery which are Befriending service utilising volunteers; Peer Navigators Support; Substance Misuse Support delivered via Change, Grow, Live (CGL) staff; Mental Health and Recovery Workshops.  Wiltshire Mental Health Inclusion Service  The service offers 3- 6 months worth of one-to-one support with an inclusion coach, which centres around goal setting and identifying hobbies and interests, and also barriers to these (e.g. living rurally with no transport). Inclusion coaches also deliver mental health drop-in hubs around the county, with support from volunteers, and we have a tailored platform online called Wiltshire Clic. Anyone can use the hubs or the website at any time; you don’t need to be accessing 1-1 support. The service also supports transition into adult mental health support for ages 16+, and there is a close link with CAMHS to support young people leaving their services. Additionally, there are Digital Tech volunteers to support people to learn about getting online and how to access support virtually, and travel training volunteers to support with getting on public transport. These are supported by two members of staff (A digital officer and a peer and volunteer co-ordinator). More information can be found here: https://www.rethink.org/help-in-your-area/services/community-support/wiltshire-mental-health-inclusion-service/  Bristol Community Support Service The Bristol Community Support service works to help people with mental health issues make a positive difference in their lives. Staff work with people to build meaningful connections within their community. Their support is centred on values of recovery and social inclusion and a belief that with the right support individuals can become more independent, confident and resilient. |

#### Appendix 6: Mapping of models and additional description of the evidence base

#### Housing and homelessness

|  |  |  |  | **Broad types of support** | | | | |
| --- | --- | --- | --- | --- | --- | --- | --- | --- |
| **Main aim** | **An example model** | **Brief description** | **Source(s) of support** | **Signposting and information** | **Support and skills development** | **Direct resource provision** | **Increased rights and entitlement** | **More informal support** |
| Gaining housing and sustaining community living | Housing First | Rent assistance to support housing choice + intensive mental health support and practical assistance | Mental health and care staff |  | X | X |  |  |
| Supporting successful transition to and sustaining community living | Critical time intervention | Time limited support from a case worker for homeless people, to support transition to a housing tenancy in the community | Mental health and care staff |  | X |  |  |  |
|  | Peer enhanced case management | Support from a peer worker in addition to mental health usual care to support transition to community living for homeless veterans through help negotiating new surroundings, locating needed services, acting as mentors and encouraging socialisation with other participants | Peer workers | X | X |  |  | X |
|  | Los Angeles' Homeless Opportunity Providing Employment (LA's HOPE) | Aims to provide support to homeless mentally ill participants by providing supportive services and housing assistance, followed by support to gain employment through a variety of avenues | Domain specialists |  | X | X |  |  |
| Continuing to live in the community | Residential care | 24-hour staffed, high intensity support. Residents do not have tenancies. | Mental health and care staff |  | X | X |  |  |
|  | Supported accommodation | Medium intensity support, staffed group homes. Residents have tenancies | Mental health and care staff |  | X | X |  |  |
|  | Floating outreach support | Low intensity, off-site support from housing workers to people living independently | Mental health and care staff |  | X |  |  |  |
|  | Shared Lives Plus | Offer of accommodation and support from a host family who they share family and community life with | Lay people and organisations |  |  | X |  | X |

The **Housing First** model was identified as an effective housing model in our published systematic review (Barnett et al., 2022). Housing First (Tsemberis, 1999) supports people with serious mental health conditions who are homeless to gain and sustain independent housing. People are offered direct resources in the form of rent assistance to gain a housing tenancy at market rates and allow a degree of choice about type and location of housing; then high-intensity community mental health support, typically following an Assertive Community treatment model and including practical support with housing related needs, to help with mental health recovery and sustain independent housing. People are offered independent housing straight away, without any requirements to move through a supported, group housing pathway, or other preconditions like treatment adherence or sobriety. Trials of Housing First programmes have found they result in significant improvements in achievement and retention of stable housing (remaining housed) at both short- and long-term follow-up for their homeless, mentally ill client group (Aubry et al., 2016; Stergiopoulos et al., 2015). Further research confirms that rent assistance may be an essential ingredient of the Housing First model (Pankratz, Nelson, & Morrison, 2017).

We identified three other models of support for homeless adults with serious mental illness, both of which offered time-limited programmes to support transition to independent housing. **Critical Time Intervention (CTI)** offers intensive mental health care, involving a key worker offering mental health and practical support for homeless adults with serious mental illness, to support successful transition from psychiatric hospital to living in the community. Time-limited CTI support is followed by referral to standard mental health services. Herman and colleagues (2011) conducted a randomised controlled trial (RCT) in the USA and report reductions in homelessness over an 18-month follow-up period, from a nine-month CTI intervention. In a small pilot study of **Peer-assisted case management** Weissman, Covell, Kushner, Irwin, and Essock (2005) describe adding a “***Peer Advisor***” to standard case management care. The peer, a person with lived experience of mental health difficulties and homelessness, offered practical support and advice for homeless veterans with serious mental health conditions living in residential care, one month before transition to independent living, and for 12 months afterwards. They explicitly sought to help the person form connections with others in their local community, thus increasing their informal social support. We did not identify any robust evaluation of this model. The **Los Angeles’ Homeless Opportunity Providing Employment (LA’s HOPE)** was a state-funded programme in the USA, which provides support with housing, including rental assistance from domain specialists. The model also assists with employment. An observational comparison study (Burt, 2012) reported that service users spent more days housed and were more likely to be employed and work more days in competitive employment.

Three types of supported accommodation models have been distinguished, which have been used to support adults with serious mental health conditions in general, not just those who were previously homeless: residential care, supported housing, floating outreach (Killaspy et al., 2016; Priebe, Saidi, Want, Mangalore, & Knapp, 2009) . Examples of each of these types of support are provided in Appendices 2 and 3. These types of housing support can but need not necessarily be used as a pathway of stepped care towards independent living. **Residential care** provides 24-hour high-intensity support, usually from a staff team including qualified mental health professionals, including provision of meals and medication. Residents do not have tenancies. **Supported housing** involves group homes or cluster housing with housing support staff onsite, either 24 hours or some of the time, providing medium intensity support. Residents hold tenancies. Residential care and supported housing both involve direct resource provision, i.e. provide specific housing. ***Floating outreach*** involves housing support staff based off-site providing home visits to people living independently, providing practical support to help people maintain stable housing. Supported accommodation may help improve housing retention and clinical outcomes (McPherson, Krotofil, & Killaspy, 2018). However, most of this evidence came from longitudinal studies rather than RCTs, so is less conclusive. A recent systematic review also found that less than half of people moved on from higher to lower intensity supported accommodation within expected timeframes (Dalton-Locke, Marston, McPherson, & Killaspy, 2021). Appendices 2 and 3 provides examples from current practice of residential care, supported housing and floating outreach programmes in the UK and internationally.

We have located the UK **Shared Lives** programme within the Citizenship and multi-domain models section. However, for about half of its participants, the scheme offers people with mental health problems (or intellectual disabilities) who are struggling to live independently a home with a host family in their family home. Shared Lives is a nationwide scheme coordinated by a voluntary sector organisation. Host families are accredited and paid, and can offer practical and emotional support to the person living with them. Families and individuals meet first to discuss the person’s needs and check if both parties wish to proceed. The length of support is open-ended. We did not identify any evaluation of this model of support.

#### Money and basic needs

|  |  |  |  | **Broad types of support** | | | | |
| --- | --- | --- | --- | --- | --- | --- | --- | --- |
| **Main aim** | **An example model** | **Brief description** | **Source(s) of support** | **Signposting and information** | **Support and skills development** | **Direct resource provision** | **Increased rights and entitlement** | **More informal support** |
| Short-term relief from debt enforcement | Mental Health Crisis Breathing Space | Legal right to a break from creditors enforcing debts or non-payment charges so they can focus on mental health recovery. Debt support and money advice is also available. | Structural help, Mental health and care staff | X |  |  | X |  |
| Reduced debts and money worries through better access to advice and support | Money Advisors within mental health teams | Money advice and support from specialist advisors based in mental health teams | Domain specialists | X | X |  |  |  |
|  | Mental Health and Money Toolkit | Guidance for service users and mental health staff in managing money problems via a self-help “toolkit” alongside guidance for mental health professionals to help people manage finances and money stresses | Mental health and care staff | X | X |  |  |  |
| Better money management by outsourcing control of finances | Payeeship schemes | A representative payee receives and manages disability payments on behalf of a service user | Mental health and care staff, Domain specialists |  |  | X |  |  |

***Mental Health Crisis Breathing Space*** (Mental Health and Money Advice) is an English scheme for people using mental health crisis services, supported by legislation passed in 2021. Mental health staff may refer and confirm eligibility for someone to register with the scheme, which confers legal protection from creditors enforcing debts or applying interest or non-payment charges for the duration of the person’s crisis treatment plus 30 days afterwards. The scheme is administered by a mental health voluntary sector organisation (Rethink). We did not identify any evaluations of this recently-established scheme.

Appendix 3 provides several examples of specialist **Money Advisors within mental health teams**, within primary and secondary mental health care settings. Mental health staff can refer service users to the advisors, who provide practical help with benefits claims, budgeting and debt management. A pilot study in south London is currently conducting a preliminary evaluation of a scheme to embed money advisors within primary care psychological treatment (IAPT) services.

Through our online expert consultation, we also identified a UK model to improve support with money and debt provided by mental health and care staff who are not money advice specialists. The **Mental Health and Money Toolkit** (Mental Health and Money Advice) is a government funded, online resource with guidance for people using mental health services and mental health staff to support the development of money management skills and access to specialist money advice and support.

We have not identified any completed evaluations of these schemes.

We identified three US evaluations of **Payeeship** schemes, whereby a mental health service user’s finances are directly controlled and managed by a legally-registered, appointed representative. This could be the person’s mental health case manager (Luchins et al., 1998), a para-professional money manager within the mental health team with separated responsibilities from the care team (Rosen, Desai, Bailey, Davidson, & Rosenheck, 2001) or a money manager in a separate organisation from the mental health care team (Conrad 2006). These schemes were primarily used for adults who were assessed as lacking capacity to make financial decisions for themselves, but in one RCT (Conrad et al., 2006), people could also voluntarily self-refer to the scheme. Evaluations indicate positive effects on improved financial and clinical outcomes. Payeeship programmes are similar in nature to UK appointeeships (Disability Support Project), for which we found no evaluations.

#### Work and education

|  |  |  |  | **Broad types of support** | | | | |
| --- | --- | --- | --- | --- | --- | --- | --- | --- |
| **Main aim** | **An example model** | **Brief description** | **Source(s) of support** | **Signposting and information** | **Support and skills development** | **Direct resource provision** | **Increased rights and entitlement** | **More informal support** |
| Gaining and retaining paid employment | Individual Placement and Support (IPS) | Time-unlimited support from an employment specialist to find open market employment, then ongoing support, including engaging with employers, to retain and manage the job | Domain specialists | X | X |  |  | X |
|  | User Employment Programme | IPS support plus organisational-level action to increase mental health representation in the workforce | Domain specialists, Structural support | X | X |  |  | X |
|  | Peer-Support Workers (CORE Programme) | Provision of job opportunities as peer workers explicitly for people with mental health lived experience. Experience of using mental health crisis services, including inpatient wards and home treatment, was explicitly stated as desirable experience. Peer support workers were employed through a competitive interviewing process and provided with training and supervision through their NHS trust to support individuals’ recovery following discharge from community crisis services. | Structural support |  | X | X |  |  |
|  | Social Firms | Social enterprises which specifically employ people with disabilities or disadvantage, sometimes with government funding and support from employers | Structural support, Lay people and organisations |  | X | X |  | X |
|  | Access to Work programme | Increased entitlement to government funding to support employment for people with a physical or mental health condition or disability | Structural support |  |  | X | X |  |
| Job retention and reduced sickness absence | MENTOR Programme | Early intervention from a specialist liaison worker to help people experiencing mental health conditions to stay in the workplace and reduce the risk of having to go off on longer term sick leave through support to improve workplace functioning, wellbeing and productivity | Domain specialists |  | X |  |  | X |
|  | Mind@Work Programme | Modular programme addressing psychological barriers to employment | Mental Health staff |  | X |  |  |  |
| Gaining and completing educational qualifications | Bridge Programme | 12-week group programme developing study skills to assist people to pursue post-secondary education, employment or both. Sessions explore qualification options, skills for study and stress management alongside mentoring | Mental Health staff |  | X |  |  |  |
|  | VIBE Programme | Education-focused IPS support | Domain specialists | X | X |  |  | X |

**Individual Placement and Support (IPS)** has a very well-established evidence base of international studies as an effective intervention to help people with serious mental health problems get competitive, open-market paid employment (Barnett et al., 2022; Suijkerbuijk et al., 2017). IPS involves support from an ***employment specialist*** based within a mental health team to find paid work opportunities. The IPS worker offers help with developing skills in job applications and interviews, and supports people in managing challenges with work and sustaining employment. They work with employers to identify job opportunities and to support people to retain work, thus building up the informal support offered to people by their employer. IPS offers time-unlimited support, involves helping people find work of their choice, and seeks to help people find paid work right away. It is demonstrably more effective than alternative “train then place” approaches. A variety of psychological therapy augmentations to IPS have been developed and evaluated, with a lack of clear evidence for added value beyond the original IPS model (Barnett et al., 2022).

The **User Employment Programme** (Rinaldi et al., 2004) is a model of support within one NHS mental health Trust in England. It has offered IPS support to service users, and additional liaison with local employment agencies (job centres) to enhance the support provided to people with mental health problems, and proactive attempts to encourage the employment of service users within the mental health Trust – in all jobs including non-mental health care roles. This is achieved by developing a service charter and inclusive advertising. The initiative, evaluated with a diverse population, saw 15% of all jobs within the organisation awarded to people with mental health conditions over a five-year period, and substantial gains in overall rates of employment among the Trust’s service users.

***Peer support worker*** roles offer a specific employment opportunity where lived experience of mental health difficulties (including severe mental illness) is a requirement of the role, and peer workers use their expertise through experience to support others, offer specific employment opportunities for people with serious mental health conditions. **The peer-delivered self-management intervention model** as part of the **UCL CORE study** (Crisis Resolution Team Optimisation and Relapse Prevention (CORE UCL)) is one example model among many internationally of a mental health care provider organisation creating new peer support roles within mental health services: Peer workers were employed through a competitive interviewing process and provided with supervision and training.

**Social Firms** are a model of not-for-profit social enterprise which are mainly funded through their own activities, but often also receive government subsidies. They actively seek to employ and support people who are disadvantaged in the labour market, including people with serious mental illness. A UK survey in 2013 identified 76 social forms employing over 600 people with a mental health condition (Gilbert et al., 2013). The model is common in many European countries, and may provide employment opportunities for people who struggle to gain or retain competitive employment, however in the UK this model is more prevalent for groups of people with physical, sensory or intellectual disabilities. We did not identify any evaluations of their effectiveness.

**Access to Work** is an English government scheme which offers financial assistance to fund support for people with disabilities to find or retain work, including self-employment. For people with serious mental health conditions, this could include funding taxis or a support person to help them get to work, and individual support or a tailored plan to help them get or keep a job. The scheme is administered by ***voluntary sector organisations*** on behalf of the government and people can self-refer. The scheme thus directly provides resources to support employment, although not a job itself.

While the above models all seek to help people find and retain work, in-work support models for people already in employment focus exclusively on job retention and reducing sickness absence. The **MENTOR Programme** (The Mental Health and Productivity Pilot) involves support for people with mental health conditions including serious mental health conditions. Support is provided by an ***employment liaison worker*** employed by a mental health voluntary sector organisation. It involves support to the employee to manage challenges with work, and help and advice to employers to better support staff with mental health conditions. A pilot trial for the programme is due to report later in 2022. The **Minds@Work Programme** (Sauvé, Buck, Lepage, & Corbière, 2021) focuses exclusively on supporting the employee. Designed for people with psychosis who are in a job, it offers a nine-module programme of psychological support from a ***“psychosocial worker or counsellor”,*** delivered in work hours. We have not found any evaluations of this model.

Gutman, Kerner, Zombek, Dulek, and Ramsey (2009) conducted a small RCT of **The Bridge Programme**, a 12-week classroom-based group intervention designed to help adults with serious mental health conditions access educational programmes and develop study skills. Compared to usual care, the Bridge Programme model was effective, with over two thirds of participants enrolling in education or gaining employment within six months. **The VIBE Programme** (Major et al., 2010) is a UK example of IPS employment support being adapted to include support with accessing educational programmes leading to recognised academic or vocational qualifications.

#### Social isolation

|  |  |  |  | **Broad types of support** | | | | |
| --- | --- | --- | --- | --- | --- | --- | --- | --- |
| **Main aim** | **An example model** | **Brief description** | **Source(s) of support** | **Signposting and information** | **Support and skills development** | **Direct resource provision** | **Increased rights and entitlement** | **More informal support** |
| Improving problem solving, assertiveness and relationship skills to increase social contact | Education Groups | **Psychoeducation and social skills:** 20-week, psychoeducation and problem-solving group covering topics such as relating to other people, problem solving and assertiveness, and more general illness related topics such as causes of schizophrenia, managing symptoms and signs of relapse, with the hypothesis that being provided with information about their condition improves social functioning | Mental health and care staff | X | X |  |  |  |
| Improving social skills and identifying desired social activities to reduce social isolation | Social Skills Training and Occupational Therapy | **Psychoeducation and social skills:** 12-week group programme: sessions aimed at improving occupational and social skills with personalised goals. Themes include preparation for community living, practicing basic conversation skills, identifying desired activity goals and finding strategies to achieve them. Problem solving strategies to deal with daily living challenges are also taught. | Mental health and care staff |  | X |  |  |  |
| Support to identify and access social activities and increase social contact | Social Network Intervention | **Supported socialisation:** Suggestions for areas of interest for social activity participation for patients which is outside of the community mental health centre resources, followed by 3-6 months of support to help integrate the patient in the activity by either a member or staff or natural facilitators such as volunteers or family members | Mental health and care staff | X |  |  |  |  |
| Increased social contact through arts-based activity | Creative Workshops | **Supported socialisation:** 6, three-hour arts based creative workshops based in local museums | Mental health and care staff, Domain specialists |  | X |  |  |  |
| Signposting to community groups and activities and peer befriending to reduce social isolation | Rotherham Social Prescribing Mental Health Service | **Supported socialisation:** Support over 6 months from a social prescribing advisor and access to peer befriending and social groups | Domain specialists, Peer workers | X | X |  |  |  |
| Support to enhance community connections and reduce loneliness (Loneliness) | Community Navigators | **Supported socialisation:** 6-months, 10-sessions and group support from a social connections coach to help develop social relationships | Domain specialists | X | X | X |  |  |
| increased social activity, achieved by a structured self-management programme and peer support | Peer Specialists | **Supported socialisation:** 9-month, 40-session programme of structured self-management help and unstructured 1:1 peer support. | Peer workers | X | X |  |  |  |
| Increased social activity through regular meetings with a volunteer befriender | Volunteer Befriending Service | **Supported socialisation:** 1:1 support to do enjoyable activities with a matched, volunteer befriender who does not have a mental illness – over 1 year | Lay people and organisations |  |  |  |  | X |
| Changing cognitions and developing positive social identities through a structured group programme | Groups4Health (G4H) | **Changing cognitions:** Classroom-based, clinician-led, group programme helping to recognise and develop positive social identities and maintain social group relationships. | Mental health and care staff | X | X |  |  |  |
| Changing cognitions about social relationships to reduce social isolation | Social Cognition and Interaction Training (SCIT) | **Changing cognitions:** Weekly, clinician led, group programme helping with understanding emotions, recognising cognitive biases and applying these new skills to social behaviour | Mental health and care staff | X | X |  |  |  |

Of all the domains, we found most models of support in the social isolation domain, reflecting a diversity of approaches. Mann et al. (2017) distinguish three broad approaches to helping individuals with loneliness. These have also been used to typify models of support for subjective and objective social isolation (Ma et al., 2020). **Social skills training and psychoeducation** describes skills development models of support. **Supported socialisation** describes a support provider offering guidance and help with finding and accessing opportunities for social interaction. **Changing cognitions** describes psychological approaches to helping someone overcome intra-personal barriers to achieving fulfilling social relationships. The table above provides examples of all three types of support, delivered from a range of sources.

**1. Social skills training and psychoeducation** These models have provided mixed results from evaluations overall (Ma et al., 2020), but some successful trial evaluations have been undertaken. Atkinson, Coia, Gilmour, and Harper (1996) conducted a trial of 20-week, clinician-facilitated **Education Groups** for people with psychosis using community mental health services in Glasgow, covering illness management, but also relationships, housing and social issues. The programme increased the amount of social contact (social network size) for participants, compared to a usual care group. Ercan Doğu, Kayıhan, Kokurcan, and Örsel (2021) conducted a small trial of a group programme for people with psychosis using community mental health services in Turkey, comparing a 12-week group programme combining **Social Skills Training and Occupational Therapy *(OT)*** activity planning support to social skills training alone. Community integration (in family, social and occupational domains) improved in both groups over time, but significantly more in the social skills training plus OT group.

**2. Supported socialisation** These models, the most common among those we identified, are provided by mental health staff, domain specialists, peer workers or non-professional supporters, and offer a range of broad types of support. High quality evidence regarding the effectiveness of these models has yet to be established in most cases.

***Mental health staff-provided programmes*:** Terzian et al. (2013) describe a **Social Network Intervention** for people with psychosis using community mental health services in Canada. In addition to usual care, mental health staff offered regular support to individual service users to undertake desired social activities, also seeking to mobilise support from family and friends to achieve these goals. A trial evaluation found a positive effect for the programme in increasing participants’ social network size and quality, compared to a comparison group receiving usual community mental health care. **Connecting People** *(Webber, 2014; Webber et al., 2019; Webber et al., 2021; Webber, Reidy, Ansari, Stevens, & Morris, 2016)* is a model of support developed and tested in the UK, also delivered by ***mental health staff*** (following training) to better support people using mental health services to develop social connections and increase access to social capital. Evaluations suggest it may help achieve these goals when delivered with high fidelity, as intended (Webber, 2014), but it has proved hard to implement within NHS mental health services (Webber et al., 2021).

Saavedra, Arias, Crawford, and Pérez (2018) describe **Creative Workshops*,*** one of several arts-based models for reducing isolation we identified. Participants with a serious mental health condition were invited to attend six, three-hour workshops with their mental health worker, to create visual art and discuss it together. Sessions were held in local community arts venues and co-facilitated by a psychologist and arts specialist. We did not identify a robust evaluation of effectiveness for any of the arts-based models we found.

***Domain specialist and peer-provided programmes*:** The **Rotherham Social Prescribing Mental Health Service** (Dayson, Painter, & Bennett, 2020) adapts the social prescribing approach (University of Westminster) usually provided in primary care to offer more sustained and higher-intensity support for people with serious mental health conditions, supporting discharge from specialist community mental health care. People were offered linking support from a ***social prescribing advisor*** over a six-month period, plus access to a range of ***peer-provided*** befriending; providing support services. The **Community Navigator** programme (Frerichs et al., 2020; Lloyd-Evans et al., 2020) also uses a specialist link worker based within community mental health teams, but with a more exclusive focus on helping the person develop meaningful relationships and address loneliness, through individual and group support, and seeking to mobilise support from the person’s existing friends and family. The Community Navigator programme also directly provides financial help to support social participation, with access to a budget of up to £100 per participant (Lloyd-Evans et al., 2020). It is currently being evaluated in a UK-based RCT.

McCarthy et al. (2019) describe the addition of **Peer Specialists** to a community service for homeless veterans with mental health and substance misuse difficulties. Over nine months, as an addition to usual care, ***peer support workers*** offered 40 sessions: half offering structured self-management support, and half more unstructured support to meet individuals needs, with a common focus on reducing isolation, increasing community integration, and encouraging activities. This model is one example of a range of peer-provided models of support focusing on social isolation, including group and 1:1 support programmes (see Appendix 2).

While preliminary evaluations of several topic-expert and peer-provided models suggest promise, we did not identify any with definitive evidence of effectiveness in reducing subjective or objective social isolation.

***Lay people-provided programmes*:** Priebe and colleagues conducted a trial of a **Volunteer Befriending Service** for people with psychosis in community mental health care (Priebe et al., 2020). Participants were matched with a volunteer befriender (based on their interests and availability) and offered meetings at least monthly over a year, to undertake chosen, enjoyable activities together. The programme did not achieve all of its intended outcomes, but did lead to a significant increase in participants’ amount of social contact. A follow-up Australian exploratory study in a group format (rather than the original one-to-one format) reported improvements in objective social isolation following the intervention alongside qualitative reports of the development of trusting bonds between participants and befrienders (Botero-Rodríguez et al., 2021). While many the models we identified in this domain sought to mobilise available support from family and friends and help people build community connections, volunteering models directly provided additional support from members of the public, so have been marked as directly increasing informal support.

***3.* Changing cognitions models*:*** Compared to supported socialisation models, we found fewer psychological models of support for social isolation which have been developed or evaluated for people with serious mental health conditions. **Groups4Health (G4H)** is a five-session, clinician-facilitated, classroom-based programme focusing on the development and maintenance of valued social group memberships. It has trial evidence of effectiveness in reducing loneliness for adults experiencing psychological distress (Haslam et al., 2019; Haslam, Cruwys, Haslam, Dingle, & Chang, 2016), and is currently being piloted with a serious mental health condition population-adults with psychosis. **Social Cognition and Interaction Training (SCIT)** was designed for people with psychosis to help with understanding emotions and cognitive biases relating to social interactions, and adapt social behaviour accordingly. A trial in Israel (Hasson-Ohayon, Mashiach-Eizenberg, Avidan, Roberts, & Roe, 2014) found that practical support from a ***social mentor*** and SCIT psychological support was more effective than social mentor support alone in increasing social engagement for adults with psychosis.

A recent systematic review of trials of interventions for objective and subjective social isolation (Ma et al., 2020) concluded that current evidence is not sufficient to make recommendations for policy and practice about the most effective approaches. Our scoping review of the broader literature corroborates this, but demonstrates that a range of approaches, using different providers and types of support have been developed and used with people with serious mental health conditions in research and practice contexts.

#### Family relationships

|  |  |  |  | **Broad types of support** | | | | |
| --- | --- | --- | --- | --- | --- | --- | --- | --- |
| **Main aim** | **An example model** | **Brief description** | **Source(s) of support** | **Signposting and information** | **Support and skills development** | **Direct resource provision** | **Increased rights and entitlement** | **More informal support** |
| Support and provision of skills and education to help parents maintain custody of children | Parents, Advocacy, Coordination, Education (PACE) | Supportive and unconditional relationships developed with families and services such as case management parenting skills training, child development education and individual therapy provided. 24-hour on-call support and emergency assistance, monthly social support group for mothers, tenant-landlord mediation, crisis planning, financial assistance and transitional planning after hospitalisation are also provided, with most services provided within the home | Mental health and care staff, Domain specialists | X | X | X |  |  |
|  | Integrated Family Treatment | A family specialist clinician provides home-based services to parents and children while they participate in mental health services, e.g. engagement into treatment, linkage to environmental supports, education about child development, parenting skills training, modelling and coaching | Domain specialists | X | X |  |  |  |
|  | Mothers and Children’s Project | Program designed to supplement other mental health care which involves a visit by the program director, followed by weekly group meetings that focus on developmental education and role modelling | Mental health and care staff, Domain specialists, Lay people and organisations | X | X |  |  |  |
| Provide parents with support to meet basic needs in order to maintain or retain custody of their children | Thresholds mothers project | A problem-solving approach to psychosocial rehabilitation and intensive case management. Practical problems in daily living are a focus of the model, and mothers are helped to meet their basic needs, stabilize their living arrangements, and begin addressing psychiatric symptoms. Case managers help to secure entitlements, find independent apartments, and function as representative payees when needed. Care managers also assist with enrolling children in regular or special education | Mental health and care staff, domain specialists | X |  | X |  |  |

Three models described an approach to providing mothers with the necessary skills and education in child development. The **Mother’s and Children’s Project,** described in 1987, was the first to use this approach, which was designed to supplement other mental health care. A very small observational evaluation of ten families (Waldo, Roath, Levine, & Freedman, 1987) reported promising initial results for children in custody returning to their mothers.

**Integrated family treatment** also includes engagement into treatment, and linkage to environmental supports alongside education about child development and parental skills trainings. A small observational study (Brunette, Richardson, White, Bemis, & Eelkema, 2004) showed inconclusive results about impacts on custody of children.

**Parents, Advocacy, Coordination, Education (PACE)** provides a complete system of support, including the development of supportive and unconditional relationships between families and services, individual therapy, 24-hour on-call support, landlord-tenant mediation and financial planning. This model was described in 2003 (APA achievement awards, 2003) but no evaluation has been published.

The **Thresholds Mothers Project** differs in its focus on provision of support to meet basic needs in contrast to teaching parental skills. The model consists of intensive case management with practical problems being the main focus: stabilisation of living arrangements, representative payeeship and securing entitlements are all part of the approach. A retrospective chart review (Hanrahan et al., 2005) reported inconclusive but potentially promising findings regarding children remaining in their mother’s care following the programme.

#### Victimisation and exploitation

|  |  |  |  | **Broad types of support** | | | | |
| --- | --- | --- | --- | --- | --- | --- | --- | --- |
| **Main aim** | **An example model** | **Brief description** | **Source(s) of support** | **Signposting and information** | **Support and skills development** | **Direct resource provision** | **Increased rights and entitlement** | **More informal support** |
| Increase identification of patients suffering from domestic abuse and improve referral to specialist services | LARA (Linking Abuse and Recovery through Advocacy) | Domestic violence training for clinicians in community mental health teams, training for domestic violence advocacy professionals as well as referral to domestic violence advocacy for service users. | Mental health and care staff, domain specialists | X | X (for staff) |  |  |  |
| Directly contribute to the prevention of total, violent or property victimisation in people with mental health conditions | Self-wise, Other-wise, Streetwise (SOS) training | A six-week group training programme focused on enhancing emotion regulation skills, conflict resolution skills and street skills. | Mental health and care staff |  | X |  |  |  |
| Facilitate social participation as recovery for victimisation | The Victoria Intervention | Explores the victimisation experience with participants and then develops action plans to promote social participation | Mental health and care staff, Domain specialists, Peer workers | X | X |  |  |  |

Interventions aiming to improve identification and referral of people experiencing domestic abuse alongside mental health conditions involved a combination of ***mental health and care*** staff, and ***domain specialists.*** These included **Linking Abuse and Recovery through Advocacy (LARA)** (Trevillion et al., 2014)**,** a UK-based pilot intervention which trains clinicians to identify and respond to domestic violence, and also provides mental illness training to domestic violence advisors, and The **Better Reduction and Assessment of Violence (BRAVE)** (Ruijne et al., 2022) model, based on LARA. The LARA pilot reported promising improvements in clinician knowledge and attitudes, although the BRAVE cluster RCT (Ruijne et al., 2022) suggested that there was no accompanying increase in detection and referral rates, potentially due to clinician reported problems with retaining knowledge acquired in training (Ruijne et al., 2020). Another similar project (the **LINKS Project)** piloted a similar model, although unlike LARA and BRAVE, this pilot did not integrate a referral pathway. An observational mixed methods pre-post evaluation (Safe Lives) reported a 660% increase in referrals and an increase in staff understanding of how to record safeguarding concerns, enquire about domestic abuse and make referrals. However, there were reports of slow staff uptake of training.

The **Health Pathfinder** similarly is a multilevel system change intervention to change health response to domestic violence and abuse, including, as with similar models, training professionals, co-locating domestic violence and abuse advisors in clinical settings, and supporting identification and referral. In a pre-post evaluation (Melendez-Torres et al.) across eight different UK sites, Health Pathfinder reported an increased rate of domestic violence and abuse detection across a wide spectrum of risk.

**Self-wise, Other-wise, Streetwise (SOS) training** is a model of support in the Netherlands which focuses more broadly on victimisation of people with mental health conditions and aims instead to provide patients with the social skills and education to reduce conflict. The training is provided over six weeks by ***mental health and care staff*** and consists of 12 sessions focusing on emotion regulation skills training, conflict resolution skills training and behavioural training to help patients understand behavioural factors that contribute to their risk of victimisation. The model was evaluated in an RCT (De Waal et al., 2019) and reported that the training significantly reduced the odds of total victimisation, although when examining only violent victimisation the difference between participants receiving the intervention compared to usual care was not significant.

We identified one model which aimed to improve victimisation recovery for people with mental health problems through improving safe social participation: **The Victoria Intervention** involves ***mental health and care staff*** as well as ***domain specialists***, and ***peer workers***, who work together to explore how victimisation may impact social participation and develop an action plan to support service users to safely participate in their community. However, an RCT of the intervention (Albers et al., 2021) found no benefit in reducing victimisation although participants reported feeling more acknowledged and supported in their recovery process.

#### Offending

|  |  |  |  | **Broad types of support** | | | | |
| --- | --- | --- | --- | --- | --- | --- | --- | --- |
| **Main aim** | **Example models** | **Brief description** | **Source(s) of support** | **Signposting and information** | **Support and skills development** | **Direct resource provision** | **Increased rights and entitlement** | **More informal support** |
| Court diversion | Liaison and Diversion Schemes | Used to divert people with certain needs or vulnerabilities away from the criminal justice system and refer them to appropriate interventions or treatment in the healthcare system. | Structural support | X |  |  | X |  |
|  | Mental Health Courts | Link offenders with mental illness who would normally go to prison to long-term community-based mental health treatment. They have ongoing judicial monitoring to ensure offenders adhere to community treatment plans | Structural support | X |  |  | X |  |
| Facilitate community re-entry | Forensic Peer Support Services | A peer supporter with experience of mental health service use assists forensic service users (can be in a variety of roles) to help model recovery, connect with their community and address any re-entry difficulties e.g. accessing services when out of prison. | Peer workers | X |  |  |  |  |
|  | Severe and Persistent Mental illness (SPMI) release planning service | A pre-release transition service that attempts to connect offenders with needed services in the community following release e.g. housing, vocational, chemical dependency, psychiatric, disability, medical, medication, and transport needs | Domain specialists (release planners) | X |  |  |  |  |
|  | Supporting Prisoners upon Prison Release Service (RESET) | Supports prisoners with mental health needs for 12 weeks after release to coordinate their transition into the community and obtaining secure housing | Domain specialists | X |  | X |  |  |
| Target changeable factors associated with criminal activity | Thinking for a change (T4C) | A structured programme of 25 sessions delivered over 3 months covering social skills training, cognitive restructuring and problem-solving | Domain specialists |  | X |  |  |  |

Of models aimed at diverting people with mental health conditions away from the criminal justice system, we found the most evidence for **Mental Health Courts**, originating predominantly from the USA (Bagwell, 2013; Bonfine, Ritter, Teller, & Munetz, 2018; Canada, Trawver, & Barrenger, 2020; Eckberg, Podkopacz, Zehm, & Kubits, 2006; Han, 2020; Han & Redlich, 2016; Hiday, Ray, & Wales, 2016; Hiday, Wales, & Ray, 2013; Linhorst et al., 2010; Linhorst, Kondrat, Eikenberry, & Dirks-Linhorst, 2020; Lowder, Desmarais, & Baucom, 2016; Luskin, 2013; McNiel & Binder, 2007; Neiswender, 2005; O’Keefe, 2006; Ray, 2014; Roman, 2011; Steadman, Redlich, Callahan, Robbins, & Vesselinov, 2011; Trupin & Richards, 2003), but also with evidence from Australia (Richardson & McSherry, 2010) and Canada (Campbell et al., 2015). These provide ***structural support,*** linking offenders with mental illness to long-term community-based mental health treatment as an alternative to prison. Ongoing judicial monitoring ensures offenders adhere to community treatment plans, although the decision to participate in the court instead of prison is voluntary. Among evaluation evidence found, nine quasi-experimental studies reported outcomes related to changes in recidivism: the majority (seven of nine) reported that participants who completed the prescribed treatment had significantly reduced rearrests and time in jail, although one trial reported that in those that were arrested, the offenses were no less severe. Participants who dropped out of the Mental Health Court and returned to prison settings had similar rearrests to participants who did not participate at all. Similarly promising results were reported in observational studies (Appendix 4).

**Liaison and Diversion Schemes** are a similar model of support described in both the UK (Disley et al., 2021) and Australia (Richardson & McSherry, 2010), however they differ in that, unlike Mental Health Courts, there is no ongoing offender supervision role through court. Disley et al. (2021) evaluated the impact of the Liaison and diversion schemes in 27 sites in England. They concluded that the likelihood of service users receiving a custodial sentence is halved following involvement with these services. They may reduce the proportion of offences resulting in custodial sentences and thus increase diversion from the criminal justice system.

Three models focused on facilitating re-entry to the community following a prison sentence for people with mental health problems. **Forensic Peer Support Services** involve a ***peer supporter*** with experience of mental health service use who assists people, in a variety of roles, to connect with their community, including facilitating resolution of any difficulties accessing mental health and community services when out of prison (Bellamy et al., 2019). No evaluation of these services has been carried out. **Severe and Persistent Mental Illness (SPMI) release planning services** are provided by ***domain specialists*** who connect service users *prior* to release from prison with required services in the community to facilitate their reintegration once they are released. A quasi-experimental trial (Duwe, 2015) reported that the release planning group had slightly lower (non-significantly different) rates of rearrests and reincarceration compared to comparators, although reconviction rates were slightly higher (54% vs 53% in comparators), suggesting that pre-release planning may not contribute to reduced re-offending. **The Supporting Prisoners upon Release Service (RESET)** similarly signposts service users to useful community supports but also helps them to obtain housing following release from prison. In an observational two-group study (MacInnes et al., 2021), RESET participants were more likely to have contact with primary health services, and receive benefits, housing and mental health services, although few entered employment or education. Reoffending was significantly lower in the RESET group, and further follow-up reduced offending, although non-significant, was reported.

A final model aimed at reducing offending was the **Thinking for a Change (T4C)** intervention (A. B. Wilson, Farkas, Bonfine, & Duda-Banwar, 2018), delivered by ***mental health and care staff*** (community mental health practitioners and social workers with training in facilitating group sessions). The training covers social skills, cognitive restructuring and problem-solving.

#### Citizenship and multi-domain models

|  |  |  |  | **Broad types of support** | | | | |
| --- | --- | --- | --- | --- | --- | --- | --- | --- |
| **Main aim** | **Example models** | **Brief description** | **Source(s) of support** | **Signposting and information** | **Support and skills development** | **Direct resource provision** | **Increased rights and entitlement** | **More informal support** |
| Provide a place where people can go to meet social needs | Mental health day centres | Local centres funded by social services and the voluntary sector which offer patients a "drop in" facility, cheap food, therapeutic groups, member's meetings, social activities and advice and support. | Mental health and care staff |  | X |  |  | X |
| Provide a place where people can go to meet social needs | Clubhouses | Designed to transition individuals from hospital to community living and to address concerns of social isolation, readjustment to society, and community integration. They provide settings designed to foster social connections and community integration including employment opportunities, housing support, case management and social programs for individuals living with schizophrenia and other psychiatric conditions | Mental health and care staff |  |  | X |  | X |
| Provide a place where people can go to meet social needs | Recovery Colleges | Take an educational rather than a clinical or rehabilitation approach to improving mental health and there is an emphasis on co-production, co-delivery and co-participation in the learning. They provide educational platforms to help self-directed recovery and provide learning opportunities for people with mental health problems. They also aim to put people back in control of their lives, increase confidence and skills and provide support for accessing further opportunities. . | Mental health and care staff, Peer workers |  | X |  |  |  |
| Provide a place where people can go to meet social needs | Empowerment of Mental Illness Service Users: Lifelong Learning and Action (EMILIA) | Training related to mental health and social inclusion, personal development and planning, employment and recovery, as well as opportunities for unpaid and paid activities mainly within the demonstration sites themselves | Peer workers |  | X | X |  |  |
| Provide a time-limited therapeutic support programme | The Citizens/Citizenship Project | A 5-month programme involving formal and informal contacts with Peer Mentors and staff, group classes on social participation and community integration, projects to foster valued social roles and peer support. The model theorizes that greater community integration will reduce offending and drug use. | Mental health and care staff, Peer workers | X | X |  |  |  |
| Provide a time-limited therapeutic support programme | Horyzons Project | A digital platform combining: peer-to-peer social networking; theory-driven and evidence-informed therapeutic interventions targeting social functioning, vocational recovery and relapse prevention; expert clinician and vocational support; and peer support and moderation | Mental health and care staff, Peer workers |  | X |  |  | X |
| Provide signposting and navigation | Shared Lives | Providing up to two years of support for young people or adults transitioning from care, matching people with a Shared Lives carer who supports them to move in and share family and community life. | Lay people and organisations |  |  |  |  | X |
| Provide signposting and navigation | The Wiltshire Mental Health Inclusion Service | One-to-one support with an inclusion coach to identify hobbies and interests as well as barriers to these | Domain specialists | X | X |  |  |  |
| Provide signposting and navigation | Social Welfare Legal Services in Health Settings | Provide specialist legal advice and assistance, within a healthcare setting, e.g. issues relating to housing and homelessness, welfare benefits, debt, employment and family issues, which can have an impact on physical and mental health and wellbeing | Domain specialists | X |  | X |  |  |
| Provide signposting and navigation | Mental Health Navigation | A model of support for people with mental illness who present at primary care with unmet non-clinical needs. This model builds on the well-established social prescribing model, offering a dedicated capacity to assist and empower people to access the right support, and manage a range of needs and social distresses that can affect an individual’s mental health. | Domain specialists | X |  |  |  |  |
| Provide signposting and navigation | Community-Enhanced Social Prescribing | A new model of social prescribing combining community engagement, organisational change and individual-level practice. It aims to improve both community and individual wellbeing by recognising that individuals enrich the health of communities and developing opportunities for more active engagement with them. | Lay people and organisations (members of a “citizens panel”),  Domain specialists | X |  |  |  |  |
| Provide signposting and navigation | Rethink mental illness community services: Brent mental health service | Offer a befriending service utilising volunteers; peer navigators support; substance misuse support delivered via Change, Grow, Live (CGL) staff; mental health and recovery workshops. | Lay people and organisations, Mental health and care staff |  | X |  |  | X |
| Provide signposting and navigation | Sustaining Tenancies Accommodation and Resettlement (STAR) | Service users are able to access training and employment, and improve their physical and mental health. Support is also provided by the charity Shelter. | Mental health and care staff |  | X | X |  |  |
